# Identifying predictors of neuropathic pain medication prescribing, adherence, and discontinuation: a systematic review and meta-analysis

**DOI:** 10.1101/2025.04.03.25325072

**Authors:** Mia E. Koponen, Dhaneesha N. S. Senaratne, Harry L. Hébert, Blair H. Smith, Lesley A. Colvin

## Abstract

**Background:** Pharmacoepidemiological studies show that large proportions of people with neuropathic pain (NeuP) are not prescribed the medications recommended for NeuP, do not adhere well to these medications, and/or often discontinue the treatment within six months. Identifying what predicts these outcomes can inform a strategy to ensure that people with NeuP are prescribed recommended medication and continue their treatment when appropriate.

**Methods:** We carried out a systematic review and meta-analysis to identify predictors of pain medication prescribing, adherence, and discontinuation in adults with NeuP. Electronic searches were conducted in Embase, PubMed, Web of Science, and CINAHL Plus.

**Results:** We identified 69 relevant studies and divided them into non-mutually exclusive categories based on the outcomes they had investigated. There were 53 studies on prescribing, 14 on adherence, and 27 on discontinuation. Predictors associated with being prescribed recommended NeuP medications included having a mental health disorder, diabetes, and/or white ethnicity. Predictors associated with better adherence included being prescribed serotonin-norepinephrine reuptake inhibitors compared to gabapentinoids or tricyclic antidepressants; dose titration, and/or implementing a medicine reminder. Predictors associated with a higher likelihood of discontinuation included being prescribed tricyclic antidepressants rather than other antidepressants and/or having a combination of NeuP medications rather than monopharmacotherapy.

**Discussion:** There is a need to focus on improving NeuP medication prescribing for ethnic minorities and people without diabetes. Adherence may be improved by using dose titration and/or a medicine reminder. Prescribing monopharmacotherapy rather than combination pharmacotherapy for NeuP may result in better treatment persistence.

**Systematic review protocol:** PROSPERO CRD42023464307.

## Introduction

Neuropathic pain (NeuP) is ‘pain that arises as a direct consequence of a lesion or disease affecting the somatosensory system’ and is associated with unpleasant sensations including those described as burning, pins and needles, electric shocks, and numbness.^1, 2^ NeuP affects around 10% of the general population and has a substantial negative effect on people’s quality of life even compared to those experiencing other types of chronic pain.^3–5^ In addition, common analgesics such as non-steroidal anti-inflammatory drugs (NSAIDs) and opioids that are used to treat nociceptive pain may be ineffective in treating neuropathic pain.^6^ According to the pharmacotherapy guidelines by the International Association for the Study of Pain (IASP) Neuropathic Pain Special Interest Group (NeuPSIG), standard first-line medications for NeuP are gabapentinoids, serotonin-norepinephrine reuptake inhibitors (SNRIs), or tricyclic antidepressants (TCAs).^6, 7^ Second-line NeuP medications include topical capsaicin or lidocaine, and third-line medications include botulinum toxin A or opioids.^6, 7^

Despite the existing treatment guidelines and variety of choices, studies suggest that there are major inconsistencies in NeuP treatment. For example, in a retrospective cohort study conducted in the USA during 2001-2014, only 52.9% of 14,426 people with NeuP were prescribed a first-line medication in the year before or three years after diagnosis.^8^ This suggests that a large proportion of people with NeuP are not being prescribed recommended treatment. Another retrospective cohort study conducted in the USA during 2001-2016 showed that in a sample of 57,495 people with NeuP, adherence to gabapentinoids and SNRIs was on average ∼40-50% and ∼50-60% respectively,^9^ which suggests that adherence to NeuP medications is moderate or poor.^10^ Moreover, in a retrospective cohort study conducted in Sweden during 2004-2009, 60% of 4,718 people with NeuP had discontinued their treatment five months after first prescription.^11^ Other studies have shown that only a small proportion of people who discontinue their initial treatment switch to another medication.^12, 13^

Currently, it is not clear what predicts pharmacotherapy choice for NeuP, or adherence and persistence with treatment. To our knowledge, no published reviews have summarised the literature on the predictors of any of these outcomes. Identifying these predictors may inform strategies to ensure that people with NeuP are prescribed recommended medication and adhere to and persist with their treatment when appropriate. For example, understanding which patient characteristics or healthcare factors are associated with these outcomes could help clinicians target interventions, such as tailored patient education, closer follow-up, or system-level changes to reduce disparities and improve long-term treatment outcomes. Therefore, we carried out a systematic review and meta-analysis to identify predictors of pain medication prescribing, adherence, and discontinuation in adults with NeuP.

## Methods

### Protocol and registration

The protocol of this systematic review and meta-analysis was prospectively registered on the PROSPERO international prospective register of systematic reviews (Registration ID: CRD42023464307). This report follows the Preferred Reporting Items for Systematic Reviews and Meta-analysis (PRISMA) checklist.^14^

### Search strategy

To identify relevant studies, Embase, PubMed, Web of Science, and CINAHL Plus were searched from inception to the 1^st^ of August 2025. The search strategy combined keywords and medical subject headings for 1) neuropathic pain 2) pharmacological treatments and 3) prescribing, adherence, and discontinuation. The full search terms for all the databases are reported in Table S1. The retrieved records were exported to Covidence (Veritas Health Innovation, Melbourne, Australia) and screened independently by two reviewers (MK and DS). Discrepancies were resolved by discussion with the wider review team where necessary. The reference lists of the included studies were reviewed to identify further eligible studies not identified by the search strategy.

### Eligibility criteria

Studies were included if they: (1) investigated potential predictors of pain medication prescribing, adherence, or discontinuation in adults (≥18 years old) with NeuP; (2) were a cohort study, case-control study, cross-sectional study, randomised controlled trial (RCT), or systematic review with meta-analysis; (3) were published in full-text in English. Studies were excluded if (1) they included participants who did not have NeuP, where these could not be analysed separately from those who had NeuP; (2) they included children (<18 years old) and did not present analysis on adults separately; (3) they were a case study, case series, review without a meta-analysis, protocol, editorial, or conference abstract; (4) they were an RCT that aimed to assess NeuP medication efficacy and the only relevant information for our review was discontinuation rates between trial medications; (5) the medications of our interest could not be analysed separately from other medications; and/or (6) they compared pain medications before and after surgery.

It should be noted that although the diagnosis of interest in this review was NeuP, we also included studies that defined NeuP using diagnostic codes for neuropathy, as diagnostic codes used to indicate NeuP often cannot separate painful and painless neuropathy. Our assumption was that relevant pain medicines were only prescribed for neuropathy in these studies when the neuropathy was painful - though we could not rule out the possibility of pain medicines given for other painful conditions or indications, in the presence of a painless neuropathy. We focused on peripheral neuropathic pain conditions, but did not exclude studies where central neuropathic pain conditions could not be separated from peripheral neuropathic pain conditions. The method of diagnosis used in each study is provided in Table S2. For the above discussed reason and because of the inconsistency of terms used for NeuP caused by diabetes, the following terms used in included papers, were grouped under diabetic neuropathy (DN): diabetic peripheral neuropathic pain (DPNP), diabetic peripheral neuropathy (DPN), painful diabetic peripheral neuropathy (pDPN or PDPN), painful diabetic neuropathy (PDN), painful diabetic polyneuropathy, diabetic polyneuropathy.

The medication categories of interest were anticonvulsants, antidepressants, opioids, non-opioid analgesics, combinations of opioids and non-opioid analgesics, local anaesthetics, N-methyl-D-aspartate (NMDA) receptor antagonists, topical pain medications, botulinum toxin A, baclofen, and clonidine. The medications that were not of interest included but were not limited to muscle relaxants, corticosteroids, sedatives, and antimigraine medications. A summary of medications included and excluded from our review is provided in Table S3, while Table S4 provides medication included and excluded in each study. In addition, Table S5 shows medications considered recommended in the included studies. In our review, the word ‘non-opioid analgesics’ refers to non-steroidal anti-inflammatory drugs (NSAIDs), paracetamol, aspirin, and nefopam. This categorisation was used as some of the included studies categorised their medications as antidepressants, anticonvulsants, opioids, local anaesthetics, and non-opioid analgesics, without precisely defining the ‘non-opioid analgesics’ category.^15–18^

### Data extraction

Data extraction was performed independently by two reviewers (MK and DS) and discrepancies were resolved by discussion. Study characteristics extracted included: author, year of publication, type of outcome(s) investigated, study design, country and time frame of data analysed, sample, size, proportion of females in the sample, mean age, NeuP diagnoses included in the study, and key findings.

### Risk of bias assessment

Risk of bias assessment was performed independently by two reviewers (MK and DS) and discrepancies were resolved by discussion. The Risk Of Bias In Non-randomized Studies - of Exposures (ROBINS-E)^19^ tool was used to assess cohort studies, cross-sectional studies, and case-control studies. The “exposure” in each study was defined as the potential predictors investigated (e.g. NeuP diagnosis, sex, household income). The revised Cochrane Risk of Bias tool for randomized trials (RoB 2)^20^ was used to assess three RCTs. A secondary analysis of RCTs was assessed using A MeaSurement Tool to Assess systematic Reviews 2 (AMSTAR 2).^21^

### Synthesis of results

The included studies were grouped into non-mutually exclusive categories based on the outcomes they investigated (prescribing, adherence, and discontinuation). The studies that investigated prescribing were further divided into non-mutually exclusive subgroups based on which medications were examined and how the analyses were conducted. For each category of studies, we listed all the potential predictors investigated and summarised the evidence for each potential predictor. If the potential predictor definition and outcome measurement were similar or could be converted to be similar, meta-analyses were conducted using RStudio (v4.3.1). Statistical heterogeneity was described using the I^2^ test. Random-effects models were used for all meta-analyses. The type of measure of effect estimated in each meta-analysis depended on the type of data provided in the included studies for the potential predictors. In the majority of cases, odds ratios (OR) and the associated 95% confidence intervals (CI) were calculated. In meta-analyses that were conducted using pre-calculated ORs, adjusted ORs were used rather than unadjusted ORs if possible. Finally, each potential predictor was categorised either to increase the likelihood of the investigated outcome, to not have an association with it, or to unclear or inconsistent results. Due to the extensive quantity of data generated, in this manuscript we have focused on describing the predictors that we considered to be the most useful to the readers and/or clinicians.

## Results

### Study selection

Figure 1 shows a flowchart of the study selection process. Database searches identified a total of 11,255 records. Sixty-nine studies were included in the review, and this included 6 studies identified from screening the reference lists of the eligible studies. The records excluded in full-text screening and the reasons for their exclusion are documented in Table S7.

**Figure 1.**
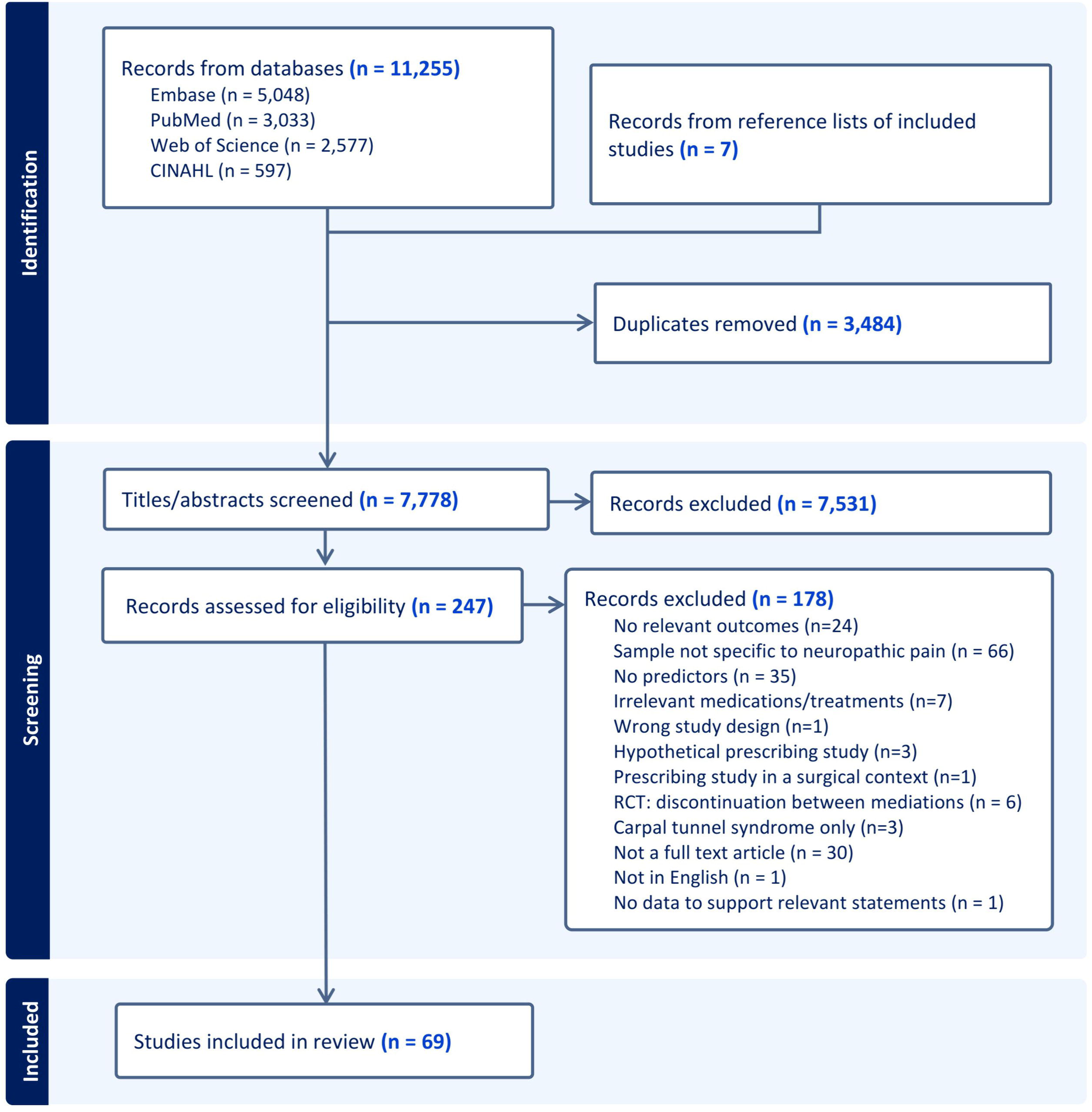
Flowchart of the study selection process. Out of 7,778 titles and abstracts, 69 studies were included in our review.

### Study characteristics

The characteristics of the included studies are summarised in Table 1. The majority of the included studies were retrospective cohort studies (44/69) conducted in North America and Europe (58/69). There were 53 studies on prescribing, 15 on adherence, and 27 on discontinuation. The division of the prescribing studies into different subcategories is shown in Table S7. Table S8 and Table S9 explain how adherence and discontinuation was assessed in each study. Most studies included people with diabetic neuropathy (DN) along with other aetiologies (55/69) – with 24/69 studies exclusively on DN.

**Table 1.**
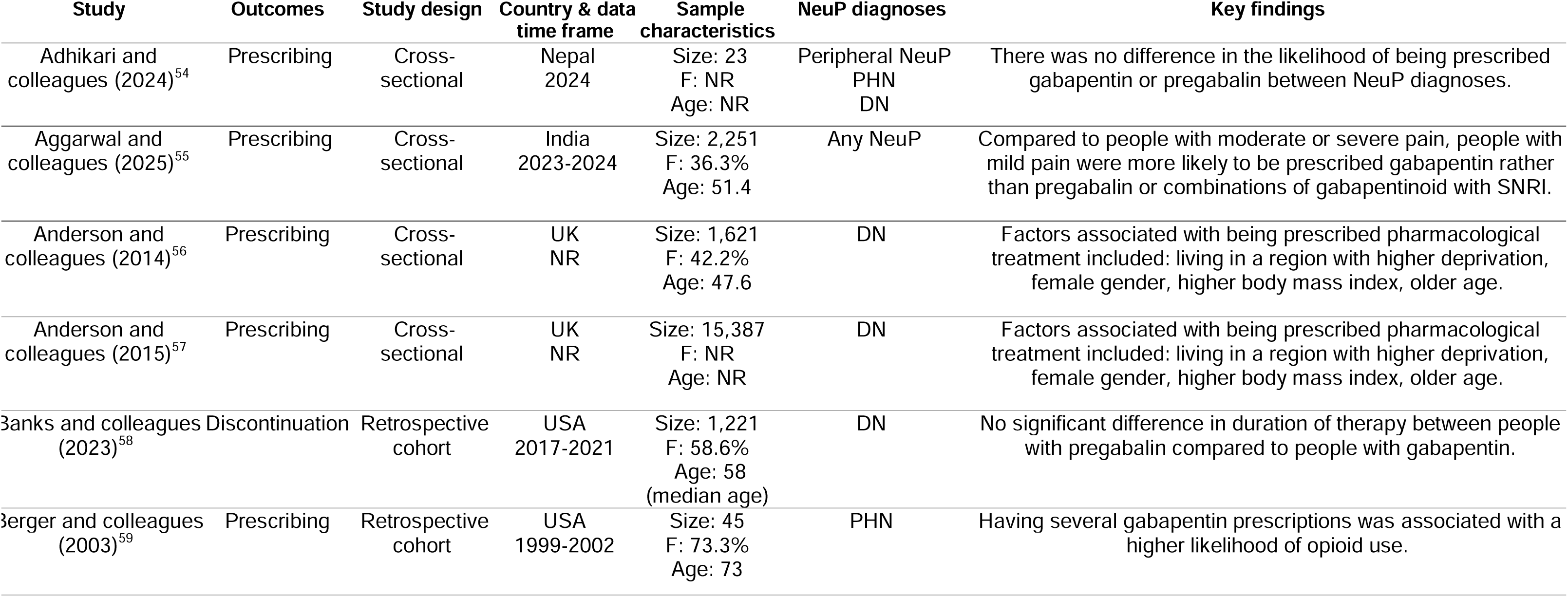
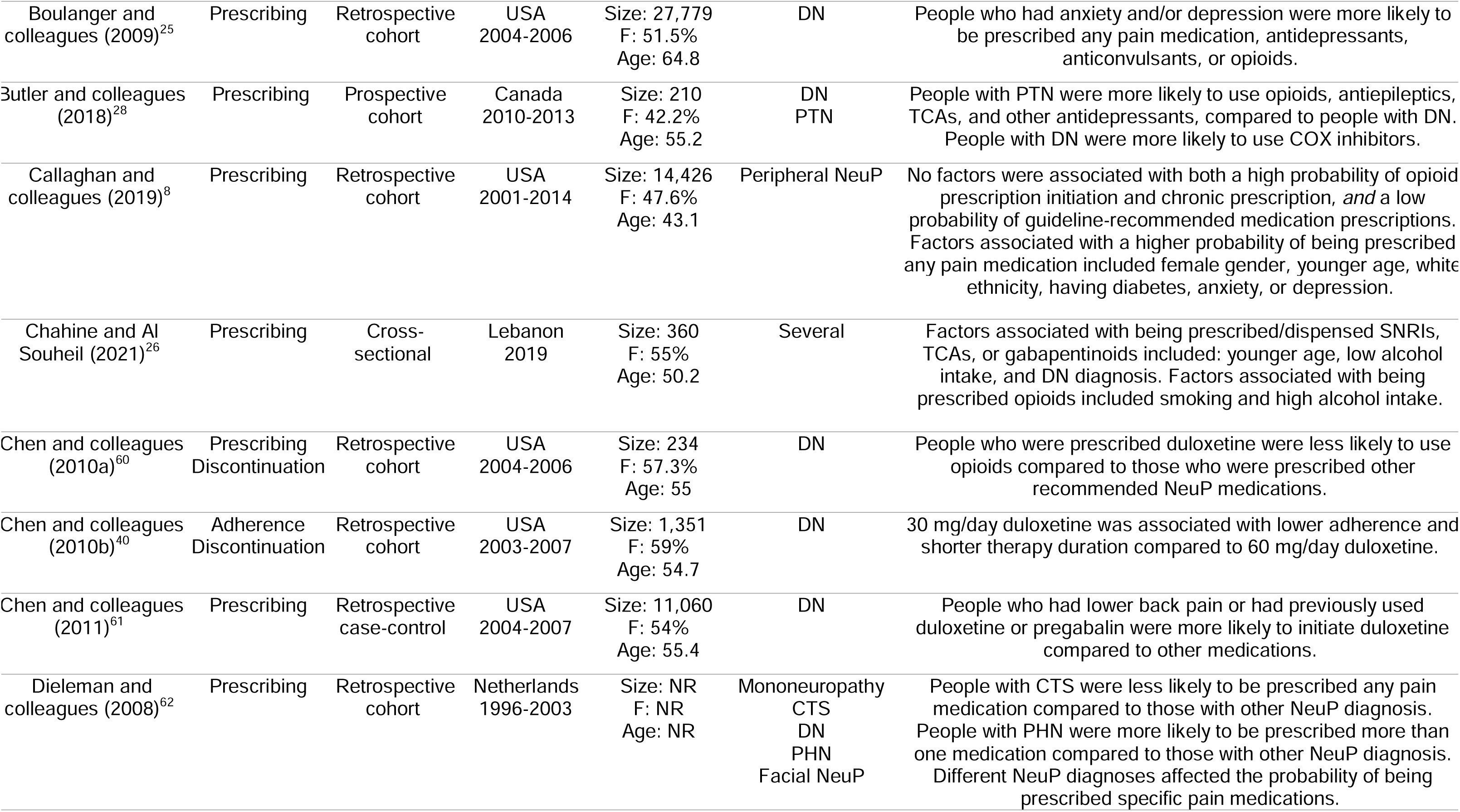
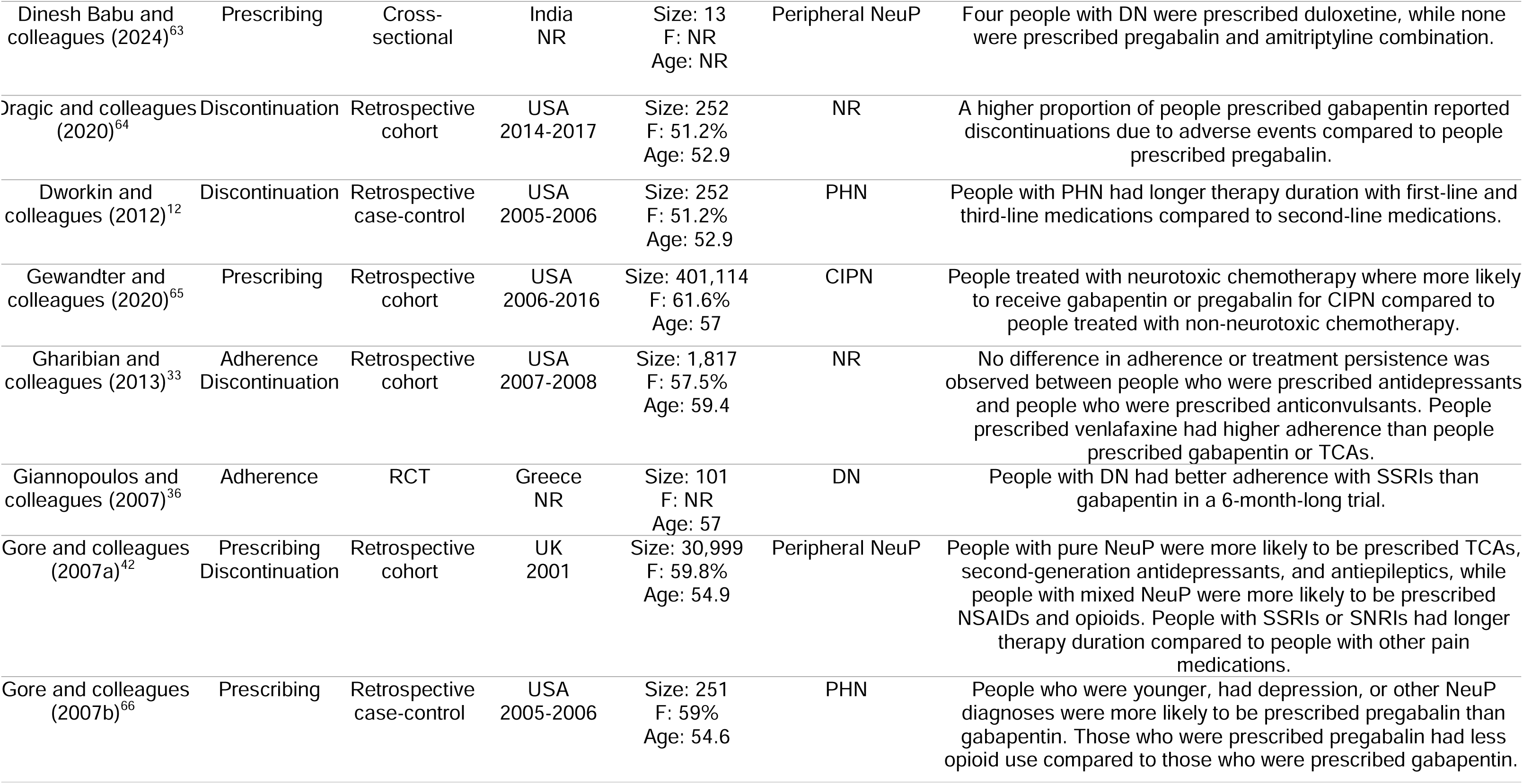
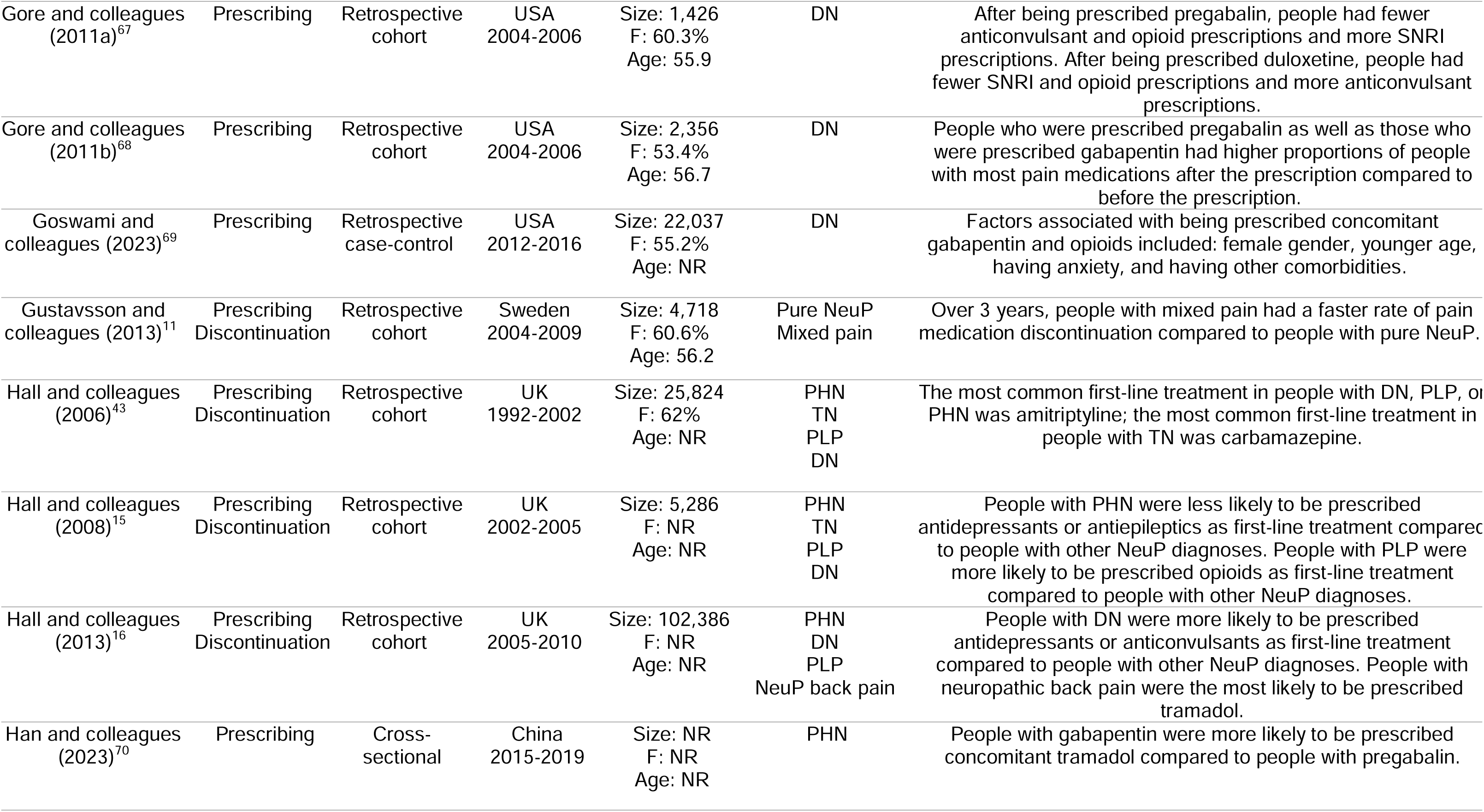
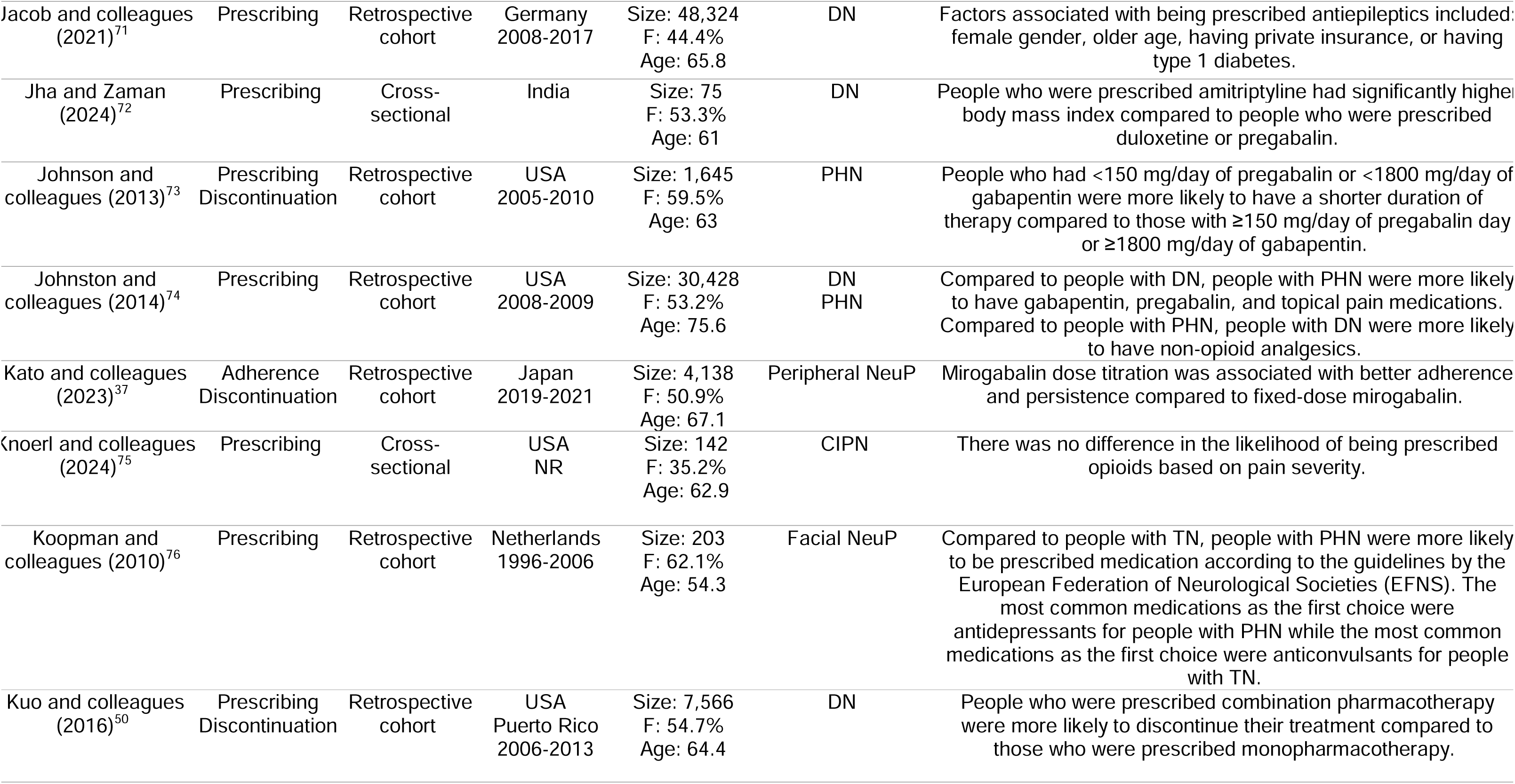
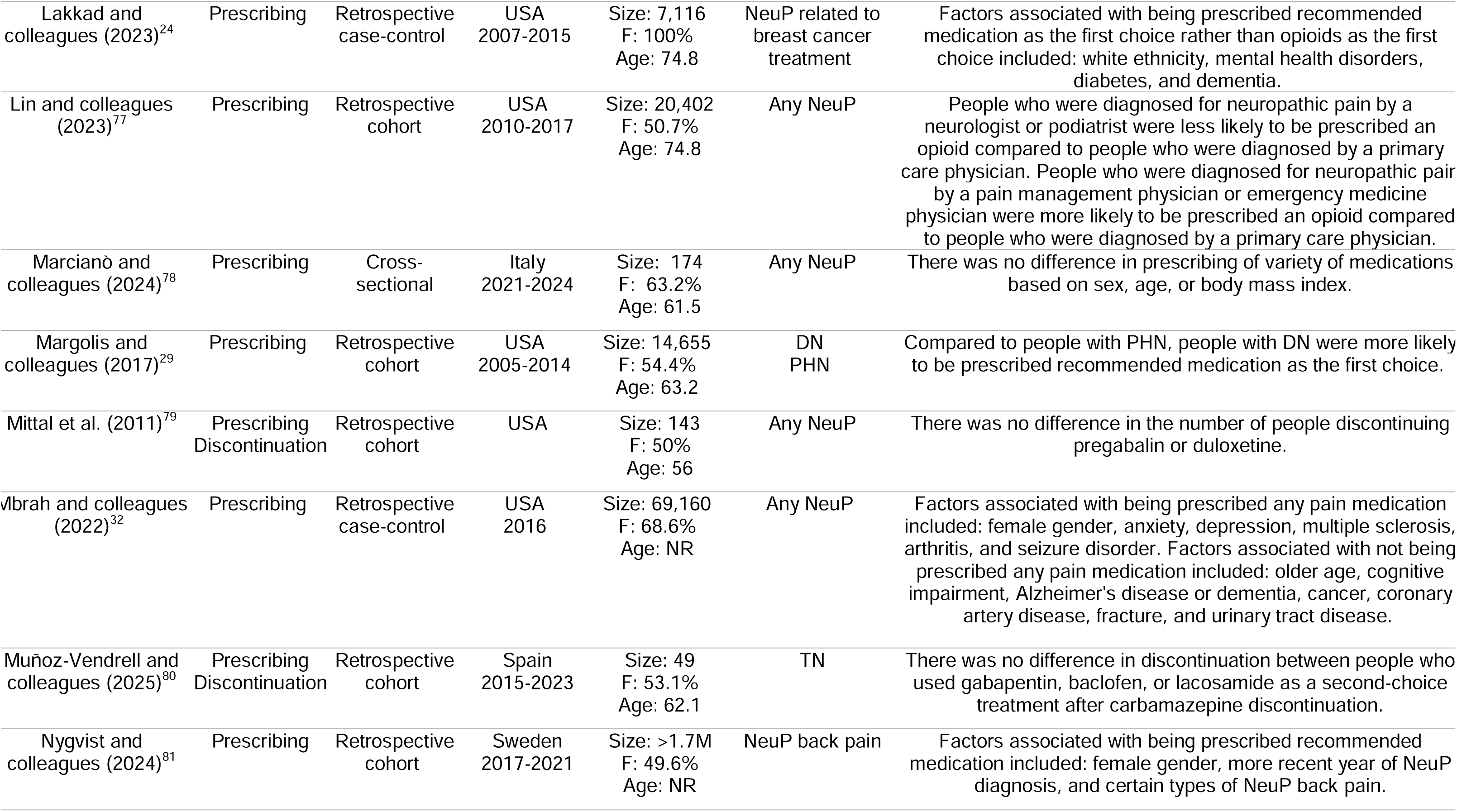
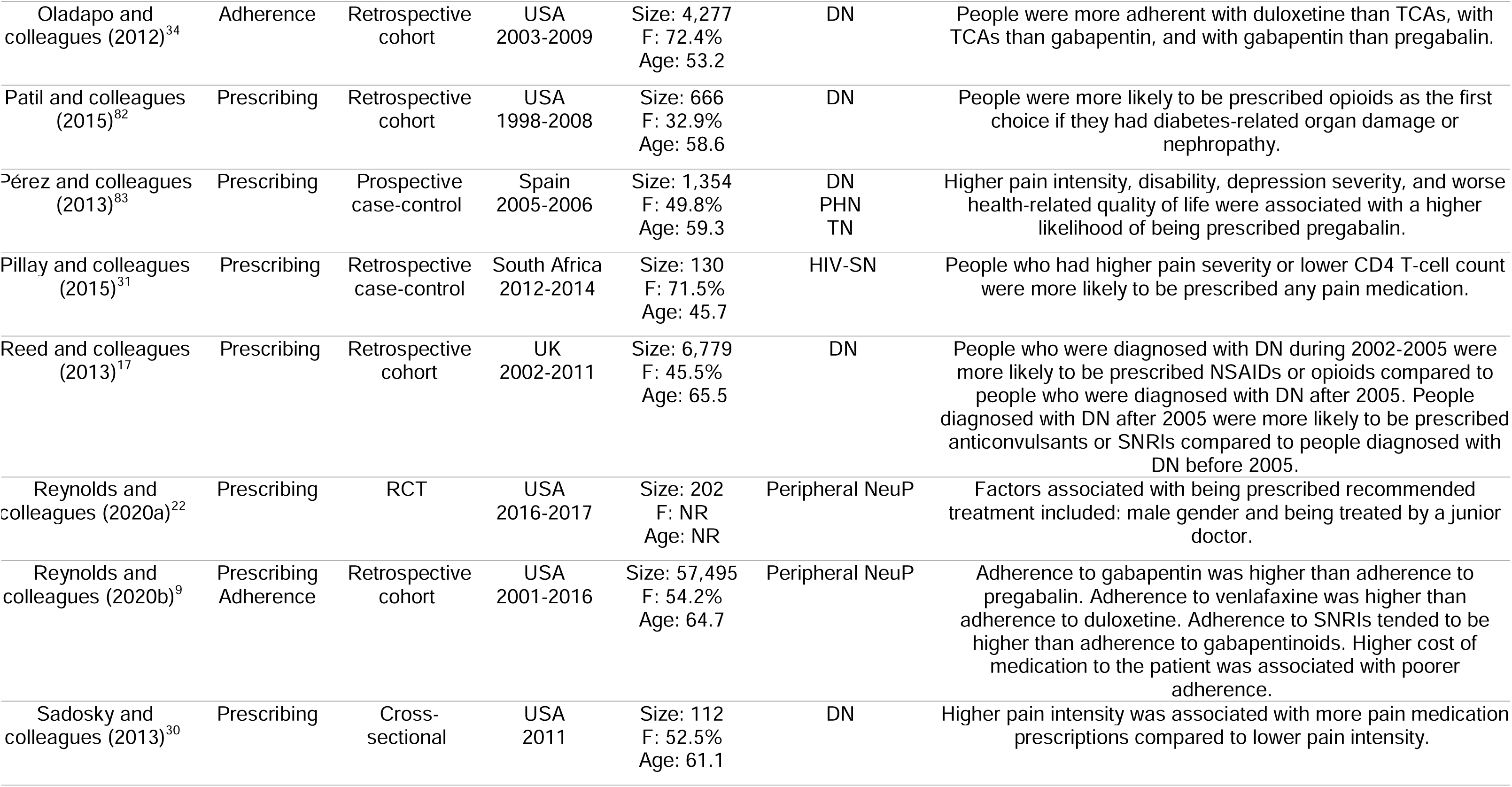
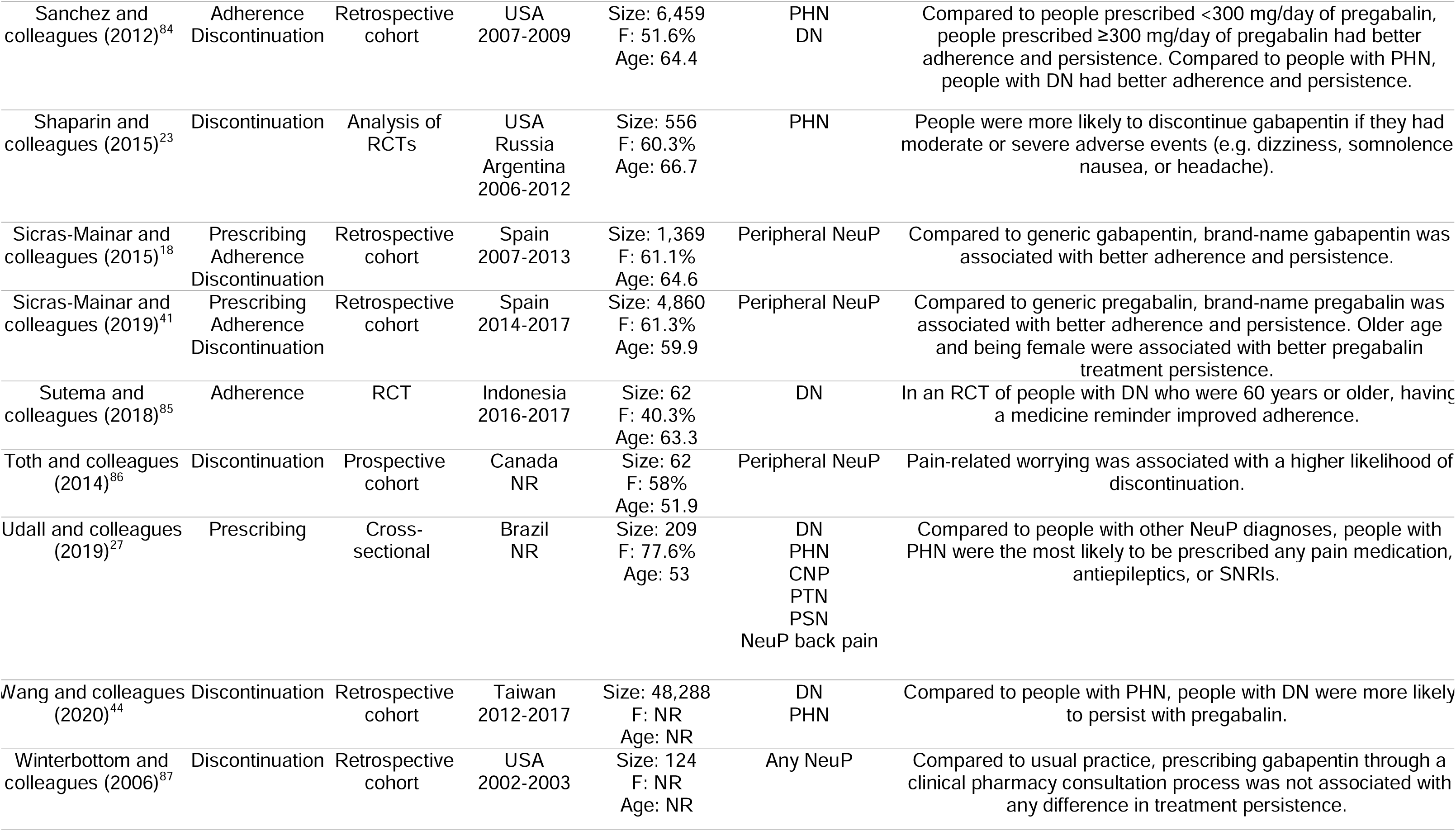
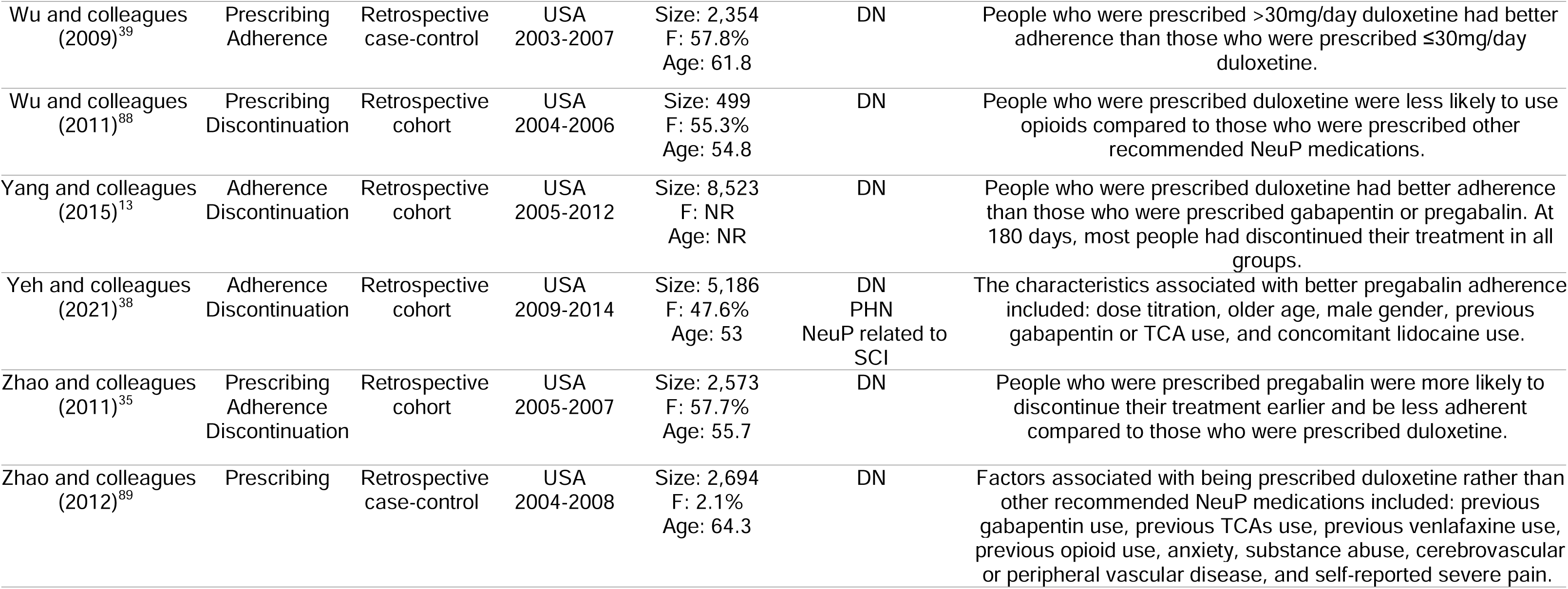
Characteristics of the included studies. Age is reported as mean unless stated otherwise. F: female, NR: not reported, NeuP: neuropathic pain, DN: diabetic neuropathy, PHN: postherpetic neuralgia, CTS: carpal tunnel syndrome, TN: trigeminal neuralgia, PLP: phantom limb pain, CIPN: chemotherapy-induced peripheral neuropathic pain, PSN: post-surgical neuropathic pain, PTN: post-traumatic neuropathic pain, HIV-SN: human immunodeficiency virus-associated sensory neuropathy, SCI: spinal cord injury, COX: cyclooxygenase, TCA: tricyclic antidepressant, SNRI: serotonin and norepinephrine reuptake inhibitor, SSRI: selective serotonin reuptake inhibitor.

### Risk of bias assessment

The majority of the studies (65/69) were assessed using ROBINS-E. The results of this assessment are shown in Table S10. All of the studies assessed using ROBINS-E were deemed to have a low risk of bias in measuring exposures, post-exposure interventions, outcome measurement, and reporting of results (although the domain “post-exposure interventions” was generally irrelevant to our research question). Sixteen studies were deemed to have a high risk of bias in “confounding variables” due to adjusting for potential predictors that could affect each other. In the participant selection domain, most of the studies had “some concerns” as the exposure window of some of the investigated potential predictors would be lifelong (e.g. age or sex) and thus would precede the data collection time frame. However, these factors were unlikely to influence our interpretation of these studies. In the missing data domain, in many of the studies with a high risk of bias, no information was provided about the handling of missing data. Three RCTs were assessed using Rob 2. The results are shown in Table S11. In these studies, the only finding of a high risk of bias was in the missing data domain in one of the studies^22^ as there was no information about missing data. One study, a non-systematic analysis of three RCTs, ^23^ was assessed using AMSTAR 2 and was concluded to have a critically low (poor) overall rating. The results for each domain are shown in Table S12.

### Summary of results

The analyses of all potential predictors, including meta-analysis results, are provided Tables S13 to S20. A summary of these results is provided in Table S21, which categorises all potential predictors to either to increase the likelihood of the investigated outcome, to not have an association with it, or to unclear or inconsistent results. The key findings are summarised in Table 2, which provides examples of association strength between the predictor and the outcome. Other relevant results for the key predictors are described below.

**Table 2.** Summary of key predictors. For each predictor, an example result is reported. OR: odds ratio, PR: prevalence ratio, IRR: incidence rate ratio, MPR: medication possession ratio, PDC: proportion of days covered, RR: relative risk, HR: hazard ratio.

| Outcome | Key predictors | Example result |
| --- | --- | --- |
| Receiving recommended NeuP medication | White ethnicity | OR: 1.40 (95% CI: 1.18 to 1.66)<sup>8, 24</sup> |
|  | Diabetes | OR: 1.41 (95% CI: 1.23 to 1.62)<sup>8, 24, 26</sup> |
|  | Mental health disorder (especially depression and/or anxiety) | OR: 3.57 (95% CI: 3.12 to 4.09)<sup>24</sup> |
|  | Pure NeuP (compared to mixed pain) | OR: 1.37 (95% CI: 1.30 to 1.45)<sup>42</sup> |
| Receiving any pain medication | Younger age (50-64 years vs +85 years) | PR: 1.45 (95% CI: 1.38 to 1.52)<sup>32</sup> |
|  | Higher pain severity (BPI severe vs mild) | OR: 25.83 (95% CI: 2.97 to 225.21)<sup>30</sup> |
|  | Anxiety and/or depression | OR: 3.66 (95% CI: 3.14 to 4.26)<sup>25</sup> |
| Adherence | Older age (55-64 years vs 18-44 years) | OR: 2.11 (95% CI: 1.50 to 2.97)<sup>39</sup> |
|  | Younger age (65-74 years vs 75-84 years) | OR: 1.33 (95% CI: 1.01 to 1.79)<sup>39</sup> |
|  | White ethnicity | IRR: 1.14 (95% 1.08 to 1.21)<sup>9</sup> |
| | Lower medication cost to patient in the USA (compared to +\$50 for 30-day supply) | IRR: 1.09 (95% CI: 1.03 to 1.14)<sup>9</sup> |
|  | SNRIs (compared to gabapentinoids) | Difference in mean MPR: >0.10<sup>9</sup> |
|  | SNRIs (compared to tricyclic antidepressants) | Difference in mean MPR: 9.6 %<sup>34</sup> |
|  | Dose titration | OR: 2.40 (95% CI: 1.51 to 3.21)<sup>37, 38</sup> |
|  | Brand-name medication (compared to generic-name medication) | MD in MPR: 0.05 (95% CI: 0.03 to 0.07)<sup>18, 41</sup> |
|  | Medicine reminder | RR: 2.38 (95% CI: 1.58 to 3.61)<sup>85</sup> |
|  | Diabetic neuropathy (compared to postherpetic neuralgia) | MD in PDC: 0.13 (95% CI: 0.10 to 0.16)<sup>84</sup> |
| Discontinuation | Tricyclic antidepressants (compared to other antidepressants) | OR: 1.56 (95% CI: 1.01 to 2.42)<sup>15</sup> |
|  | Generic rather than brand name medication | HR: 1.43 (95% CI: 1.18 to 1.72)<sup>41</sup> |
|  | Combination pharmacotherapy | HR: 1.30 (95% CI: 1.13 to 1.51)<sup>50</sup> |
|  | Not experiencing clinically significant pain relief | OR: 1.2 (95% CI: 1.0 to 1.4)<sup>41</sup> |
|  | Experiencing moderate adverse events | OR: 8.64 (95% CI: 4.42 to 16.89)<sup>23</sup> |
|  | Experiencing severe adverse events | OR: 5.37 (95% CI: 2.01 to 14.00)<sup>23</sup> |
|  | Postherpetic neuralgia (compared to diabetic neuropathy) | MD in treatment days: 51 (95% CI: 4 to 98)<sup>15, 16, 43, 44</sup> |
|  | Pain-related worrying (CSQ catastrophising subscale score) | Pearson's correlation: 0.36<sup>86</sup> |

It should be noted that studies that investigated potential predictors of NeuP medication prescribing often did not report their analysis such that the participants were exclusively prescribed the medication that was analysed and no other pain medications. For example, if potential predictors of being prescribed recommended medication were analysed, it was not reported whether these people received other pain medications.

### Receiving recommended NeuP medication

The key predictors for receiving recommended NeuP medication were having a mental health disorder (in particular anxiety and/or depression), diabetes, white ethnicity, and/or purely neuropathic pain rather than mixed pain.

Mental health disorders as predictors for receiving recommended medication were investigated in four studies. Lakkad and colleagues (2023) found that people with a mental health disorder (a variety of diagnoses included) were more likely to receive recommended NeuP medication (OR: 3.57, 95% CI: 3.12 to 4.09).^24^ Boulanger and colleagues (2009) (N=27,779) found that people with anxiety and/or depression were more likely to receive recommended medication (OR: 1.74, 95% CI: 1.63 to 1.86).^25^ Callaghan and colleagues (2019) and Chahine and Al Souheil (2021) investigated anxiety and depression separately, and meta-analyses based on their results found that people with depression were more likely to receive recommended medication (OR: 1.62, 95% CI: 1.33 to 1.98), while the result for anxiety was no significant (OR: 0.84, 95% CI: 0.20 to 3.59).^8, 26^ It is worth noting that the result from Chahine and Al Souheil (2021) on anxiety was the only non-significant result, and their study was much smaller (N=360) than the study by Callaghan and colleagues (2019) (N=14,426).

Eight studies investigated whether having diabetes or diabetic neuropathy changed the likelihood of receiving recommended NeuP medication. A meta-analysis based on three studies investigating diabetes found that people with diabetes were more likely to receive recommended medication (OR: 1.41, 95% CI: 1.22 to 1.62).^8, 24, 26^ The results relating to DN compared to other NeuP aetiologies were mixed, although the results generally showed that either people with DN were more likely to receive recommended medication or that there was no difference between the aetiologies.^15, 16, 26–29^ Only people with TN were found to be more likely to receive recommended medication than people with DN.^15^

Ethnicity as a potential predictor for receiving recommended medication was investigated in three studies. Lakkad and colleagues (2023) (N=7,116) found that people with white ethnicity were more likely to receive recommended medication compared to people with black ethnicity (OR: 1.76, 95% CI: 1.43 to 1.75) or any other ethnicity (OR: 1.37, 95% CI: 1.07 to 1.75).^24^ In contrast, Reynolds and colleagues (2020a) (N=202) found no differences based between these categories.^22^ Callaghan and colleagues (2019) found that people with white ethnicity were more likely receive recommended medication compared to people with Asian (OR: 1.64, 95% CI: 1.22 to 2.17) or Hispanic ethnicity (OR: 1.32, 95% CI: 1.12 to 1.52), but found no difference between white and black ethnicity (OR: 1.11, 95% CI: 0.97 to 1.27).^8^ A meta-analysis contrasting white and non-white groups found that people with ethnicity were more likely to receive recommended medication (OR: 1.40, 95% CI: 1.18 to 1.66).

### Receiving any pain medication

The key predictors for receiving any pain medication were higher pain severity, having anxiety and/or depression, and/or younger age (among people aged >50 years).

Sadosky and colleagues (2013) (N=112) investigated how pain severity measured using the Brief Pain Inventory (BPI) influenced the likelihood of receiving any pain medication.^30^ They found that the proportion of people with medication increased as pain severity increased (54.5% in mild pain, 84.2% in moderate pain, 96.9% in severe pain). In this small sample, the difference between severe pain and mild pain was very large (OR: 25.83, 95% CI: 2.97 to 225.21). Another small study investigated the differences in pain intensity measured using a 0-10 numerical rating scale (NRS).^31^ In this study, Pillay and colleagues (2015) (N=130) found that the median pain intensity in the group with medication was significantly higher than in the group without medication [Median NRS (IQR): 10 (2-10) vs 6 (2-10)].^31^

Two studies investigated whether having a mental health disorder changed the likelihood of receiving any pain medication. Boulanger and colleagues (2009) (N=27,779) found that people with anxiety and/or depression were more likely to receive any pain medication (OR: 3.66, 95% CI: 3.14 to 4.26).^25^ Mbrah and colleagues (2022) investigated these separately and found similar results (adjusted prevalence ratio for receiving medication, for depression: 1.47, 95 % CI: 1.43 to 1.52; for anxiety: 1.20, 95% CI: 118 to 1.23).^32^

The differences in the likelihood of receiving any pain medication based on age were investigated by Mbrah and colleagues (2022) and Pillay and colleagues (2015). Mbrah and colleagues (2022) (N=69,160) investigated people in nursing homes who were over 50 years old. They compared different age categories and found that younger people were more likely to receive medication (50-64 years vs +85, prevalence ratio: 1.45, 95% CI: 1.38 to 1.52).^32^ Pillay and colleagues (2015) (N=130) found no difference in age between the groups that received vs did not receive medication [mean (SD) age in years: 45.4 (9.3) vs 46.2 (9.8), p=0.63].^31^ The differing results may be due to differences in the ages analysed or due to the small sample size of the latter study.

### Adherence

The key predictors for having better adherence to NeuP medications were taking SNRIs rather than gabapentinoids or TCAs, having dose titration, brand-name medication, a medicine reminder, lower medication cost to the patient (USA), diabetic neuropathy rather than postherpetic neuralgia, older age (in below 75-year-olds), and/or white ethnicity. Most of the adherence studies measured the outcome using medication possession ratio (MPR) or proportion of days covered (PDC) [See Table S6].

Six studies investigated the differences in adherence between NeuP medications.^9, 13, 33–36^ Reynolds and colleagues (2020b) had the largest sample (N=57,495) and longest duration of follow-up (12 years for this data).^9^ They found that the mean MPR for gabapentinoids was around 40-50% and for SRNIs 50-60%.^9^ Three other studies compared SNRIs and gabapentinoids and found better adherence to SNRIs.^13, 33, 34^ Gharibian and colleagues (2013) found a large difference in adherence between venlafaxine and gabapentin (OR: 2.83, 95% CI: 1.45 to 5.76).^33^ Two studies compared adherence between SNRIs and TCAs. Oladapo and colleagues (2013) found that mean MPR in people taking duloxetine was 85.6% while the mean MPR in people taking TCAs was 76.0% (n=652 vs n=931, p<0.0001).^34^ Similarly, Gharibian and colleagues (2013) found that the proportion of people with ≥ 80% MPR was 69.4% in people taking venlafaxine and 44.4% in people taking TCAs (n=49 vs n=1208, p<0.01).^33^

The difference in adherence between groups receiving or not receiving dose titration was investigated in two studies. Meta-analysis of these studies found that people who received any dose titration within 45 days of initiation their medication had better adherence compared to people with no dose titration (OR: 2.20, 95% CI: 1.51 to 3.21).^37, 38^

Five studies examined adherence based on age. Wu and colleagues (2009) found that people aged 55-64 years had better adherence than people aged 18-44 years (OR: 2.12, 95% CI: 1.30 to 3.46), while there was no difference between people aged 45-54 years and 18-44 years (OR: 1.63, 95% CI: 0.99 to 2.71).^39^ The other studies that investigated age supported that increasing age was associated with better adherence.^9, 37, 38, 40^ Wu and colleagues (2009) were the only ones to examine adherence in people aged 65 years and older.^39^ They found that people aged 65-74 years had better adherence than people aged 75-84 years (OR: 1.33, 95% CI: 1.01 to 1.79).^39^ Thus, increasing age seems to be associated with better adherence until the age of 75 years, after which it may decline.

Two studies from Spain found that people with brand-name gabapentin or pregabalin had better adherence compared to people with generic name gabapentin or pregabalin (Mean MPR MD: 0.05, 95% CI: 0.03 to 0.07).^18, 41^ In contrast, a study from the USA found that lower medication cost to patient was associated with better adherence (IRR: 1.09, 95% CI: 1.03 to 1.14 when +$50 for 30-day supply).^9^

### Discontinuation

The key predictors for being more likely to discontinue NeuP medications were taking TCAs rather than other antidepressants, having generic-name medication rather than brand-name medication, combination pharmacotherapy for NeuP rather than monopharmacotherapy, not experiencing clinically significant pain relief from the medication, experiencing moderate or severe adverse effects from the medication, having postherpetic neuralgia rather than diabetic neuropathy, and/or pain-related worrying.

Only two eligible studies investigated the clinical effectiveness of NeuP medications or their adverse effects as potential predictors of discontinuation. Sicras-Mainar and colleagues (2019)^41^ (N=4,860) found that people who did not experience clinically meaningful pain relief measured using a visual analogue scale (≥4 out of 0-10 points scale) were more likely to discontinue pregabalin (OR: 1.2, 95% CI: 1.0 to 1.4). Shaparin and colleagues (2015)^23^ (N= 556) found that people with PHN taking gabapentin were more likely to discontinue if they experienced moderate (OR: 8.64, 95% CI: 4.42 to 16.89) or severe adverse events (OR: 5.37, 95% CI: 2.01 to 14.00).

The differences in therapy duration between medications were investigated by eleven studies. Most of these compared individual medications (e.g. pregabalin vs duloxetine). Two studies provided useful comparisons between different medication classes. Hall and colleagues (2008)^15^ found that people taking TCAs were more likely to not have stable pharmacotherapy after one year compared to people using other types of antidepressants (OR: 1.56, 95% CI: 1.01 to 2.42). Similarly, Gore and colleagues (2007a)^42^ found that people taking TCAs had fewer mean days with pharmacotherapy compared to people with SNRIs (MD -12, 95% CI: -21 to -3) or SSRIs (MD -35, 95% CI: -40 to -30). Heatmaps showing the pairwise meta-analyses between all the investigated medications in these studies are shown in Figures S1 and S2.

Four studies compared pharmacotherapy durations between people with PHN and people with DN. Our meta-analysis found that people with PHN were more likely to discontinue sooner than people with DN (MD in treatment days: -51, 95% CI: -4 to -98). Three of the included studies^15, 16, 43^ found a mean difference in treatment days to be around -20 to -30 days, while Wang and colleagues (2020)^44^ found a mean difference of -133 days (95% CI: 126 to 139).

## Discussion

Our systematic review and meta-analysis aimed to understand the predictors of NeuP pharmacotherapy prescribing, adherence, and discontinuation. In the 69 included studies, prescribing was analysed in 53 studies, adherence in 14 studies, and discontinuation in 27 studies. They key predictors for receiving recommended medication were having diabetes and/or white ethnicity. People with SNRIs, dose titration and/or medicine reminder were more likely to have better adherence. Pain-related worrying and/or combination pharmacotherapy were associated with shorter treatment durations.

The association between diabetes and receiving recommended medication is not unexpected. There are many potential contributing factors to this. Diabetes is the most common cause for peripheral neuropathic pain and is widely recognized in the clinical and research communities.^45^ People with diabetes usually have regular appointments for diabetes management, which increases the chance of neuropathic symptoms being detected and opportunities for medication review.

Similarly, ethnic differences in prescription medicines have been noted in Western countries for a long time.^46, 47^ Socioeconomic status is often considered to be the key explanatory variable between health care and ethnicity.^48^ However, in many cases disparities by ethnicity cannot be explained by socioeconomic status or other factors.^48^ In a key study included in our review, Callaghan and colleagues (2019) found that people with white ethnicity were more likely to receive recommended medication compared to people with Asian or Hispanic ethnicity despite controlling for education, household income, or insurance type.^8^

In the current IASP NeuPSIG pharmacotherapy guidelines (which are based on results from RCTs) SNRIs, TCAs, and gabapentinoids are recommended as first-line treatments.^6^ The number needed to treat (low indicating better efficacy) is lower for TCAs (4.6, 95% CI: 3.2 to 7.7) compared to that of SNRIs (7.4, 95% CI: 5.6 to 10.9) or gabapentinoids (8.9, 95% CI: 7.4 to 1.11).^6^ The number needed to harm (low indicating more risk) is lowest for SNRIs (13.9, 95% CI: 10.9 to 19.0), a little bit higher for TCAs (17.1, 95% CI: 11.4 to 33.6), and considerably higher for gabapentinoids (26.2, 95% CI: 20.4 to 36.5).^6^ It is also noteworthy that TCAs are not recommended for older adults due their anticholinergic and sedative side effects. Interestingly, the studies included in our review (mostly epidemiological), found that adherence and persistence with TCAs was lower compared to SNRIs.^15, 33, 34, 42^ In addition, adherence to SNRIs was better compared to adherence to gabapentinoids.^9, 13, 33–35^ Reynolds and colleagues (2020b) were a key study for this due to their large sample (n=57,495).^9^ Since these results come from epidemiological studies, it is possible that these results are confounded by variables that are not evident. On the other hand, RCTs cannot provide real life evidence, in particular for outcomes like adherence or persistence.

Dose titration and combination pharmacotherapy, which were identified as key predictors for adherence and discontinuation respectively, have been discussed in the IASP NeuPSIG pharmacotherapy guideline.^6^ The studies included in our review found that people who received any dose titration within 45 days of medication initiation were more likely to adherence to pregabalin or mirogabalin (OR: 2.40, 95% CI: 1.51 to 3.21).^37, 38^ Dose titration has been taken in account in the guideline, as the key recommendations table suggests that systemic drugs should be initiated at low doses and titrated slowly.^6^ Regarding combination pharmacotherapy, the guideline concluded that there was insufficient evidence to recommend any combination. This included one systematic review and meta-analysis which found no difference in efficacy or safety between combinations and single medications.^49^ In our review, one study (n=7,566) found that people with combination pharmacotherapy were more likely to discontinue compared to people with monopharmacotherapy.^50^

In addition to the key predictors identified for NeuP medication discontinuation, we noted that in most studies the majority of people discontinued their treatment within half a year regardless of having predictors for longer treatment duration. None of the included studies investigated potential predictors for the minority that continued treatment for a long time. Yang and colleagues (2015)^13^ and Dworkin and colleagues (2012)^12^ found that only a small proportion of people who discontinued their initial treatment switched to another one. It is possible that NeuP pharmacotherapy is discontinued due to the resolution of pain, for example in people with PHN,^51^ but this is unlikely to be the case for people with diabetic neuropathic pain, whose pain is highly likely to persist and exacerbate.^52^

### Limitations

It should be noted that we investigated associations, meaning that the predictors identified in our review do not provide information about causality. Although some studies provided results adjusted for many confounding variables, unidentified confounding variables may be present in these results.

Although this review identified a good number of studies, investigating many different potential predictors, the overlap of potential predictors investigated between the studies was limited. Thus, the conclusions about each potential predictor were generally based on a small number of studies. Only two eligible studies investigated the clinical effectiveness of NeuP medications and their adverse effects as potential predictors of discontinuation. It is interesting that our review did not identify any other studies specifically investigating adverse events or clinical effectiveness as potential predictors of discontinuation although they are key aspects of successful drug development as well as pharmacoepidemiology.

Studies that investigated potential predictors of NeuP medication prescribing often did not report their analysis such that the participants were exclusively prescribed the medication that was analysed and no other pain medications. For example, if potential predictors of being prescribed recommended medication were analysed, it was not reported whether these people received other pain medications. Analysing the predictors of prescribing this way provides useful binary categorisation between people with recommended medication and people without recommended medication. However, this simple categorisation misses other treatments which could be informative about real life prescribing. In future studies, it would be worth exploring whether there are differences between people who exclusively receive recommended medications and people with recommended medication and additional pain medications.

Although the diagnosis of interest in this review was NeuP, most of the included studies were not able to differentiate between painful and painless neuropathy based on the information provided in the databases they investigated. Studies often defined their sample using International Classification of Diseases (ICD) 9 or 10 codes and pain medication claims, but it was only in ICD-11 that a specific code for neuropathic pain was developed.^53^ The lack of information about the presence and level of pain in the included studies creates a major limitation in identifying NeuP pharmacotherapy predictors in those with the most need for treatment. Future studies could be improved by seeking additional data sources that allow classifying the sample into mild, moderate, and severe pain categories.

Another limitation in our work is that our analysis focused on finding an overall association for each potential predictor and thus we did not seek associations within the potential predictors. It is possible that some predictors are context-specific (e.g. specific to a type of NeuP diagnosis or medication) and more detailed conclusions may arise later with more studies. Analysing within-predictor associations was not considered worthwhile in this review due to the small number of studies per each potential predictor.

We made a pragmatic decision to exclude RCTs that are primarily designed to assess the efficacy of NeuP medications from our review. The medications given in these trials could be compared as the potential predictors for discontinuation. However, as our review aims to summarise all the potential predictors for NeuP medication prescribing, adherence, and discontinuation; including these trials in our review would focus disproportionately on one type of predictor (>200 RCTs ^7^). We do not believe that the inclusion of this group of studies would materially change the conclusions derived from our review.

### Conclusion

Our review has identified useful predictors of pain medication prescribing, adherence, and discontinuation in adults with NeuP. The results show that there is a need to focus on improving treatment allocation for people without diabetes and to non-white populations. Adherence may be improved by using dose titration and/or a medicine reminder. Monopharmacotherapy rather than combination pharmacotherapy for NeuP may result in better treatment persistence. These, and the other factors we identified should be considered in the development of future NeuP pharmacotherapy guidelines and in policy making. Future research should investigate how treatment effectiveness and adverse effects are associated with adherence and persistence.

## Data Availability

All data produced in the present work are contained in the manuscript.

## Details of authors’ contributions

Study conception and design: MK, LC, BS, HH

Acquisition of data: MK, DS

Analysis and interpretation of data: MK, DS, BS, HH, LC

Drafting of final manuscript: MK

Revision and critical appraisal of the manuscript: MK, BS, HH, DS, LC

Final approval of the submitted manuscript: MK, BS, HH, DS, LC

## Acknowledgments

We would like to acknowledge Mr Scott McGregor (University of Dundee Library Academic and Cultural Services) for his assistance in developing the search strategy.

## References

1. Bouhassira D, Attal N, Alchaar H, Boureau F, Brochet B, Bruxelle J, et al. Comparison of pain syndromes associated with nervous or somatic lesions and development of a new neuropathic pain diagnostic questionnaire (DN4). Pain 2005; 114: 29–36

2. Treede R-D, Jensen TS, Campbell JN, Cruccu G, Dostrovsky JO, Griffin JW, et al. Neuropathic pain: redefinition and a grading system for clinical and research purposes. Neurology 2008; 70: 1630–5

3. Baskozos G, Hébert HL, Pascal MM, Themistocleous AC, Macfarlane GJ, Wynick D, et al. Epidemiology of neuropathic pain: an analysis of prevalence and associated factors in UK Biobank. Pain Rep 2023; 8: e1066

4. van Hecke O, Austin SK, Khan RA, Smith BH, Torrance N. Neuropathic pain in the general population: A systematic review of epidemiological studies. PAIN 2014; 155: 654

5. Langley PC, Van Litsenburg C, Cappelleri JC, Carroll D. The burden associated with neuropathic pain in Western Europe. J Med Econ 2013; 16: 85–95

6. Soliman N, Moisset X, Ferraro MC, Andrade DC de, Baron R, Belton J, et al. Pharmacotherapy and non-invasive neuromodulation for neuropathic pain: a systematic review and meta-analysis. Lancet Neurol Elsevier; 2025; 24: 413–28

7. Finnerup NB, Attal N, Haroutounian S, McNicol E, Baron R, Dworkin RH, et al. Pharmacotherapy for neuropathic pain in adults: a systematic review and meta-analysis. Lancet Neurol 2015; 14: 162–73

8. Callaghan BC, Reynolds E, Banerjee M, Kerber KA, Skolarus LE, Burke JF. Longitudinal pattern of pain medication utilization in peripheral neuropathy patients. Pain 2019; 160: 592–9

9. Reynolds EL, Burke JF, Banerjee M, Kerber KA, Skolarus LE, Magliocco B, et al. Association of out-of-pocket costs on adherence to common neurologic medications. Neurology 2020; 94

10. Burnier M. Is There a Threshold for Medication Adherence? Lessons Learnt From Electronic Monitoring of Drug Adherence. Front Pharmacol Frontiers; 2019; 9

11. Gustavsson A, Bjorkman J, Ljungcrantz C, Rhodin A, Rivano-Fischer M, Sjolund K-F, et al. Pharmacological Treatment Patterns in Neuropathic Pain—Lessons from Swedish Administrative Registries. Pain Med 2013; 14: 1072–80

12. Dworkin RH, Panarites CJ, Armstrong EP, Malone DC, Pham SV. Is treatment of postherpetic neuralgia in the community consistent with evidence-based recommendations? Pain 2012; 153: 869–75

13. Yang M, Qian C, Liu Y. Suboptimal Treatment of Diabetic Peripheral Neuropathic Pain in the United States. Pain Med 2015; 16: 2075–83

14. Page MJ, Moher D, Bossuyt PM, Boutron I, Hoffmann TC, Mulrow CD, et al. PRISMA 2020 explanation and elaboration: updated guidance and exemplars for reporting systematic reviews. BMJ British Medical Journal Publishing Group; 2021; 372: n160

15. Hall GC, Carroll D, McQuay HJ. Primary care incidence and treatment of four neuropathic pain conditions: A descriptive study, 2002–2005. BMC Fam Pract 2008; 9: 26

16. Hall GC, Morant SV, Carroll D, Gabriel ZL, McQuay HJ. An observational descriptive study of the epidemiology and treatment of neuropathic pain in a UK general population. BMC Fam Pract 2013; 14: 28

17. Reed C, Hong J, Novick D, Lenox-Smith A, Happich M. Incidence of diabetic peripheral neuropathic pain in primary care –; a retrospective cohort study using the United Kingdom General Practice Research Database. Pragmatic Obs Res Dove Press; 2013; 4: 27–37

18. Sicras-Mainar A, Rejas J, Navarro-Artieda R. Comparative effectiveness and costs of generic and brand-name gabapentin and venlafaxine in patients with neuropathic pain or generalized anxiety disorder in Spain. Clin Outcomes Res 2015; 299

19. Higgins JPT, Morgan RL, Rooney AA, Taylor KW, Thayer KA, Silva RA, et al. A tool to assess risk of bias in non-randomized follow-up studies of exposure effects (ROBINS-E). Environ Int 2024; 186: 108602

20. Sterne JAC, Savović J, Page MJ, Elbers RG, Blencowe NS, Boutron I, et al. RoB 2: a revised tool for assessing risk of bias in randomised trials. BMJ British Medical Journal Publishing Group; 2019; 366: l4898

21. Shea BJ, Reeves BC, Wells G, Thuku M, Hamel C, Moran J, et al. AMSTAR 2: a critical appraisal tool for systematic reviews that include randomised or non-randomised studies of healthcare interventions, or both. BMJ British Medical Journal Publishing Group; 2017; 358: j4008

22. Reynolds EL, Burke JF, Banerjee M, Callaghan BC. Randomized controlled trial of a clinical decision support system for painful polyneuropathy. Muscle Nerve 2020; 61: 640–4

23. Shaparin N, Slattum PW, Bucior I, Nalamachu S. Relationships Among Adverse Events, Disease Characteristics, and Demographics in Treatment of Postherpetic Neuralgia With Gastroretentive Gabapentin. Clin J Pain 2015; 31: 983–91

24. Lakkad M, Martin B, Li C, Harrington S, Dayer L, Painter JT. Factors Associated With Guideline-Concordant Pharmacological Treatment for Neuropathic Pain Among Breast Cancer Survivors. Clin Breast Cancer 2023; 23: 598–619

25. Boulanger L, Zhao Y, Foster TS, Fraser K, Bledsoe SL, Russell MW. Impact of comorbid depression or anxiety on patterns of treatment and economic outcomes among patients with diabetic peripheral neuropathic pain. Curr Med Res Opin 2009; 25: 1763–73

26. Chahine B, Al Souheil F. Dispensing patterns of drugs used for neuropathic pain and adherence to NeuPSIG guideline: an observational study. Egypt J Neurol Psychiatry Neurosurg 2021; 57: 142

27. Udall M, Kudel I, Cappelleri JC, Sadosky A, King-Concialdi K, Parsons B, et al. Epidemiology of physician-diagnosed neuropathic pain in Brazil. J Pain Res 2019; Volume 12: 243–53

28. Butler S, Eek D, Ring L, Gordon A, Karlsten R. The utility/futility of medications for neuropathic pain – an observational study. Scand J Pain De Gruyter; 2019; 19: 327–35

29. Margolis J, Princic N, Smith D, Abraham L, Cappelleri J, Shah S, et al. Development of a novel algorithm to determine adherence to chronic pain treatment guidelines using administrative claims. J Pain Res 2017; Volume 10: 327–39

30. Sadosky A, Schaefer C, Mann R, Bergstrom F, Baik R, Parsons B, et al. Burden of illness associated with painful diabetic peripheral neuropathy among adults seeking treatment in the US: results from a retrospective chart review and cross-sectional survey. Diabetes Metab Syndr Obes Targets Ther 2013; 79

31. Pillay P, Wadley AL, Cherry CL, Karstaedt AS, Kamerman PR. Pharmacological treatment of painful HIV-associated sensory neuropathy. S Afr Med J 2015; 105: 769–72

32. Mbrah AK, Nunes AP, Hume AL, Zhao D, Jesdale BM, Bova C, et al. Prevalence and treatment of neuropathic pain diagnoses among U.S. nursing home residents. Pain 2022; 163: 1370–7

33. Gharibian D, Polzin JK, Rho JP. Compliance and Persistence of Antidepressants Versus Anticonvulsants in Patients With Neuropathic Pain During the First Year of Therapy. Clin J Pain 2013; 29: 377–81

34. Oladapo AO, Barner JC, Rascati KL, Strassels SA. A Retrospective Database Analysis of Neuropathic Pain and Oral Antidiabetic Medication Use and Adherence Among Texas Adults With Type 2 Diabetes Enrolled in Medicaid. Clin Ther 2012; 34: 605–13

35. Zhao Y, Sun P, Watson P. Medication adherence and healthcare costs among patients with diabetic peripheral neuropathic pain initiating duloxetine versus pregabalin. Curr Med Res Opin 2011; 27: 785– 92

36. Giannopoulos S, Kosmidou M, Sarmas I, Markoula S, Pelidou S-H, Lagos G, et al. Patient Compliance With SSRIs and Gabapentin in Painful Diabetic Neuropathy. Clin J Pain 2007; 23: 267–9

37. Kato K, Kodama S, Shiosakai K, Kimura T. Relationship between the dose titration and adherence of mirogabalin in patients with peripheral neuropathic pain depending on renal function: a nationwide electronic medical record database study. Expert Opin Pharmacother 2023; 24: 267–82

38. Yeh Y-C, Cappelleri JC, Marston XL, Shelbaya A. Effects of dose titration on adherence and treatment duration of pregabalin among patients with neuropathic pain: A MarketScan database study. Mallhi TH, editor. PLOS ONE 2021; 16: e0242467

39. Wu N, Chen S, Boulanger L, Fraser K, Bledsoe SL, Zhao Y. Duloxetine compliance and its association with healthcare costs among patients with diabetic peripheral neuropathic pain. J Med Econ 2009; 12: 192–202

40. Chen S-Y, Wu N, Boulanger L, Fraser KA, Zhao Y. The Relationship Between Average Daily Dose, Medication Adherence, and Health-Care Costs among Diabetic Peripheral Neuropathic Pain Patients Initiated on Duloxetine Therapy: Duloxetine Average Daily Dose, Adherence and Costs. Pain Pract 2010; 10: 530–9

41. Sicras-Mainar A, Rejas-Gutiérrez J, Pérez-Paramo M, Sánchez-Alvarez L, Navarro-Artieda R, Darbà J. Consequences on economic outcomes of generic versus brand-name drugs used in routine clinical practice: the case of treating peripheral neuropathic pain or generalized anxiety disorder with pregabalin. Expert Rev Pharmacoecon Outcomes Res 2019; 19: 45–57

42. Gore M, Dukes E, Rowbotham DJ, Tai K, Leslie D. Clinical characteristics and pain management among patients with painful peripheral neuropathic disorders in general practice settings. Eur J Pain 2007; 11: 652–64

43. Hall GC, Carroll D, Parry D, McQuay HJ. Epidemiology and treatment of neuropathic pain: The UK primary care perspective. Pain 2006; 122: 156–62

44. Wang Y-F, Chen Y-T, Tsai C-W, Yen Y-C, Chen Y-C, Shia B-C, et al. Persistence of pregabalin treatment in Taiwan: a nation-wide population-based study. J Headache Pain 2020; 21: 54

45. Lehmann HC, Wunderlich G, Fink GR, Sommer C. Diagnosis of peripheral neuropathy. Neurol Res Pract 2020; 2: 20

46. Morgan S, Hanley G, Cunningham C, Quan Q. Ethnic differences in the use of prescription drugs: a cross-sectional analysis of linked survey and administrative data. Open Med Peer-Rev Indep Open-Access J Open Med; 2011; 5

47. Fiscella K, Franks P, Doescher MP, Saver BG. Disparities in Health Care by Race, Ethnicity, and Language Among the Insured: Findings From a National Sample. Med Care 2002; 40: 52

48. Mayberry RM, Mili F, Ofili E. Racial and ethnic differences in access to medical care. Med Care Res Rev MCRR Med Care Res Rev; 2000; 57 Suppl 1

49. Balanaser M, Carley M, Baron R, Finnerup N, Moore R, Rowbotham M, et al. Combination pharmacotherapy for the treatment of neuropathic pain in adults: systematic review and meta-analysis. Pain 2023; 164

50. Kuo K-L, Brixner D, Lipman AG, Goodman M, Hung M, Oderda GM. Single-Versus Multiple-Drug Pharmacotherapy in the Management of Diabetic Painful Neuropathy. J Pain Palliat Care Pharmacother 2016; 30: 184–94

51. Reda H, Greene K, Rice FL, Rowbotham MC, Petersen KL. Natural history of herpes zoster: Late follow-up of 3.9 years (n = 43) and 7.7 years (n = 10). PAIN® 2013; 154: 2227–33

52. Boulton AJM, Armstrong WD, Scarpello JHB, Ward JD. The natural history of painful diabetic neuropathy—a 4-year study. Postgrad Med J 1983; 59: 556–9

53. Scholz J, Finnerup NB, Attal N, Aziz Q, Baron R, Bennett MI, et al. The IASP classification of chronic pain for ICD-11: chronic neuropathic pain. Pain 2019; 160: 53–9

54. Adhikari S, Bista D, Shrestha R, Woods D. Prescribing Patterns and Off-Label Use of Gabapentinoid Agents at Dhulikhel Hospital, Nepal: A Cross-Sectional Study. J Pain Res 2024; 17: 4377–91

55. Aggarwal P, Mishra PN, Mathur VN, Velivela KC, Khan S, Deshmukh P, et al. Drug Usability Survey (DUS) of Gabapentinoid and Its Combinations Among Indian Patients With Neuropathic Pain: Results From a Real-World, Multicenter, Retrospective Survey at Neurology Clinics. Cureus Cureus; 2025; 17

56. Anderson SG, Malipatil NS, Roberts H, Dunn G, Heald AH. Socioeconomic deprivation independently predicts symptomatic painful diabetic neuropathy in type 1 diabetes. Prim Care Diabetes 2014; 8: 65– 9

57. Anderson SG, Narayanan RP, Dunn G, Heald AH. Socioeconomic deprivation independently predicts symptomatic painful diabetic neuropathy in people with type 2 diabetes. 2016; 20

58. Banks C, A. Bowman L, Merrey J, Waldfogel JM. Characterization of Outpatient Gabapentinoid Prescribing for Pain. J Pain Palliat Care Pharmacother 2023; 37: 143–7

59. Berger A, Dukes E, McCarberg B, Liss M, Oster G. Change in opioid use after the initiation of gabapentin therapy in patients with postherpetic neuralgia. Clin Ther 2003; 25: 2809–21

60. Chen S, Wu N, Fraser K, Boulanger L, Zhao Y. Opioid use and healthcare costs among patients with DPNP initiating duloxetine versus other treatments. Curr Med Res Opin 2010; 26: 2507–16

61. Chen S, Wu N, Boulanger L, Fraser K, Zhao Z, Zhao Y. Factors associated with pain medication selection among patients diagnosed with diabetic peripheral neuropathic pain: a retrospective study. J Med Econ 2011; 14: 411–20

62. Dieleman JP, Kerklaan J, Huygen FJPM, Bouma PAD, Sturkenboom MCJM. Incidence rates and treatment of neuropathic pain conditions in the general population IZ. Pain 2008; 137: 681–8

63. Dinesh Babu U, Pradeep Kumar B. T, Gomathi G. Drug utilization patterns for peripheral neuropathy in a neurology outpatient department at a tertiary care center: A cross-sectional observational study. Natl J Physiol Pharm Pharmacol 2024; 14: 940–940

64. Dragic L, Webb T, Chandler M, Harrington SB, McDade E, Dayer L, et al. Comparing Effectiveness of Gabapentin and Pregabalin in Treatment of Neuropathic Pain: A Retrospective Cohort of Palliative Care Outpatients. J Pain Palliat Care Pharmacother 2020; 34: 192–6

65. Gewandter JS, Kleckner AS, Marshall JH, Brown JS, Curtis LH, Bautista J, et al. Chemotherapy-induced peripheral neuropathy (CIPN) and its treatment: an NIH Collaboratory study of claims data. Support Care Cancer Off J Multinatl Assoc Support Care Cancer 2020; 28: 2553–62

66. Gore M, Sadosky A, Tai K-S, Stacey B. A Retrospective Evaluation of the Use of Gabapentin and Pregabalin in Patients with Postherpetic Neuralgia in Usual-Care Settings. Clin Ther 2007; 29: 1655–70

67. Gore M, Zlateva G, Tai K, Chandran AB, Leslie D. Retrospective Evaluation of Clinical Characteristics, Pharmacotherapy and Healthcare Resource Use among Patients Prescribed Pregabalin or Duloxetine for Diabetic Peripheral Neuropathy in Usual Care. Pain Pract 2011; 11: 167–79

68. Gore M, Tai K, Zlateva G, Bala Chandran A, Leslie D. Clinical Characteristics, Pharmacotherapy, and Healthcare Resource Use among Patients with Diabetic Neuropathy Newly Prescribed Pregabalin or Gabapentin. Pain Pract 2011; 11: 528–39

69. Goswami S, Ramachandran S, Sharma M, Barnard M. Gabapentin and Opioids Utilization in Patients With Diabetic Neuropathy Enrolled in Medicare (2012–2016): A Cohort Study. J Pharm Pract 2023; 36: 1085–94

70. Han G, Han Y, Yu L, Zhao Y, Yu Z. Patterns and Trends in Pharmacological Treatment for Outpatients with Postherpetic Neuralgia in Six Major Areas of China, 2015–2019. Healthcare Multidisciplinary Digital Publishing Institute; 2023; 11: 764

71. Jacob L, Kaiser M, Kostev K. Incidence of antiepileptic drug therapy and factors associated with their prescribing in outpatients with diabetic polyneuropathy. Prim Care Diabetes 2021; 15: 535–40

72. Jha KK, Zaman ZA. An Observational Study on Effectiveness and Safety of Amitriptyline, Duloxetine, and Pregabalin in Painful Diabetic Neuropathy. IJPCR 2024; 16

73. Johnson P, Becker L, Halpern R, Sweeney M. Real-World Treatment of Post-herpetic Neuralgia with Gabapentin or Pregabalin. Clin Drug Investig 2013; 33: 35–44

74. Johnston SS, Udall M, Alvir J, McMorrow D, Fowler R, Mullins D. Characteristics, Treatment, and Health Care Expenditures of Medicare Supplemental-Insured Patients with Painful Diabetic Peripheral Neuropathy, Post-Herpetic Neuralgia, or Fibromyalgia. Pain Med 2014; 15: 562–76

75. Knoerl R, Sohn MB, Foust M, Francar L, O’Rourke MA, Morrow GM, et al. Exploring Analgesic Use Patterns Among Cancer Survivors With Chronic Chemotherapy-Induced Peripheral Neuropathy. Oncol Nurs Forum 2024; 51: 445–50

76. Koopman JSHA, Huygen Frank J, Dieleman JP, De Mos M, Sturkenboom MCJM. Pharmacological Treatment of Neuropathic Facial Pain in the Dutch General Population. J Pain 2010; 11: 264–72

77. Lin CC, Callaghan BC, Burke JF, Kerber KA, Bicket MC, Esper GJ, et al. Prescription Opioid Initiation for Neuropathy, Headache, and Low Back Pain: A US Population-based Medicare Study. J Pain 2023; 24: 2268–82

78. Marcianò G, Siniscalchi A, Di Gennaro G, Rania V, Vocca C, Palleria C, et al. Assessing Gender Differences in Neuropathic Pain Management: Findings from a Real-Life Clinical Cross-Sectional Observational Study. J Clin Med Multidisciplinary Digital Publishing Institute; 2024; 13: 5682

79. Mittal M, Pasnoor M, Mummaneni RB, Khan S, McVey A, Saperstein D, et al. Retrospective Chart Review of Duloxetine and Pregabalin in the Treatment of Painful Neuropathy. Int J Neurosci 2011; 121: 521–7

80. Muñoz-Vendrell A, Valín-Villanueva P, Tena-Cucala R, Campoy S, Martínez-Yélamos S, Huerta-Villanueva M. Second-line pharmacological treatment strategies for trigeminal neuralgia: A retrospective comparison of lacosamide, gabapentin and baclofen. Headache 2025;

81. Nyqvist L, Åkerstedt J, Thoreson O. Current trends in the medical treatment of neuropathic low back pain: a Swedish registry-based study of 1.7 million people. BMC Musculoskelet Disord 2024; 25: 486

82. Patil PR, Wolfe J, Said Q, Thomas J, Martin BC. Opioid Use in the Management of Diabetic Peripheral Neuropathy (DPN) in a Large Commercially Insured Population. Clin J Pain 2015; 31: 414–24

83. Pérez C, Navarro A, Saldaña MT, Masramón X, Pérez M, Rejas J. Clinical and Resource Utilization Patterns in Patients with Refractory Neuropathic Pain Prescribed Pregabalin for the First Time in Routine Medical Practice in Primary Care Settings in Spain. Pain Med 2013; 14: 1954–63

84. Sanchez RJ, Mardekian J, Clair AG, Cappelleri JC. Therapeutic and Subtherapeutic Dosing of Pregabalin: Medication Adherence, Healthcare Resource Utilization, and Costs. Peripher Neuropathy

85. Sutema IAMP, Jaya MKA, Bakta IM. Medicine reminder to improve treatment compliance on geriatric patients with diabetic neuropathy at Sanglah Central Hospital, Bali-Indonesia. Bali Med J 2018; 7: 516

86. Toth C, Brady S, Hatfield M. The importance of catastrophizing for successful pharmacological treatment of peripheral neuropathic pain. J Pain Res 2014; 327

87. Winterbottom LM, Fong AM, Benkstein KL, Liang B, Snodgrass LS, Parks-Huitron H. Impact of a Clinical Pharmacy Consult Service on Guideline Adherence and Management of Gabapentin for Neuropathic Pain. J Manag Care Pharm 2006; 12: 61–9

88. Wu N, Chen S, Hallett LA, Boulanger L, Fraser KA, Patel CK, et al. Opioid Utilization and Health-Care Costs among Patients with Diabetic Peripheral Neuropathic Pain Treated with Duloxetine vs. Other Therapies. Pain Pract 2011; 11: 48–56

89. Zhao Y, Liu J, Zhao Y, Thethi T, Fonseca V, Shi L. Predictors of Duloxetine versus Other Treatments among Veterans with Diabetic Peripheral Neuropathic Pain: A Retrospective Study. Pain Pract 2012; 12: 366–73

