## Supplemental materials for "Identifying predictors of neuropathic pain medication prescribing, adherence, and discontinuation: a systematic review and meta-analysis"

Table S1. Search strategy. …………………………………………………………………………………………………………………………………………………………………………………… 2

Table S2. Neuropathic pain diagnosis in the included studies. …………………………………………………………………………………………………………………………………. 4

Table S3. Summary of medication included and excluded. …………………………………………………………………………………………………………………………………….. 11

Table S4. Medications included and excluded from our analysis for each included study. ……………………………………………………………………………………………16

Table S5. Medications considered recommended in the included studies. ……………………………………………………………………………………………………………….. 30

Table S6. Studies excluded in full-text screening. …………………………………………………………………………………………………………………………………………………. 33

Table S7. Subcategories of prescribing studies. ……………………………………………………………………………………………………………………………………………………. 42

Table S13. Analysis of predictors of being prescribed recommended medication as the first choice. ……………………………………………………………………………. 50

Table S14. Analysis of predictors of being prescribed recommended medication at any point. ……………………………………………………………………………………. 53

Table S15. Analysis of predictors of being prescribed opioids as the first choice. ………………………………………………………………………………………………………. 60

Table S16. Analysis of predictors of being prescribed opioids at any point. ……………………………………………………………………………………………………………….. 62

Table S17. Analysis of predictors of being prescribed non-opioid analgesics. ……………………………………………………………………………………………………………. 71

Table S18. Analysis of predictors of being prescribed any pain medication. ………………………………………………………………………………………………………………. 75

Table S19. Analysis of predictors of adherence. ……………………………………………………………………………………………………………………………………………………. 79

Table S20. Analysis of predictors of discontinuation. …………………………………………………………………………………………………………………………………………….. 89

Figure S1. Meta-analysis: discontinuation between different medications based on Gore et al. (2007a) data. ……………………………………………………………….. 96

Figure S2. Meta-analysis: discontinuation between different medications based on Hall et al. (2008) data. …………………………………………………………………… 97

Figure S3. Meta-analysis: discontinuation between mixed pain and pure neuropathic pain based on Gore et al. (2007a) data. …………………………………………. 98

Table S21. Summary of all results. ……………………………………………………………………………………………………………………………………………………………………… 99

Table S1. Search strategy. The databases were searched from inception to 1st of August 2025.

| **#** | **PubMed** | **Embase via Ovid** | **Web of Science via Clarivate** | **CINAHL via EBSCO** |
| --- | --- | --- | --- | --- |
| **1** | “neuropath*”[tw] or “neuralgia”[tw] or “nerve pain”[tw] or "neuralgia"[mesh] | neuropathic pain/ or neuropathic.mp. or neuralgia/ or neuralgia.mp. or nerve pain.mp. | TS=(“neuropath*” or “neuralgia” or “nerve pain”) | (MH "Neuralgia") or "neuralgia" or “neuropath*” or “nerve pain” |
| **2** | “pharmacological treatment”[tw] or “pharmacotherapy”[tw] or “pharmacological therapy”[tw] or “drug therapy”[tw] or “drug treatment”[tw] or “drug intervention”[tw] or “pharmacological intervention”[tw] or “medicine”[tw] or “medication”[tw] or “analgesic*”[tw] or “gabapentinoid*”[tw] or “pregabalin”[tw] or “gabapentin”[tw] or “tricyclic antidepressant*”[tw] or “amitriptyline”[tw] or “imipramine”[tw] or “nortriptyline”[tw] or “clomipramine”[tw] or “desipramine”[tw] or “maprotiline”[tw] or “serotonin-noradrenaline reuptake inhibitor*”[tw] or “SRNI”[tw] or “duloxetine”[tw] or “venlafaxine”[tw] or “desvenlafaxine”[tw] or“anticonvulsant*”[tw] or “sodium channel blocker*”[tw] or “carbamazepine” or “tramadol”[tw] or “lidocaine”[tw] or “capsaicin”[tw] or “botulinum”[tw] or “botox”[tw] or “opioid*”[tw] or “opiate*”[tw] or “morphine”[tw] or “diamorphine”[tw] or “hydromorphone”[tw] or “oxycodone”[tw] or “fentanyl”[tw] or “buprenorphine”[tw] or “methadone”[tw] or “tapentadol”[tw] or “pethidine”[tw] or “pentazocine”[tw] or "pain management"[mesh] or "pain/drug therapy"[mesh] | pharmacological treatment.mp. or drug therapy/ or pharmacotherapy.mp. or pharmacological therapy.mp. or drug therapy.mp. or drug treatment.mp. or drug intervention.mp. or pharmacological intervention.mp. or medicine.mp. or medication.mp. or analgesic.mp. or analgesic agent/ or pregabalin/ or gabapentin/ or gabapentinoid*.mp. or amitriptyline.mp. or amitriptyline/ or tricyclic antidepressant agent/ or tricyclic antidepressant*.mp. or imipramine.mp. or imipramine/ or venlafaxine/ or venlafaxine.mp. or desipramine/ or desipramine.mp. or nortriptyline.mp. or nortriptyline/ or clomipramine/ or clomipramine.mp. or maprotiline/ or maprotiline.mp. or serotonin noradrenalin reuptake inhibitor/ or serotonin-noradrenaline reuptake inhibitor*.mp. or SNRI*.mp. or duloxetine/ or duloxetine.mp. or desvenlafaxine/ or desvenlafaxine.mp. or anticonvulsant*.mp. or anticonvulsive agent/ or sodium channel blocker*.mp. or sodium channel blocking agent/ or carbamazepine/ or carbamazepine.mp. or tramadol.mp. or tramadol/ or lidocaine/ or lidocaine.mp. or capsaicin/ or capsaicin.mp. or botulinum.mp. or botulinum toxin/ or botulinum toxin A/ or botox.mp. or opioid*.mp. or opiate*.mp. or opiate/ or morphine/ or morphine.mp. or diamorphine/ or diamorphine.mp. or hydromorphone/ or hydromorphone.mp. or oxycodone/ or oxycodone.mp. or fentanyl/ or fentanyl.mp. or buprenorphine/ or buprenorphine.mp. or methadone/ or methadone.mp. or tapentadol/ or tapentadol.mp. or pethidine/ or pethidine.mp. or pentazocine/ or pentazocine.mp. or pain management.mp. | TS=(“pharmacological treatment” or “pharmacotherapy” or “pharmacological therapy” or “drug therapy” or “drug treatment” or “drug intervention” or “pharmacological intervention” or “medicine” or “medication” or “analgesic*” or “gabapentinoid*” or “pregabalin” or “gabapentin” or “tricyclic antidepressant*” or “amitriptyline” or “imipramine” or “nortriptyline” or “clomipramine” or “desipramine” or “maprotiline” or “serotonin-noradrenaline reuptake inhibitor*” or “SRNI” or “duloxetine” or “venlafaxine” or “desvenlafaxine” or“anticonvulsant*” or “sodium channel blocker*” or “carbamazepine” or “tramadol” or “lidocaine” or “capsaicin” or “botulinum” or “botox” or “opioid*” or “opiate*” or “morphine” or “diamorphine” or “hydromorphone” or “oxycodone” or “fentanyl” or “buprenorphine” or “methadone” or “tapentadol” or “pethidine” or “pentazocine” or "pain management") | “pharmacological treatment” or “pharmacotherapy” or “pharmacological therapy” or “drug therapy” or “drug treatment” or “drug intervention” or “pharmacological intervention” or “medicine” or “medication” or “analgesic*” or “gabapentinoid*” or “pregabalin” or “gabapentin” or “tricyclic antidepressant*” or “amitriptyline” or “imipramine” or “nortriptyline” or “clomipramine” or “desipramine” or “maprotiline” or “serotonin-noradrenaline reuptake inhibitor*” or “SRNI” or “duloxetine” or “venlafaxine” or “desvenlafaxine” or“anticonvulsant*” or “sodium channel blocker*” or “carbamazepine” or “tramadol” or “lidocaine” or “capsaicin” or “botulinum” or “botox” or “opioid*” or “opiate*” or “morphine” or “diamorphine” or “hydromorphone” or “oxycodone” or “fentanyl” or “buprenorphine” or “methadone” or “tapentadol” or “pethidine” or “pentazocine” or "pain management" or (MH “pain management”) |
| **3** | “prescrib*”[tw] or “prescription*”[tw] or “adherence”[tw] or “nonadherence”[tw] or “non-adherence”[tw] or “noncompliance”[tw] or “medication persistence”[tw] or “compliance”[tw] or “non-compliance”[tw] or “discontinuation”[tw] or "medication adherence"[mesh] or “patient compliance”[mesh] or “prescriptions”[mesh] | prescrib*.mp.or prescription*.mp. or adherence.mp.or nonadherence.mp. or non-adherence.mp. or noncompliance.mp. or medication persistence.mp. or compliance.mp. or non-compliance.mp. or discontinuation.mp. or medication adherence.mp. or medication compliance/ or patient compliance/ | TS=(“prescrib*” or “prescription*” or “adherence” or “nonadherence” or “non-adherence” or “noncompliance” or “medication persistence” or “compliance” or “non-compliance” or “discontinuation” or "medication adherence") | “prescrib*” or “prescription*” or “adherence” or “nonadherence” or “non-adherence” or “noncompliance” or “medication persistence” or “compliance” or “non-compliance” or “discontinuation” or "medication adherence" or (MH “patient compliance”) |
| **4** | (#1 AND #2 AND #3) NOT ("animals"[mesh] NOT "humans"[mesh]) AND English language filter | (#1 AND #2 AND #3) AND Limit to humans AND English language filter | (#1 AND #2 AND #3) AND English language filter | (#1 AND #2 AND #3) AND human filter AND English language filter |

Table S2. Neuropathic pain diagnosis in the included studies. CIPN: chemotherapy-induced peripheral neuropathy, CTS: carpal tunnel syndrome, DN: diabetic neuropathy, HIV-SN: human immunodeficiency virus sensory neuropathy, NeuP: neuropathic pain, NR: not reported, PHN: postherpetic neuralgia, PLP: phantom limb pain, PSN: post-surgical neuropathic pain, PTN: post-traumatic neuropathic pain, TN: trigeminal neuralgia.

| Author & Year | Participant or data source | NeuP diagnosis types | NeuP diagnosis method |
| --- | --- | --- | --- |
| Adhikari et al. (2024) | Recruited from an outpatient pharmacy of Dhulikhel Hospital (Nepal). | Peripheral NeuP  PHN  DN | NR |
| Aggarwal et al. (2025) | Identified from patients visiting the neurology clinic. Data from the medical records of various primary healthcare clinics across India. | Any NeuP | “patients presenting with neuropathic pain…visiting the neurology clinic” |
| Anderson et al. (2014) | Type 1 diabetics attending GP practices in Central and Eastern Cheshire identified through a centralised data facility afforded by EMIS (the majority GP systems provider in this part of the UK) | DN | Symptomatic neuropathic pain of a degree to require pharmacological treatment was present in 280 patients of which 67 had a formal diagnosis of peripheral neuropathy recorded. |
| Anderson et al. (2015) | Identified from pseudo-anonymised records of people attending general practices in the catchment area of Central and Eastern Cheshire Primary Care Trust | DN | Participants (relevant to our topic) were identified on the basis of having "symptomatic neuropathic pain of a degree requiring pharmacological treatment". Only some of these participants "had a formal diagnosis".  No description in methods what this means. Could be just diabetic with NeuP medications. |
| Banks et al. (2023) | Identified from outpatients attending internal medicine clinics in an academic medical centre | DN | NR |
| Berger et al. (2003) | Identified from Protocare Sciences Managed Care Database | PHN | ICD-9-CM code 53.1X |
| Billig et al. (2022) | Identified from The Clinformatics DataMart  databases | CTS | ICD-9: 354.0  ICD-10: G56.0, G56.00, G56.01, G56.02, G56.03 |
| Boulanger et al. (2009) | Identified from Thomson's Medstat MarketScan Commercial Claims database and Medicare Supplemental database | DN | ICD-9-CM: 250.6x or 357.2x |
| Butler et al. (2018) | Identified from RELIEF study participants | DN  PTN | DN: (1) medical history of diabetes mellitus type 1 or 2 (2) pain with a distal symmetric distribution, arterial occlusion excluded, of at least 3 months’ duration (3) diagnosed sensory disturbance with a distal symmetric distribution of at least 3 months’ duration involving one or more of the following senses; light touch (examined with a brush [SENSELab Brush-05, Somedic]), pinprick (examined with a cocktail pin), warmth (examined with a metallic roller at 40°C [Somedic]), cold (examined with a metallic roller at 20°C [Somedic]).  PTN: (1) history of pain due to injury (accidental or surgical) to one or several well-defined peripheral nerves (2) pain localized to the area of the specific nerve(s), (3) a diagnosed sensory disturbance of the affected area of one or more of the following senses; light touch (examined with a brush [SENSELab Brush-05, Somedic]), pinprick (examined with a cocktail pin), warmth (examined with a metallic roller at 40°C [Somedic]), cold (examined with a metallic roller at 20°C [Somedic]) |
| Callaghan et al. (2019) | Identified from the Clinformatics Datamart database | Peripheral NeuP | ICD-9 codes: 356.x, 357.1–8, 357.82, 357.89 and 357.9. |
| Chahine and Al Souheil (2021) | Recruited from community pharmacies | Several | “Clinically diagnosed with a neuropathic pain disorder by an orthopaedist, neurologist, or neurosurgeon." |
| Chen et al. (2010a) | Identified from Thomson's Medstat MarketScan Commercial Claims database | DN | ICD-9-CM 250.6x and/or 357.2x |
| Chen et al. (2010b) | Identified from Thomson's Medstat MarketScan Commercial Claims database | DN | ICD-9-CM 250.6x and/or 357.2x |
| Chen et al. (2011) | Identified from Thomson's Medstat MarketScan Commercial Claims database | DN | ICD-9-CM 250.6x and/or 357.2 |
| Dieleman et al. (2008) | Identified from the Integrated Primary Care Information (IPCI) database | Mononeuropathy  CTS  DN  PHN  Facial NeuP | “Based on expert judgement.” |
| Dinesh Babu et al. (2024) | Recruited from neurology OPD S.S. Hospital, Institute of Medical Sciences,  Banaras Hindu University, Varanasi, Uttar Pradesh. | Peripheral NeuP | “Patients diagnosed with peripheral neuropathy, confirmed  through clinical evaluation, nerve conduction studies,  electromyography” |
| Dragic et al. (2020) | Identified from electronic health record data from outpatients receiving pregabalin or gabapentin by University of Arkansas for Medical Sciences (UAMS) Palliative Care Clinic providers | NR | NR |
| Dworkin et al. (2012) | Identified from the Thomson Reuters MarketScan database and Medicare (Medicare Supplementar and Coordination of Benefits) database | PHN | ICD-9 codes: 053.12, 053.13, 053.19 |
| Gewandter et al. (2020) | Identified from claims and administrative data from two national and one regional insurers participating in the NIH Collaboratory DRN (i.e., Anthem [via HealthCore], Aetna, and Harvard Pilgrim Health Care). | CIPN | Two different definitions explored: “CIPN “Definition 1” was satisfied by any of the 15 ICD-9 or ICD-10 codes listed in Appendix 1, which were based on common symptoms of CIPN and general peripheral neuropathy codes. Codes that were specifically related to other types of peripheral neuropathy (e.g., diabetic neuropathy) were not considered for the outcome or when excluding patients from the sample. CIPN “Definition 2” included only ICD-9-CM 357.6 (Polyneuropathy due to drugs) or ICD-10-CM G62.0 (Drug-induced polyneuropathy).” |
| Gharibian et al. (2013) | Identified Southern California Kaiser Permanente (large health care organisation) electronic medical and pharmacy database | NR | ICD-9 code for neuropathic pain (no specific codes reported) |
| Giannopoulos et al. (2007) | NR | DN | Type II diabetes lasting for at least 2 years, clinically relevant lower limb distal neuropathy, and minimum score of 2 on a pain intensity scale ranging of 0 to 4. |
| Gore et al. (2007a) | Identified from the General Practice Research Database | Peripheral NeuP | Conditions categorized as Pure PNDs included diabetic neuropathy (with or without pain), postherpetic neuralgia, Complex Regional Pain Syndrome II (formerly causalgia), phantom limb pain, trigeminal neuralgia, atypical facial pain, and other disorders of the peripheral nervous system associated with neuropathic pain (including carpal tunnel syndrome, lesions of peripheral nerves, inflammatory and toxic neuropathies, and nerve root and plexus disorders).  Conditions categorized as Mixed PNDs included low back pain with neuropathic involvement and other spinal pain (neck and upper back) with neuropathic involvement. |
| Gore et al. (2007b) | Identified from The PharMetrics Patient-Centric Database | PHN | ICD-9-CM code 053.1X |
| Gore et al. (2011a) | Identified from PharMetrics Patient-Centric Database | DN | ICD-9-CM 250.6x or 357.2x |
| Gore et al. (2011b) | Identified from LifeLink Health Plan (formerly PharmaMetrics) Claims Database | DN | ICD-9-CM 250.6x or 357.2x |
| Goswami et al. (2023) | Identified from the Medicare claims database | DN | ICD-9 codes: 250.6, 337.1, 337.9, 354.9, 355.8, 355.9, 356.9, 357.2.  ICD-10 codes: E11.40, E10.40, E11.65, E10.65, G99.0, G90.9, G56.90, G57.90, G58.9, G60.9, E08.42, E09.42, E10.42, E11.42, E13.42. |
| Gustavsson et al. (2013) | Identified from Western Sweden Vega register | Pure NeuP  Mixed pain | ICD-10 codes:  Neuropathy: Trigeminal neuropathies (G50), Diseases of other cerebral nerves (G52), Cerebral neuropathies in diseases classified elsewhere (G53), Nerve root and plexus diseases (G54), Nerve root and plexus compression in diseases classified elsewhere (G55), Mononeuropathies of the upper extremity (G56), Mononeuropathies of the lower extremity (G57), Other mononeuropathies (G58), Mononeuropathy in diseases classified elsewhere (G59), Hereditary and idiopathic neuropathy (G60), Polyneuritis (G61), Other polyneuropathies (G62), Polyneuropathy in diseases classified elsewhere (G63), Other diseases of the peripheral nervous system (G64), Paraplegia and tetraplegia (G82), Postprocedural disorders of nervous system, not elsewhere classified (G97), Other disorders of bone (M89), Other symptoms and signs involving the nervous and musculoskeletal systems (R29).  Mixed pain: Cervicalgia (M54.2), Sciatica (M54.3), Lumbago with sciatica (M54.4) |
| Hall et al. (2006) | Identified from the General Practice Research Database. | PHN  TN  PLP  DN | PHN: a specific term for PHN or an acute herpes zoster term plus either neuropathy, or neuropathic pain, 3–6 months after the first acute herpes zoster entry.  TN: a specific term for this diagnosis.  PLP: a specific term or a term for amputation plus either a neuropathy or neuropathic pain record 3–24 months after the first amputation code.  PDN: a specific term; a term for diabetic neuropathy with a prescription for a treatment for pain current at the date of diagnosis; a record of diabetes and neuropathic pain or record of diabetes and both neuralgia and a treatment for pain current on the date of the neuralgia code. |
| Hall et al. (2008) | Identified from Health Information Network database (THIN) | PHN  TN  PLP  DN | PHN: a specific term for PHN or an acute herpes zoster term plus either neuropathy, or neuropathic pain, 3–6 months after the first acute herpes zoster entry.  TN: a specific term for this diagnosis.  PLP: a specific term or a term for amputation plus either a neuropathy or neuropathic pain record 3–24 months after the first amputation code.  DN: a specific term; a term for diabetic neuropathy with a prescription for a treatment for pain current at the date of diagnosis; a record of diabetes and a general term for neuropathic pain or record of diabetes and both neuralgia and a treatment for pain current on the date of the neuralgia code. |
| Hall et al. (2013) | Identified from the General Practice Research Database | PHN  DN  PLP  NeuP back pain | NHS Read code or/and treatment.  PHN: code for PHN; code for acute zoster and a code for neuropathy or neuropathic pain (3-6 months after the first acute zoster entry).  PLP: code for PLP; code for amputation and a code for neuropathy or neuropathic pain (3-24 months after the first amputation code).  DN: code for DN; a code for diabetes and a code for neuropathic pain; code for diabetic neuropathy (or diabetes and neuralgia) with a prescription for a neuropathic pain treatment (within 28 days of the date of the neuropathy/neuralgia code).  Neuropathic back pain: code for back pain and a code for neuropathy or neuropathic pain within 28 days; code for radiculopathy or back pain with a specific neuropathic pain treatment (within 28 days of the back pain date). |
| Han et al. (2023) | Identified from Hospital Prescription  Analysis Cooperative Project of China for pharmacoepidemic studies. | PHN | “patients with a diagnosis of PHN” |
| Jacob et al. (2021) | Identified from the Disease Analyzer database (IQVIA) | DN | Diabetic polyneuropathy (ICD-10: E10.4, E11.4 and G63.2) |
| Jhan and Zaman (2024) | Recruited from the outpatient department of the medicine department at the Bhagwan Mahavir Institute of Medical Sciences, Pawapuri, Nalanda, Bihar (India). | DN | NR (but prescriptions were given by a neurologist). |
| Johnson et al. (2013) | Identified from a database affiliated with OptumInsight | PHN | ICD-9-CM 053.1x |
| Johnston et al. (2014) | Identified from Medicare Supplemental database | DN  PHN | ICD-9-CM 250.6x, 357.2x, 053.1 |
| Kato et al. (2023) | Identified from the RWD database. maintained by the Health, Clinic, and Education Information Evaluation Institute (HCEI) with support from Real World Data Co., Ltd | Peripheral NeuP | ICD-10 codes (NR) |
| Knoerl et al. (2024) | Secondary analysis of phase 2 clinical trial (NCT04367490) | CIPN | “diagnosed with CIPN by their clinician and had reported a severity score of 4 or greater on the Treatment-Induced Neuropathy Assessment Scale (TNAS) for at least two of the following symptoms in the bilateral lower extremities: worst hot/ burning pain, sharp/shooting pain, tingling, numbness, or cramping.” |
| Koopman et al. (2010) | Identified from the Integrated Primary Care Information (IPCI) database | Facial NeuP | In depth process to identify and confirm the diagnoses. Additional data was requested from the GP when needed. IASP criteria was used. |
| Kuo et al. (2016) | Identified from the Inovalon’s Medical Outcomes Research for Effectiveness and Economics Registry (MORE2) database | DN | ICD-9 codes: 250.6x, 357.2x. |
| Lakkad et al. (2023) | Identified from the Surveillance Epidemiology and End Results (SEER)-Medicare Linked Database | NeuP related to breast cancer treatment | A variety of ICD-9/ICD-10 codes and CPT/HCPCS codes were used to identify the relevant procedures and NeuP. (Listed in their appendix) |
| Lin et al. (2023) | Identified from a national sample of Medicare Research Identifiable Files. | Any NeuP | ICD-9 -CM: 356, 357.1-7, 357.82, 357.89, 357.9.  ICD-10-CM: G60, G62, G63, G65.2. |
| Marcianò et al. (2024) | Recruited from patients with neuropathic  pain referred to the Ambulatory of Pain Medicine of “Renato Dulbecco” University Hospital in  Catanzaro (Calabria, Italy). | Any NeuP | Douleur Neuropathique en 4 (DN4) questionnaire’ ≥ 4 |
| Margolis et al. (2017) | Identified from Truven Health Analytics’s MarketScan Commercial Claims database | DN  PHN | ICD-9 codes: 357.2x, 053.1x.  "In addition, to confirm that the diagnosis was painful, patients in the pDPN cohort were required to have a prescription neuropathic pain medication during the first 90 days." |
| Mbrah et al. (2022) | Identified from the Minimum Data Set (MDS) 3.0 (a mandated comprehensive assessment of clinical and functional status indicators of residents in Centers for Medicare and Medicaid Services–certified nursing homes in the United States). | Any NeuP | "at least 1 primary or secondary ICD-10-CM diagnosis code" for neuropathic pain (list of codes given in their supplemental material) |
| Mittal et al. (2011) | Identified retrospectively from a tertiary neuromuscular outpatient centre for neuropathic pain. | Any NeuP | NR (neuropathic pain etiologies were grouped into the eight categories of diabetes, hereditary, infection/toxic, autoimmune, cryptogenic sensory polyneuropathy (CSPN), nerve entrapment, nutritional, and miscellaneous) |
| Muñoz-Vendrell et al. (2025) | Recruited from “consecutive patients  treated in the neurology clinic of a tertiary reference hospital” (Barcelona) | TN | “Included if they met the diagnostic criteria for TN  as outlined in the third edition of the International Classification  of Headache Disorders” |
| Nygvist et al. (2024) | Identified from Swedish primary care registry VEGA and the medical prescription registry Digitalis for the Region of Västra Götaland. | NeuP back pain | ICD-1- codes: Lumbago with sciatica (M54.4), Nerve root and plexus compressions in intervertebral disc disorders (M51.1 K), Lumbar spinal stenosis (M48.0 K), Lumbar root canal stenosis (M48.8 K) |
| Oladapo et al. (2012) | Identified from Texas Medicaid prescription claims data files | DN | 1) was prescribed any oral antidiabetic medications 2) received at least 2 prescriptions for any of the following neuropathic pain medications: duloxetine, pregabalin, gabapentin, amitriptyline, nortriptyline, and desipramine. |
| Patil et al. (2015) | Identified from the IMS-LifeLink Health Plan claims database | DN | DN identification algorithm developed by Hartsfield et al. (2008).  ICD-9-CM codes: 250.6, 337.1, 337.9, 354.9, 355.8, 356.9, and 357.2. |
| Pérez et al. (2013) | Recruited from primary care in multiple centres. | DN  PHN  TN | ICD-10 codes (NR) |
| Pillay et al. (2015) | Recruited from Chris Hani Baragwanath Hospital (Johannesburg) | HIV-SN | "Patients were screened for HIV-SN using the AIDS Clinical Trials Group (ACTG) Brief Neuropathy Screening Tool (BPNS).[15] HIV-SN was diagnosed on the basis of at least one bilateral sign (vibration sense <10 seconds using a 128 Hz tuning fork in the great toe or absent ankle reflexes) and at least one symptom (pain, paraesthesia or numbness) in both feet. Symptom severity was rated on an 11-point numerical pain rating scale (NRS) ranging from 0 (not present) to 10 (most severe imaginable)." |
| Reed et al. (2013) | Identified from the General Practice Research Database (GPRD) | DN | "one of the following: a diagnosis of DN; a diagnosis of diabetic neuropathy with a prescription for treatment for pain current at the date of diagnosis; a diagnosis of diabetes and neuropathic pain; and a diagnosis of both diabetes and neuralgia plus a treatment for pain current on the data of the neuralgia code." |
| Reynolds et al. (2020a) | Recruited from the neurologists at the University of Michigan | Peripheral NeuP | Identified using ICD-10 codes (G60-G65, E08-11.40/42, E13.40/42, M79.2, A36.83, B27.01/11/81/91, B26.84, B02.23, M34.83) or when “peripheral neuropathy” was included as the chief complaint or in the problem summary list.  Only patients/data with uncontrolled pain were included. |
| Reynolds et al. (2020b) | Identified from Clinformatics Datamart database | Peripheral NeuP | Peripheral neuropathy (356 [all-inclusive], 357 [except 357.0, 357.81], G60, G62, G63, G652 |
| Sadosky et al. (2013) | Recruited from community-based physician practises (general practitioners/primary care physicians, neurologists, pain specialists, endocrinologists, podiatrists, or rheumatologists) across US | DN | "defined as subjects with diabetic distal symmetrical sensorimotor polyneuropathy (peripheral neuropathy)" |
| Sanchez et al. (2012) | Identified from the MarketScan Commercial Claims database and Encounters and Medicare Supplemental Database (Thomson Reuters) | PHN  DN | PHN: (ICD-9-CM code 053.1)  DN: (ICD-9-CM codes 250.6, 357.2) |
| Shaparin et al. (2015) | Trial 1: “95 study centres”  Trial 2: “89 investigative sites: 57 in USA, 24 in Russia, 8 in Argentina”  Trial 3: “37 investigation sites” | PHN | Trial 1: NeuP for ≥3 months after the healing of acute herpes zoster skin rash. NRS ≥4. (No information about the diagnostic process)  Trial 2: NeuP for ≥6 months after the healing of acute herpes zoster skin rash (but started within 5 years). NRS ≥4. (No information about the diagnostic process)  Trial 3: People with "active PHN" (No information about the diagnostic process. No baseline pain score requirement.) |
| Sicras-Mainar et al. (2015) | Identified from patients included in the health plan of Badalona Serveis Assistencials (BSA) | Peripheral NeuP | ICPC-2 codes N92–N94 or ICD-9-CM, codes 350.1, 352.1, 352.9, 353.1, 352.2, 353.3, 353.6, 353.8, 354.0, 355.1, 355.5, 357.2, 357.4, 357.8, 357.9, 053.12. |
| Sicras-Mainar et al. (2019) | Identified from the RedISS database | Peripheral NeuP | ICPC-2 codes N92–N94 or ICD-9-CM, codes 350.1, 352.1, 352.9, 353.1, 352.2, 353.3, 353.6, 353.8, 354.0, 355.1, 355.5, 357.2, 357.4, 357.8, 357.9, 053.12. |
| Sutema et al. (2018) | Recruited from the outpatients of Central Hospital of Sanglah | DN | NR |
| Toth et al. (2014) | Recruited from a tertiary care neuropathic pain clinic in Calgary | Peripheral NeuP | Diagnosis of peripheral polyneuropathy (using clinical information and laboratory and electrophysiological investigations)  DN4 questionnaire ≥4  Neuropathic pain duration ≥6 months  Neuropathic pain severity in VAS ≥4  Neuropathic pain severe enough to require pharmacological treatment |
| Udall et al. (2019) | Recruited from patients attending Brazilian general hospitals in São Paulo or Ceará, or a pain clinic Bahia | DN  PHN  (Central NeuP) PTN  PSN  NeuP back pain | Patients with chronic pain for ≥3 months were evaluated. Physicians followed their own procedures for collecting patients’ history and conducting a physical examination to ascertain whether a diagnosis of NeuP was warranted. For patients in whom a NeuP diagnosis was suspected, DN4 questionnaire was used to support the diagnosis. |
| Wang et al. (2020) | Identified from the Taiwanese National Health Insurance Research Database | DN  PHN | Herpes zoster: 053.x (ICD-9-CM); B02.x, B00.9 (ICD-10-CM)  DN: 250.x (ICD-9-CM); E08.x, E09.x, E10.x, E11.x, E13.x (ICD-10-CM) |
| Winterbottom et al. (2006) | Identified from Portland Veterans Affairs Medical Center (PVAMC) Veterans Health Information System and Technology Architecture (VistA) | Any NeuP | NR |
| Wu et al. (2009) | Identified from Thomson's Medstat MarketScan Commercial Claims database and Medicare Supplemental database | DN | ICD-9-CM: 250.6x or 357.2x |
| Wu et al. (2011) | Identified from Thomson's Medstat MarketScan Commercial Claims database | DN | ICD-9-CM: 250.6x or 357.2x |
| Yang et al. (2015) | Identified from the MarketScan Commercial and Medicare Supplemental database | DN | ICD-9 codes: 250.6x, 357.2 |
| Yeh et al. (2021) | Identified from the Truven MarketScan Commercial database and Medicare Supplement Database | DN  PHN  (NeuP related to Spinal cord injury) | ICD-9-CM codes: 250.6, 357.2, 344.0, 344.1, 344.6, 806, 952, 053.1. |
| Zhao et al. (2011) | Identified from Thomson's Medstat MarketScan Commercial Claims database | DN | ICD-9-CM: 250.6x or 357.2 |
| Zhao et al. (2012) | Identified from Veterans Integrated Service Network | DN | ICD-9-CM: 250.6x or 357.2 |

Table S3. Summary of medication included and excluded. For study-level information see Table S4.

| **Medications included in the analysis** | | |
| --- | --- | --- |
| Medication categories included without the study needing a defined list of medications | | |
| Category | Words used in the included studies | Comments (Citations of studies included in the review have been provided for infrequently used words) |
| Pain medication | Diabetic neuropathic pain medication therapy | Sutema et al. (2018) |
| Anticonvulsants | Anticonvulsants |  |
|  | Antiepileptics |  |
|  | Gabapentinoids |  |
| Antidepressants | Antidepressants |  |
|  | SNRIs |  |
|  | TCAs |  |
|  | SSRIs |  |
|  | 2nd-generation antidepressants | Gore et al. (2007a) |
|  | Tetracyclic and miscellaneous antidepressants | Gore et al. (2011a), Gore et al. (2011b) |
| Opioids | Opioids |  |
|  | Narcotics |  |
|  | Strong opioids |  |
|  | Weak opioids |  |
|  | Long-acting opioids |  |
|  | Short-acting opioids |  |
|  | Other opioid combinations with paracetamol or aspirin | Johnston et al. (2014) |
| Non-opioid analgesics | Non-opioid analgesics | Hall et al. (2008), Hall et al. (2013), Reed et al. (2013) |
|  | Non-narcotic analgesics | Sicras-Mainar et al. (2015) |
|  | NSAIDs |  |
|  | COX1 inhibitors |  |
|  | COX2 inhibitors |  |
|  | Non-selective NSAIDs |  |
| Local anaesthetics | Local anaesthetics | Hall et al. (2008) |
| Topical pain medications | Topical agents approved for neuropathic pain | Gore et al. (2011a), Gore et al. (2011b) |
|  | Rubefacients | Gore et al. (2007a) |
|  | Rubefacients/other topical antirheumatics | Hall et al. (2008) |
| NMDA antagonist | NMDA antagonists | Dworkin et al. (2012) |
| Medications named in the included studies | | |
| Category | Named medications | Comments (Citations of studies included in the review have been provided for infrequently mentioned medications) |
| Anticonvulsants | Mirogabalin | Kato et al. (2023) |
|  | Gabapentin |  |
|  | Pregabalin |  |
|  | Carbamazepine |  |
|  | Oxcarbazepine |  |
|  | Lamotrigine |  |
|  | Topiramate |  |
|  | Valproic acid / valproate |  |
|  | Phenytoin | Mbrah et al. (2022), Jonston et al. (2014) |
|  | Lacosamide | Muñoz-Vendrell et al. (2025) |
| Antidepressants | Duloxetine |  |
|  | Venlafaxine |  |
|  | Desvenlafaxine |  |
|  | Amitriptyline |  |
|  | Imipramine (pamoate) |  |
|  | Nortriptyline |  |
|  | Clomipramine |  |
|  | Bupropion |  |
|  | Citalopram |  |
|  | Paroxetine |  |
|  | Fluoxetine |  |
| Opioids | Alfentanil (hydrochloride) |  |
|  | Buprenorphine |  |
|  | Butorphanol (tartrate) |  |
|  | Codeine |  |
|  | Dezocine |  |
|  | Dihydrocodeine |  |
|  | Fentanyl |  |
|  | Hydrocodone |  |
|  | Hydromorphone |  |
|  | Levorphanol (tartrate) |  |
|  | Meperidine |  |
|  | Methadone |  |
|  | Morphine (sulphate) |  |
|  | Nalbuphine |  |
|  | Oxycodone |  |
|  | Oxymorphone |  |
|  | Pentazocine |  |
|  | Propoxyphene |  |
|  | Tapentadol |  |
|  | Tramadol |  |
| Combinations of opioid & non-opioid analgesic | Codeine & paracetamol / co-codamol |  |
|  | Dihydrocodeine & paracetamol / co-dydramol |  |
|  | Tramadol & paracetamol |  |
|  | Opioid extracts & paracetamol |  |
|  | Dextropropoxyphene & paracetamol / co-proxamol |  |
| Non-opioid analgesics | Aspirin / acetylsalicylic acid |  |
|  | Bromfenac (sodium) |  |
|  | Celecoxib |  |
|  | Diclofenac |  |
|  | Etodolac |  |
|  | Fenoprofen |  |
|  | Flurbiprofen |  |
|  | Ibuprofen |  |
|  | Indomethacin |  |
|  | Ketoprofen |  |
|  | Ketorolac (tromethamine) |  |
|  | Meclofenamate |  |
|  | Mefenamic acid |  |
|  | Meloxicam |  |
|  | Nabumetone |  |
|  | Naproxen (sodium) |  |
|  | Naproxen & lansoprazole |  |
|  | Nefopam |  |
|  | Oxaprozin |  |
|  | Piroxicam |  |
|  | Paracetamol / acetaminophen |  |
|  | Sulindac |  |
|  | Tolmetin (sodium) |  |
|  | Valdecoxib |  |
| NMDA antagonists | Dextromethorphan | Dworkin et al. (2012) |
|  | Memantine | Dworkin et al. (2012) |
| Local anaesthetics | Lidocaine |  |
|  | Prilocaine | Johnston et al. (2014) |
|  | Mexiletine | Dworkin et al. (2012), Johnston et al. (2014) |
| Other | Capsaicin |  |
|  | Botulinum toxin A |  |
|  | Baclofen |  |
|  | Clonidine |  |
| **Medications excluded from the analysis** | | |
| Category | Words used in the included studies | Comments (citations refer to studies included in the review which had analysed the medication mentioned in the related row) |
| Somewhat pain related medications | “Anaesthetics” | Hall et al. (2006), Hall et al. (2008), Hall et al. (2013) |
|  | Analgesics 02B | Sicras-Mainar et al. (2019) |
|  | “Calcium channel blockers” | Koopman et al. (2010) |
|  | Muscle relaxants |  |
|  | Oxygen | Koopman et al. (2010) |
|  | Steroids |  |
|  | Corticosteroids |  |
|  | Pamidronate disodium (Bisphosphate) | Johnston et al. (2014) |
| Antimigraine medications | Antimigraine medications |  |
|  | Triptans | Johnston et al. (2014) |
|  | Tizanidine |  |
| Sedatives | Sedatives |  |
|  | Hypnotics |  |
|  | Sleep medications |  |
|  | Benzodiazepines |  |
|  | Anxiolytics |  |
|  | Anxiety medications |  |
| Other | Vitamins |  |
|  | Herbs |  |
|  | Antidiabetic medications |  |
|  | Antipsychotics |  |
|  | “Face joint injections” | Marcianò et al. (2024) |

Table S4. Medications included and excluded from our analysis for each included study. Medication categories mentioned in the included studies are in quotation marks. If the medications in these categories were specified they are provided in brackets. COX: cyclo-oxygenase, NA: not applicable, NSAID: nonsteroidal anti-inflammatory drugs, SNRI: serotonin norepinephrine reuptake inhibitor, SSRI: selective serotonin reuptake inhibitor, TCA: tricyclic antidepressant.

| Author & Year | Medications included in our analysis | Medications excluded from our analysis | Comments |
| --- | --- | --- | --- |
| Adhikari et al. (2024) | Pregabalin  Gabapentin | NA | Predictors analysed for prescribing of these medications as individual medications. |
| Aggarwal et al. (2025) | Pregabalin  Gabapentin  Nortriptyline  Duloxetine | NA | Predictors analysed for prescribing of these medications as individual medications or combinations. Cannot be included in the recommended medication analysis because there is no comparison group. |
| Anderson et al. (2014) | Amitriptyline  Gabapentin  Pregabalin  Carbamazepine  Duloxetine  Nortriptyline  Imipramine  Capsaicin cream | NA | Predictors analysed for prescribing of these medications as a group. Included in the recommended medication analysis. |
| Anderson et al. (2015) | Amitriptyline  Tramadol  Gabapentin  Pregabalin  Carbamazepine  Duloxetine  Nortriptyline  Imipramine  Capsaicin cream | NA | Predictors analysed for prescribing of these medications as a group. Included in the recommended medication analysis. |
| Banks et al. (2023) | Gabapentin  Pregabalin | NA | Discontinuation between these medications was compared. |
| Berger et al. (2003) | Morphine  Oxycodone  Meperidine  Propoxyphene  Codeine  Tramadol  Pentazocine  Dihydrocodeine  Levorphanol  Hydrocodone  Hydromorphone  Fentanyl  Methadone | NA | Predictors analysed for prescribing of these medications as a group. |
| Billig et al. (2022) | Gabapentin  Pregabalin | NA | Predictors analysed for prescribing of these medications as a group. Cannot be included in the recommended medication analysis because no TCAs or SNRIs were included. |
| Boulanger et al. (2009) | “Any pain medication”  “Any pain medication excluding antidepressants”  “[Diabetic peripheral neuropathic pain] DPNP-related medication” (TCAs, venlafaxine, duloxetine, pregabalin, gabapentin, opioids)  “Antidepressants”  “SSRIs”  “TCAs”  Venlafaxine  Duloxetine  Bupropion  “Other antidepressants”  “Anticonvulsants”  Gabapentin  Pregabalin  “Other anticonvulsants”  “Non-narcotic analgesics”  “Narcotics”  Oxycodone  Tramadol  Codeine  Hydrocodone  Propoxyphene  “Other narcotics”  “NSAIDs”  “COX2 inhibitors”  “Other NSAIDs”  “Other topical and oral analogs” | “Antidiabetic medications” | “Other topical and oral analogs” was included because it was listed under pain medications. Included in the recommended medication analysis. |
| Butler et al. (2018) | “Opioids”  “COX inhibitors”  “Antiepileptics”  “Tricyclic antidepressants”  “Other antidepressants”  “Combinations” (“primarily COX inhibitors with opioids”) | Benzodiazepines  “Other” | “Other” was excluded because it was not defined. Included in the recommended medication analysis. |
| Callaghan et al. (2019) | “Gabapentinoids”  “TCAs”  “SNRIs”  “Opioids” | NA | Gabapentinoids, TCAs, and SNRIs were grouped as recommended medication. Included in the recommended medication analysis. |
| Chahine and Al Souheil (2021) | Pregabalin  Gabapentin  Amitriptyline  Nortriptyline  Imipramine  Desipramine  Venlafaxine  Duloxetine  Tramadol  “NSAIDs” | NA | Pregabalin, gabapentin, amitriptyline, nortriptyline, imipramine, desipramine, venlafaxine, and duloxetine were grouped as recommended medication. Included in the recommended medication analysis. |
| Chen et al. (2010a) | Codeine  Hydrocodone  Oxycodone  Pentazocine  Propoxyphene  Tramadol  Fentanyl  Hydromorphone  Meperidine  Morphine  Butorphanol  Oxymorphone  Levorphanol  Methadone | NA | The association between the use of concomitant NeuP medications and prescribing / discontinuation of opioids was analysed (as a group “opioids”) and individual opioids. Individual medication data provided for tramadol and oxycodone. (Note in in their Table 3 medications would be considered as predictors for receiving duloxetine or other standard or care medications). |
| Chen et al. (2010b) | Duloxetine | NA | Predictors for adherence of duloxetine were analysed. |
| Chen et al. (2011) | Duloxetine  “TCAs”  Venlafaxine  Gabapentin  Pregabalin  “Opioids” | NA | Compares characteristics between these medications / groups. Analyses were conducted using duloxetine as the reference.  Included in the recommended medication analysis.  Note: Their tables 1-4, report data for medications/categories individually which can be used to calculate predictors for receiving recommended medication in comparison to opioids. However, this would require re-calculating all of the data. Only key predictors added in our analysis. |
| Dieleman et al. (2008) | “Anticonvulsants”  “TCAs”  “SSRIs”  “Opioid” | “Treated”  “Not treated” | “Treated” was excluded as it included corticosteroids, muscle relaxants, “sedative/hypnotic”, benzodiazepines, and “miscellaneous”. Similarly, “not treated” was excluded as it excluded participants who did not receive the above-mentioned medications. Excluded from recommended medication analysis because no SNRIs included. |
| Dinesh Babu et al. (2024) | Pregabalin & amitriptyline  Duloxetine | Methylcobalamin (B12)  Methylcobalamin (B12) & pregabalin | The prescribing of these medications was compared between different types of NeuP. Cannot be included in the recommended medication analysis because no data reported for amitriptyline alone. |
| Dragic et al. (2020) | Gabapentin  Pregabalin | NA | Discontinuation between these medications was compared. |
| Dworkin et al. (2012) | “SSRIs”  Duloxetine  Venlafaxine  “TCAs”  Gabapentin  Pregabalin  Topical lidocaine  “Opioids”  Tramadol  Bupropion  Citalopram  Paroxetine  Carbamazepine  Lamotrigine  Oxcarbazepine  Topiramate  Valproic acid  Capsaicin (low concentration)  Mexiletine  Dextromethorphan  Memantine | “Treated”  “Not treated”  “Non recommended treatment” | Provided analyses for categories “first-line treatment”, “second-line treatment”, and “third-line treatment”.  “First-line treatment” included SSRIs, duloxetine, venlafaxine, TCAs, gabapentin, pregabalin, and topical lidocaine.  “Second-line treatment” included tramadol and other opioids.  “Third-line treatment” included bupropion, citalopram, paroxetine, carbamazepine, lamotrigine, oxcarbazepine, topiramate, valproic acid, capsaicin (low concentration), mexiletine, dextromethorphan, and memantine.  “Treated” and “non recommended treatment” were excluded as they included benzodiazepines and muscle relaxants in addition to the relevant medications. Similarly, “not treated” was excluded as they excluded participants without benzodiazepines and muscle relaxants. |
| Gewandter et al. (2020) | Pregabalin  Gabapentin  Duloxetine | NA | Predictors of prescribing of pregabalin and gabapentin were analysed. |
| Gharibian et al. (2013) | “Tricyclic antidepressants”  Venlafaxine  Duloxetine  Gabapentin  Lamotrigine  Pregabalin  Valproic acid  Topiramate  Carbamazepine | NA | Adherence and discontinuation between these medications was analysed. |
| Giannopoulos et al. (2007) | Gabapentin  Paroxetine  Citalopram | NA | Adherence between gabapentin and SSRIs was compared. |
| Gore et al. (2007a) | “Neuropathic pain-related medications only” (TCAs, antiepileptic drugs, opioids, SSRIs, SNRIs)  “Neuropathic pain medications or anti-inflammatory analgesics”  “Opioids”  “TCAs”  Amitriptyline  “Antiepileptic drugs”  Gabapentin  Carbamazepine  “Any 2nd generation antidepressant”  “SSRIs”  “SNRIs”  “NSAIDs” | “Anxiety/sleep medications”  “Benzodiazepines”  “Sleep agents”  “Sedatives/hypnotics” | Prescribing and discontinuation of these medications / categories were compared between people with pure NeuP and mixed pain. Included in the recommended medication analysis. |
| Gore et al. (2007b) | Pregabalin  Gabapentin  “Opioids”  “Anticonvulsants”  “TCAs”  “SSRIs”  “SNRIs”  Tramadol  5% lidocaine patch | NA | Predictors of prescribing of pregabalin and gabapentin were analysed.  Prescribing of other medications before and after taking pregabalin or gabapentin was analysed.  Excluded from recommended medication analysis because the only predictor was gabapentin vs pregabalin – so both groups already have recommended medication. |
| Gore et al. (2011a) | “Opioids”  “NSAIDs”  “SSRIs”  “SNRIs”  “TCAs”  “Anticonvulsants”  Tramadol  “Tetracyclic and miscellaneous antidepressants”  “Topical agents approved for neuropathic pain”  “Miscellaneous agents” (e.g. acetaminophen, butorphanol, nalbuphine, pentazocine)" | “Muscle relaxants”  “Benzodiazepines”  “Sedatives/hypnotics”  “Topical corticosteroids”  “Corticosteroids” | Prescribing of other medications before and after taking pregabalin or duloxetine was analysed.  Excluded from recommended medication analysis because the only predictor was duloxetine vs pregabalin – so both groups already have recommended medication. |
| Gore et al. (2011b) | “Opioids”  “NSAIDs”  “SSRIs”  “SNRIs”  “TCAs”  “Anticonvulsants”  Tramadol  “Tetracyclic and miscellaneous antidepressants”  “Topical agents approved for neuropathic pain”  “Miscellaneous agents” (e.g. acetaminophen, butorphanol, nalbuphine, pentazocine)" | “Muscle relaxants”  “Benzodiazepines”  “Sedatives/hypnotics”  “Topical corticosteroids”  “Corticosteroids” | Prescribing of other medications before and after taking pregabalin or gabapentin was analysed.  Excluded from recommended medication analysis because the only predictor was gabapentin vs pregabalin – so both groups already have recommended medication. |
| Goswami et al. (2023) | Gabapentin  “Opioids” | NA | Predictors for prescribing of these medications were analysed. Cannot be included in the recommended medication analysis because no SNRIs or TCAs were included. |
| Gustavsson et al. (2013) | Amitriptyline  Clomipramine  Nortriptyline  Duloxetine  Venlafaxine  Gabapentin  Pregabalin  “NSAIDs”  Aspirin  Paracetamol  “Weak opioids”  “Strong opioids”  “SSRIs” | “Triptans”  “Benzodiazepines”  “Other sedatives” | Prescribing and discontinuation of these medications was compared between people with pure NeuP and mixed pain.  Discontinuation between NeuP medications.  Amitriptyline, clomipramine, nortriptyline, duloxetine, venlafaxine, gabapentin, and pregabalin were considered as recommended medication.  Cannot be included in recommended medication analysis because there was no comparison group. |
| Hall et al. (2006) | “Analgesics (excluding low dose aspirin)”  “Anticonvulsants”  “Antidepressants” (divided into TCAs and “other”)  (Results for many individual medications in these categories are reported but it is not clear what was the full list of medications examined) | “Anaesthetics”  “Initial treatment” | “Anaesthetics” was excluded because it was no defined. “Initial treatment” was excluded because it included “anaesthetics”. Cannot be included in the recommended medication analysis because no comparison group. |
| Hall et al. (2008) | “Analgesics (excluding low dose aspirin)”  “Anticonvulsants”  “Antidepressants” (divided into TCAs and “other”)  (Results for many individual medications in these categories are reported but it is not clear what was the full list of medications examined) | “Anaesthetics”  “Initial treatment” | “Anaesthetics” was excluded because it was no defined. “Initial treatment” was excluded because it included “anaesthetics”. Included in the recommended medication analysis. |
| Hall et al. (2013) | “Opioids”  “TCAs”  “Antidepressants other than TCAs”  “Antiepileptics”  “Non-opioid analgesics”  “Rubefacients/other topical antirheumatics”  “Local anaesthetics”  “Analgesics”  (Results for many individual medications in these categories are reported but it is not clear what was the full list of medications examined) | “Anaesthetics”  “Initial treatment” | “Anaesthetics” was excluded because it was no defined. “Initial treatment” was excluded because it included “anaesthetics”. Included in the recommended medication analysis. |
| Han et al. (2023) | Tramadol | “Anticonvulsant”  “Opioid”  “Antidepressant”  “Topical drug”  Mecobalamin  Thiamine  Oxycodone  Thiamine  Cobamamide | The prescribing of concomitant tramadol was compared between pregabalin and gabapentin.  Oxycodone data was no provided for pregabalin group.  Data related to anticonvulsants, opioids, antidepressants, and topical drugs was ignored because it was not possible to determine whether the changes in these prescriptions by year would be predicted by year of diagnosis or duration of neuropathic pain. |
| Jacob et al. (2021) | Gabapentin  Pregabalin  Carbamazepine  “Antiepileptic drugs” (all other antiepileptic drugs) | NA | Predictors for prescribing of these medications were analysed. Cannot be included in the recommended medication analysis because no SNRIs or TCAs were included. |
| Jhan and Zaman (2024) | Amitriptyline  Duloxetine  Pregabalin | NA | Predictors analysed for prescribing of these medications as individual medications. |
| Johnson et al. (2013) | Pregabalin  Gabapentin | NA | Predictors for prescribing of these medications were analysed. Discontinuation between these medications was compared. Cannot be included in the recommended medication analysis because no SNRIs or TCAs were included. |
| Johnston et al. (2014) | “Non-opioid analgesics” (bromfenac sodium, celecoxib, diclofenac, etodolac, fenoprofen, flurbiprofen, ibuprofen, indomethacin, ketoprofen, ketorolac tromethamine, lansoprazole & naproxen, meclofenamate sodium, mefenamic acid, meloxicam, nabumetone, naproxen, naproxen sodium, oxaprozin, piroxicam, sulindac, tolmetin sodium, valdecoxib)  “Opioids” (alfentanil hcl, buprenorphine, butorphanol tartrate, codeine, dezocine, dihydrocodeine, fentanyl, hydrocodone, hydromorphone, levorphanol tartrate, meperidine, methadone, morphine sulfate, oxycodone, oxymorphone, pentazocine, propoxyphene, tapentadol, tramadol, and combinations with acetaminophen or aspirin or others)  “Antidepressants” (amitriptyline, clomipramine, desipramine, duloxetine, fluoxetine, imipramine, imipramine pamoate, nortriptyline, venlafaxine)  “Anticonvulsants” (carbamazepine, gabapentin, lamotrigine, oxcarbazepine, phenytoin, pregabalin, topiramate, valproate sodium, valproic acid)  “Other topical pain medications” (capsaicin cream, diclofenac cream, lidocaine patch, lidocaine/prilocaine cream" | “Concomitant medications”  “Other oral pain medications”  “All pain medications” | “Concomitant medications” was excluded because it included non-NP antidepressants, migraine medications, muscle relaxants, abdominal pain medications, anxiolytics, hypnotics/insomnia medications, etc.  “Other oral pain medications” was excluded because it included baclofen, clonidine, mexiletine, pamidronate disodium, tizanidine HCL as a group.  “All pain medications” was excluded because it included “other pain medications”.  Excluded from recommended medication analysis because antidepressants were divided into tricyclic and other, meaning that people with SNRIs could not be separated from those with fluoxetine. |
| Kato et al. (2023) | Mirogabalin | NA | Adherence and discontinuation of mirogabalin was analysed. |
| Knoerl et al. (2024) | Gabapentin  Pregabalin  Duloxetine  “Opioid”  Venlafaxine  Amitriptyline | “Steroid”  “Multiple”  “None” | “Multiple” was excluded because it included “Steroid”. “None” was excluded because it excluded people without “Steroid”. |
| Koopman et al. (2010) | Gabapentin  Pregabalin  Carbamazepine  Oxcarbazepine  Capsaicin  Lidocaine  “Opioids”  Tramadol  Valproate  Baclofen  Lamotrigine  Paracetamol  “NSAIDs”  Acetylsalicylic acid  “Antiepileptics”  “Antidepressants”  Clonidine | “Antimigraine drugs”  “Benzodiazepines”  “Calcium channel blocker”  Oxygen | Predictors of receiving recommended medication (as defined by European Federation of Neurological Societies [EFNS]) was compared between postherpetic neuralgia and trigeminal neuralgia.  Postherpetic neuralgia: TCAs, gabapentin, pregabalin, opioids, capsaicin, tramadol, lidocaine, and valproate.  Trigeminal neuralgia: carbamazepine, oxcarbazepine, baclofen, and lamotrigine.  Other medications and categories were compared between different NeuP diagnoses.  Included in the recommended medication analysis. |
| Kuo et al. (2016) | Duloxetine  Pregabalin  Gabapentin  Amitriptyline  Desipramine  Nortriptyline  Lidocaine  Tramadol  Oxycodone  Morphine  Oxymorphone  Methadone  Levorphanol  Hydrocodone  Hydromorphone | NA | Analysed predictors for prescribing and discontinuation between monopharmacotherapy and combination pharmacotherapy. |
| Lakkad et al. (2023) | Gabapentin  Pregabalin  Amitriptyline  Nortriptyline  Desipramine  Venlafaxine  Duloxetine  5% lidocaine patch  Capsaicin cream  “Opioids” | NA | Medications that were considered recommended as first choice: gabapentin, pregabalin, amitriptyline, nortriptyline, desipramine, venlafaxine, duloxetine, 5% lidocaine patch, capsaicin cream. People with these medications as first choice were compared to people with opioids as first choice. Included in the recommended medication analysis. |
| Lin et al. (2023) | “Opioids” | NA | No list of opioids considered provided, but “Opioid prescriptions were identified through National Drug Codes33 or by generic drug names; injections were excluded.” |
| Marcianò et al. (2024) | Amitriptyline  Duloxetine  Pregabalin  Gabapentin  Capsaicin cream  Lidocaine  Oxycodone (with or without naloxone)  Buprenorphine  Codeine  Tramadol  Fentanyl  Tapentadol | Eperisone  Cannabidiol + beta-caryophyllene  Cyclobenzaprine  Tizanidine  L-acetyl-carnitine  “Nutraceuticals”  Oxygen-ozone therapy  “Antipsychotics”  “Face joint injections” | Predictors analysed for prescribing of these medications as individual medications. Included in the recommended medication analysis. |
| Margolis et al. (2017) | Pregabalin  Gabapentin  “SNRIs”  “TCAs”  Lidocaine | NA | Medications considered recommended for DN included pregabalin, gabapentin, SNRIs, and TCAs. For people with PHN, also lidocaine was considered recommended. Included in the recommended medication analysis. |
| Mbrah et al. (2022) | “No prescription analgesia or adjuvant” (Anticonvulsants: Gabapentin, Pregabalin, Carbamazepine, Oxcarbazepine, Phenytoin, Topiramate, Valproic acid. Non-opioid analgesics: Acetaminophen, Celecoxib, Diclofenac, Etodolac, Ibuprofen, Meloxicam, Nabumetone, Naproxen. Opioids: Fentanyl, Hydrocodone (including combinations), Hydromorphone, Methadone, Morphine, Oxycodone (including combinations), Oxymorphone, Tapentadol, Tramadol (including combination), Acetaminophen with codeine. Antidepressants: Amitriptyline (including combinations), Buproprion, Desvenlafaxine, Doxepin, Duloxetine, Fluoxetine, Imipramine, Nortriptyline, Trazodone, Venlafaxine. Lidocaine (monotherapy or in combination with other drugs)." | NA | Analysed predictors of not receiving any pain medication = “no prescription analgesia or adjuvant”. |
| Mittal et al. (2011) | Pregabalin  Duloxetine | NA | Predictors for these medications were analysed as individual medications. |
| Muñoz-Vendrell et al. (2025) | Gabapentin  Lacosamide  Baclofen | NA | Predictors for receiving gabapentin, lacosamide, or baclofen as a second-choice medication after carbamazepine treatment were analysed. Discontinuation between these medications was compared. Cannot be included in the recommended medication analysis because no SNRIs or TCAs were included. |
| Nygvist et al. (2024) | Amitriptyline  Nortriptyline  Gabapentin  Duloxetine  Morphine  Buprenorphine  Oxycodone (including combinations)  Tapentadol  Codeine & paracetamol  Tramadol | NA | Predictors were analysed for people with neuropathic pain medications or opioids. Neuropathic pain medications were amitriptyline, nortriptyline, gabapentin, and duloxetine. Included in recommended medication analysis. |
| Oladapo et al. (2012) | “TCAs” (amitriptyline, nortriptyline, desipramine)  Gabapentin  Pregabalin  Duloxetine | NA | Adherence between these medications was analysed. |
| Patil et al. (2015) | Opioids | "DPN drugs” | "DPN [diabetic peripheral neuropathy] drugs" was excluded because it analysed all of these as a group: topical agents, anticonvulsants, opioids, antidepressants, skeletal muscle relaxants, ion channel blockers (not defined). |
| Pérez et al. (2013) | Pregabalin  “Pregabalin add-on to other treatments”  “Other existing marketed drugs for NeP according with physician own judgment” | NA | Predictors for prescribing between pregabalin and other NeuP medications were compared. Cannot be included in the recommended medication analysis because no SNRIs or TCAs were included. |
| Pillay et al. (2015) | “Analgesics” (amitriptyline alone or in combination with paracetamol & codeine, ibuprofen, codeine, paracetamol & codeine & carbamazepine. Paracetamol & codeine) | NA | It is unclear with “Analgesics” included other medications. |
| Reed et al. (2013) | “Non-opioid analgesics”  “Opioids”  “NSAIDs”  “Anticonvulsants”  “TCAs”  “SSRIs”  “SNRIs (duloxetine and venlafaxine)  Duloxetine  “Other antidepressants”  “Combination therapy” | NA | Prescribing of these medications / categories were compared during different time frames. |
| Reynolds et al. (2020a) | Gabapentin  Pregabalin  Amitriptyline  Nortriptyline  Doxepin  Duloxetine  Venlafaxine  Oxycodone  Methadone  Morphine  Buprenorphine  Naloxone  Fentanyl  Hydrocodone  Tramadol  Carbamazepine  Lidocaine  Topiramate  Zonisamine  Lamotrigine  Baclofen  Levetiracetam  “Other potential pain medications” | NA | Predictors for prescribing of these medications were analysed as a groups. Some predictors provided for individual medications. Gabapentin, pregabalin, amitriptyline, nortriptyline, doxepin, duloxetine, and venlafaxine were considered recommended medication. Included in the recommended medication analysis. |
| Reynolds et al. (2020b) | Gabapentin  Pregabalin  Duloxetine  Venlafaxine | NA | Predictors for prescribing between these medications were analysed. Adherence between these medications was analysed. Cannot be included in the recommended medication analysis because no TCAs were included. |
| Sadosky et al. (2013) | Aspirin  Acetaminophen  Ibuprofen  Naproxen | "Prescription treatments”  Supplements  Vitamins  Herbs | "Prescription treatments” was excluded as it was reported as a group including muscle relaxants and “topical agents” which was not defined, in addition to antiepileptics, TCAs, SNRIs, opioids, NSAIDs, and SSRIs. |
| Sanchez et al. (2012) | Pregabalin | NA | Adherence and discontinuation between different doses of pregabalin was analysed. |
| Shaparin et al. (2015) | Gabapentin | NA | Predictors of discontinuing gabapentin were analysed. |
| Sicras-Mainar et al. (2015) | Gabapentin  “NSAIDs”  “Non-narcotic analgesics”  “Opiates”  “Antidepressants" | “Anxiolytics” | Predictors of adherence and persistence with gabapentin were analysed. Changes in prescribing of the other medications were analysed. |
| Sicras-Mainar et al. (2019) | Pregabalin  “Antidepressants” (N06A)  “Opioids” (N02A)  “NSAIDs” (M01) | “Concomitant anxiolytics”  “Analgesics” (N02B) | Predictors of adherence and persistence with pregabalin were analysed. Changes in prescribing of the other medications were analysed. Anatomical Therapeutic Chemical Classification System codes were given as the definition of these groups. |
| Sutema et al. (2018) | "Diabetic neuropathic pain therapy" | NA | No other information was given about the medications, but it was clear that this was relevant. |
| Toth et al. (2014) | Amitriptyline  Nortriptyline  Gabapentin  Pregabalin | NA | Discontinuation between these medications was compared. In addition, predictors for discontinuation were analysed as a group. |
| Udall et al. (2019) | “Antiepileptics”  “NSAIDs”  “Opioids”  “SSRIs”  “TCAs”  “SNRIs”  “COX2 selective inhibitors”  “Local anaesthetics” | Muscle relaxants  Benzodiazepines  “Other treatments”  “None” | “Other treatments” was excluded because it was not defined, and it was not clear whether it referred to pain medications.  “None” was excluded because it would have excluded people with irrelevant medications to our analysis such as muscle relaxants. |
| Wang et al. (2020) | Pregabalin | NA | Predictors of pregabalin discontinuation were analysed. |
| Winterbottom et al. (2006) | Gabapentin | NA | Predictors of discontinuation discontinuation were analysed. |
| Wu et al. (2009) | Duloxetine  “Any antidepressant”  “SSRIs”  “TCAs”  Venlafaxine  Bupropion  “Other antidepressants”  “Any anticonvulsant agents”  Gabapentin  Pregabalin  “Other anticonvulsants”  “Any non-narcotic agent”  “Any narcotic agents”  Hydrocodone  Oxycodone  Propoxyphene  Tramadol  Codeine  “Other narcotics”  “Any NSAID”  “COX2 inhibitors”  “Other NSAIDs”  “DPNP-related medications” | NA | Adherence to different doses of duloxetine were compared. Predictors of prescribing of the other medications were compared.  “DPNP-related medications” were defined as TCAs, venlafaxine, duloxetine, pregabalin, gabapentin, and opioids (i.e., tramadol, oxycodone, morphine, hydrocodone, methadone, levorphanol).  Excluded from recommended medication analysis because the only predictor would have bee duloxetine compliance, so both groups already had recommended medication. The results show that people with higher duloxetine compliance are more likely to have other recommended medications later. |
| Wu et al. (2011) | "Duloxetine”  “Other standard of care medications” (TCAs, venlafaxine, gabapentin, and pregabalin)  “Opioids” (codeine, hydrocodone, oxycodone, pentazocine, propoxyphene, tramadol, fentanyl, hydromorphone, meperidine, morphine, butorphanol, oxymorphone, levorphanol, and methadone.) | NA | Prescribing of opioids compared between people with duloxetine and people with other medications. Discontinuation between duloxetine and “other standard of care medications” was compared. Cannot be included in the recommended medication analysis because no comparison group without recommended medication was included. |
| Yang et al. (2015) | Duloxetine  Gabapentin  Pregabalin | NA | Adherence and discontinuation between these medications were compared. |
| Yeh et al. (2021) | Pregabalin  Gabapentin  “Opioids excluding tramadol”  “SNRIs”  “TCAs”  Lidocaine  BTX | NA | Predictors of adherence and discontinuation of pregabalin were analysed. Prescribing of other concomitant medications was analysed. Cannot be included in the recommended medication analysis because gabapentinoids could not be included. |
| Zhao et al. (2011) | Duloxetine  Pregabalin  “Antidepressants”  “SSRIs”  “TCAs”  “SNRIs excluding duloxetine”  Duloxetine  “Other antidepressants”  “Anticonvulsants”  Gabapentin  Pregabalin  “Other anticonvulsants”  “Opioids”  “NSAIDs” | “Antidiabetic medications” | Predictors of adherence and discontinuation of pregabalin and duloxetine were analysed. Prescribing of other medications was analysed between these medications. The predictors for receiving duloxetine or pregabalin could be analysed. Cannot be included in the recommended medication analysis because the prescribing included the number of people with the predictive medication (duloxetine or pregabalin) [Their table 3]. |
| Zhao et al. (2012) | Duloxetine  “Other standard of care medication” (TCAs, venlafaxine, gabapentin, pregabalin) | NA | Predictors of prescribing between these medications were compared. Cannot be included in the recommended medication analysis because there was no comparison group without a recommended medication. |

Table S5. Medications considered recommended in the included studies. Most studies compared people with recommended medication and people without recommended medication, meaning that it is not clear what other medications people with recommended medications could have also received. Opioids were included in recommended medications by three studies. NA: not applicable, SNRI: serotonin norepinephrine reuptake inhibitor, SSRI: selective serotonin reuptake inhibitor, TCA: tricyclic antidepressant.

| Author & Year | Medications considered recommended | Comments |
| --- | --- | --- |
| Anderson et al. (2014) | Amitriptyline  Gabapentin  Pregabalin  Carbamazepine  Duloxetine  Nortriptyline  Imipramine  Capsaicin cream | NA |
| Anderson et al. (2015) | Amitriptyline  Tramadol  Gabapentin  Pregabalin  Carbamazepine  Duloxetine  Nortriptyline  Imipramine  Capsaicin cream | NA |
| Boulanger et al. (2009) | “TCAs”  Venlafaxine  Duloxetine  Pregabalin  Gabapentin  “Opioids” | In the article, the recommended medications are referred as “[Diabetic peripheral neuropathic pain] DPNP-related medication”. |
| Butler et al. (2018) | “Anti-epileptics”  “Tricyclic antidepressants”  “Other antidepressants” | NA |
| Callaghan et al. (2019) | “Gabapentinoids”  “TCAs”  “SNRIs” | NA |
| Chahine and Al Souheil (2021) | Pregabalin  Gabapentin  Amitriptyline  Nortriptyline  Imipramine  Desipramine  Venlafaxine  Duloxetine | NA |
| Chen et al. (2011) | Duloxetine  “TCAs”  Venlafaxine  Gabapentin  Pregabalin | People with these medications were compared to people with opioids.  Note: Their tables 1-4, report data for medications/categories individually which can be used to calculate predictors for receiving recommended medication in comparison to opioids. However, this would require re-calculating all of the data. Only key predictors added in our analysis. |
| Gore et al. (2007a) | “TCAs”  “Antiepileptic drugs”  “SNRIs”  “SSRIs  “Opioids” | Based on *Dworkin, R.H., Backonja, M., Rowbotham, M.C., Allen, R.R., Argoff, C.R., Bennett, G.J., Bushnell, M.C., Farrar, J.T., Galer, B.S., Haythornthwaite, J.A. and Hewitt, D.J., 2003. Advances in neuropathic pain: diagnosis, mechanisms, and treatment recommendations. Archives of neurology, 60(11), pp.1524-1534.* |
| Hall et al. (2008) | “Antidepressants (tricyclic)”  “Antidepressants (other)”  “Antiepileptics” | For our analysis, people with one of these medications, regardless of what other medications they received, were included in the recommended medication group. |
| Hall et al. (2013) | “Antidepressants (tricyclic)”  “Antidepressants (other)”  “Antiepileptics” | For our analysis, people with/without antidepressant or antiepileptic were compared. |
| Koopman et al. (2010) | Gabapentin  Pregabalin  Carbamazepine  Oxcarbazepine  Capsaicin  Lidocaine  “Opioids”  Tramadol  Valproate  Baclofen  Lamotrigine | Predictors of receiving recommended medication (as defined by European Federation of Neurological Societies [EFNS]) was compared between postherpetic neuralgia and trigeminal neuralgia.  Postherpetic neuralgia: TCAs, gabapentin, pregabalin, opioids, capsaicin, tramadol, lidocaine, and valproate.  Trigeminal neuralgia: carbamazepine, oxcarbazepine, baclofen, and lamotrigine. |
| Lakkad et al. (2023) | Gabapentin  Pregabalin  Amitriptyline  Nortriptyline  Desipramine  Venlafaxine  Duloxetine  5% lidocaine patch  Capsaicin cream | People with these medications as first choice were compared to people with opioids as first choice. This was a case-control study. |
| Marcianò et al. (2024) | Amitriptyline  Duloxetine  Pregabalin  Gabapentin | Other medications that could have been added to this analysis: lidocaine, capsaicin cream, tramadol, and other opioids. Gabapentin was only included for the predictor sex, but not age or BMI. |
| Margolis et al. (2017) | Pregabalin  Gabapentin  “SNRIs”  “TCAs”  Lidocaine | Prescribing of these medications as first choice was analysed. For people with diabetic neuropathy, lidocaine was not included in the recommended medications. For people with postherpetic neuralgia, lidocaine was included in the recommended medications. |
| Nygvist et al. (2024) | Amitriptyline  Nortriptyline  Gabapentin  Duloxetine | People with these medications as first choice were compared to people with opioids as first choice. However, this was not a case-control study as participants could have had recommended medication and opioids. |
| Reynolds et al. (2020a) | Gabapentin  Pregabalin  Amitriptyline  Nortriptyline  Doxepin  Duloxetine  Venlafaxine | NA |
| Udall et al. (2019) | “Antiepileptics”  “TCA”  “SNRI” | NA |

Table S6. Studies excluded in full-text screening.

| **First author** | **Year** | **Title** | **DOI** |
| --- | --- | --- | --- |
| Exclusion reason: No relevant outcomes (n=24) | | | |
| Ahn | 2025 | Comparative analysis of the therapeutic effects of pregabalin, gabapentin, and duloxetine in diabetic peripheral neuropathy: A retrospective study. | 10.1016/j.jdiacomp.2025.109001 |
| Anastassiou | 2011 | Impact of pregabalin treatment on pain, pain-related sleep interference and general well-being in patients with neuropathic pain: a non-interventional, multicentre, post-marketing study. | 10.2165/11589370-000000000-00000 |
| Anderson | 2024 | Pharmacy Closures and Anticonvulsant Medication Prescription Fills | 10.1001/jama.2024.19993 |
| Bartkova | 2024 | Current trends in pain therapy in burn patients Opioid, non-opioid and non-pharmacological effects on pain | 10.36290/far.2024.017 |
| Brown | 2019 | The use of botulinum toxin for the treatment of refractory peripheral neuropathic pain in a tertiary specialist centre | 10.1177/2049463719836538 |
| Carbonara | 2025 | Adherence to ESMO guidelines on cancer pain management and their applicability to specialist palliative care centers: An observational, prospective, and multicenter study | 10.1111/papr.13418 |
| Carlos | 2012 | Economic evaluation of duloxetine as a first-line treatment for painful diabetic peripheral neuropathy in Mexico. | 10.3111/13696998.2011.640730 |
| Dahlmiwal | 2024 | Changing age pattern and diverse outcomes of herpes zoster ophthalmicus: exploring the temporal trend, decrease in incident age and influence of treatment strategies | 10.1007/s10792-024-03369-2 |
| Fraile-Martinez | 2025 | Delving into the Perception, Use, and Context of Duloxetine in Clinical Practice: An Analysis Based on the Experience of Healthcare Professionals. | 10.3390/brainsci15070757 |
| Galindo | 2012 | Cost-effectiveness of an opioid in combination with gabapentin versus monotherapy for the treatment of neuropathic pain | 10.1111/j.1533-2500.2011.00528.x |
| Galindo | 2011 | Cost-effectiveness of an opioid in combination with gabapentin versus monotherapy for the treatment of neuropathic pain | 10.1016/j.jval.2011.08.1569 |
| García-Mata | 2018 | A survey of perceptions, attitudes, knowledge and practices of medical oncologists about cancer pain management in Spain. | 10.1007/s12094-017-1826-8 |
| Geerts | 2012 | Effective pharmacological treatment of painful diabetic neuropathy by nurse practitioners: results of an algorithm-based experience. | 10.1111/j.1526-4637.2012.01469.x |
| Iivanainen | 2025 | Development of a Comprehensive Decision Support Tool for Chemotherapy-Cycle Prescribing: Initial Usability Study. | 10.2196/62749 |
| JesusPalma | 2025 | Pharmacological Treatment of Chemotherapy-Induced Neuropathy: A Systematic Review of Randomized Clinical Trials. | 10.1016/j.pmn.2025.01.007 |
| Julien | 2023 | Real-world treatment patterns and diagnosis of charcot foot in franco-belgian diabetic foot expert centers (The EPiChar Study) | 10.1007/s00592-023-02101-3 |
| Knoerl | 2018 | Chemotherapy-Induced Peripheral Neuropathy: Use of an Electronic Care Planning System to Improve Adherence to Recommended Assessment and Management Practices. | 10.1188/18.CJON.E134-E140 |
| Marchand | 2016 | Evaluation of the impact of a patient therapeutic educational on compliance and on efficacy on allodynic symptoms of lidocaine 5% medicated plaster in postoperative localized neuropathic pains | 10.1016/j.rehab.2016.07.358 |
| Meng | 2014 | Efficacy and safety of gabapentin for treatment of postherpetic neuralgia: a meta-analysis of randomized controlled trials. | N/A |
| Nepal | 2024 | Amitriptyline, Pregabalin and Duloxetine for Treatment of Painful Diabetic Peripheral Neuropathy. | 10.33314/jnhrc.v22i01.5120 |
| Newton | 2025 | Carriers of SCN9A variants linked to inherited and acquired pain syndromes show no alteration in the prevalence of pain or analgesic usage in the UK Biobank cohort | 10.1101/2025.03.12.25323817 |
| Peppin | 2011 | Tolerability of NGX-4010, a capsaicin 8% patch for peripheral neuropathic pain. | 10.2147/JPR.S22954 |
| Roster | 2023 | Prescription trends of antidepressant, anxiolytic, and anticonvulsant medications among dermatologists from 2013 to 2020 | 10.5070/D329562418 |
| Rustagi | 2014 | Lamotrigine Versus Pregabalin in the Management of Refractory Trigeminal Neuralgia: A Randomized Open Label Crossover Trial. | 10.1007/s12663-013-0513-8 |
| Exclusion reason: No data specific to people with NeuP (n=66) | | | |
| Acharya | 2024 | Opioid therapy trajectories of patients with chronic non-cancer pain over 1 year of follow-up after initiation of short-acting opioid formulations | 10.1093/pm/pnad169 |
| Adhikari | 2024 | Prescribing Patterns and Off-Label Use of Gabapentinoid Agents at Dhulikhel Hospital, Nepal: A Cross-Sectional Study. | 10.2147/JPR.S493542 |
| Ahomäki | 2023 | Effect of Information Intervention on Prescribing Practice for Neuropathic Pain in Older Patients: A Nationwide Register-Based Study. | 10.1007/s40266-022-00993-4 |
| Ahmed | 2025 | Long-term opioid therapy in older adults: Incidence and risk factors related to patient characteristics and initial opioid dispensed | 10.1016/j.japh.2024.102311 |
| Allen | 2023 | Disparities in the use of pain medication in patients with pancreatic cancer: Focus on racial and ethnic minorities | 10.1158/1538-7755.DISP22-A026 |
| Benassayag Kaduri | 2024 | Trends in Pregabalin Use and Prescribing Patterns in the Adult Population: A 10-Year Pharmacoepidemiologic Study | 10.1007/s40263-024-01064-5 |
| Bilir | 2025 | Trends in Opioid and Non-opioid Prescriptions in Austria (2016-2021): A Nationwide Study on Utilization and Concomitant Benzodiazepine Use | 10.1007/s40122-025-00736-4 |
| Bordson | 2012 | Tricyclic antidepressants in the treatment of neuropathic pain: Is your patient taking them? | 10.1111/pme.12300 |
| Burghle | 2023 | Use of analgesics in Denmark: A national survey | 10.1111/bcpt.13837 |
| Burkill | 2017 | Pain and painkiller use among multiple sclerosis patients in Sweden | 10.1002/pds.4275 |
| Carbonara | 2024 | Pain, symptoms and therapy satisfaction in adult oncologic patients at admission to palliative care: An Italian prospective, multicenter, observational study | 10.1111/papr.13395 |
| Casagrande | 2023 | Opioid prescription and diabetes among Medicare beneficiaries | 10.1016/j.diabres.2023.110240 |
| Chaitoff | 2024 | Gabapentinoid Use by Self-Reported Indication and Level of Evidence | 10.1007/s11606-023-08418-7 |
| Chaitoff | 2025 | Assessing the Risk for Falls in Older Adults After Initiating Gabapentin Versus Duloxetine. | 10.7326/ANNALS-24-00636 |
| Chan | 2023 | Prevalence and healthcare utilization in managing herpes zoster in primary care: a retrospective study in an Asian urban population | 10.3389/fpubh.2023.1213736 |
| Chen | 2023 | Gabapentin, Concomitant Prescription of Opioids, and Benzodiazepines among Kidney Transplant Recipients | 10.2215/CJN.0000000000000019 |
| Choo | 2025 | Outcomes After a Statewide Policy to Improve Evidence-Based Treatment of Back Pain Among Medicaid Enrollees in Oregon | 10.1007/s11606-024-08776-w |
| Dooling | 2024 | Prescirption opioids following herpes zoster: An observation study amon insured adults, United States, 2007-2021 | 10.5055/jom.0845 |
| Elayyan | 2025 | Association of medication adherence and glycemic control with pain severity among patients with diabetes mellitus: a cross-sectional study from Palestine | 10.1186/s12902-025-02000-4 |
| Ellison | 2025 | Opioid Discharging Prescribing Habits for Geriatric Fracture Patient | 10.1093/ajhp/zxae345 |
| Freedman | 2008 | Pregabalin has opioid-sparing effects following augmentation mammaplasty. | 10.1016/j.asj.2008.04.004 |
| Garrell | 2023 | Characteristics, treatment, and healthcare resource utilisation in patients diagnosed with chronic pain in a United Kingdom Primary Care database | 10.1177/20494637231177771 |
| George | 2023 | Adverse drug events associated with nortriptyline compared with paroxetine and alternative medications in an older adult population: a retrospective cohort study in Southern California. | 10.1136/bmjopen-2023-076028 |
| Goudman | 2024 | Incidence and Prevalence of Pain Medication Prescriptions in Pathologies with a Potential for Chronic Pain. | 10.1097/ALN.0000000000004863 |
| Guilcher | 2021 | Prevalence of Prescribed Opioid Claims Among Persons With Traumatic Spinal Cord Injury in Ontario, Canada: A Population-Based Retrospective Cohort Study | 10.1016/j.apmr.2020.06.020 |
| Hamilton | 2024 | Understanding general practitioners' prescribing choices to patients with chronic low back pain: a discrete choice experiment. | 10.1007/s11096-023-01649-y |
| Hansen | 2023 | Epidemiological Factors Associated with Prescription of Opioids for Chronic Non-Cancer Pain in Adults: A Country-Wide, Registry-Based Study in Denmark Spans 2004-2018 | 10.2147/JPR.S388674 |
| Helm | 2024 | Pain management in hidradenitis suppurativa: a retrospective analysis of cross-sectional data from Black and White patients demonstrates racial disparity | 10.1111/ijd.17090 |
| Herrarte | 2024 | Evaluation of tricyclic antidepressant deprescribing in the treatment of diabetic peripheral neuropathy within federally qualified health centers. | 10.1016/j.japh.2024.102113 |
| Johnson | 2012 | Report of an HIV clinic-based pain management program and utilization of health status and health service by HIV patients. | 10.5055/jom.2012.0092 |
| Joyce | 2022 | Changes in Interventional Pain Physician Decision-Making, Practice Patterns, and Mental Health During the Early Phase of the SARS-CoV-2 Global Pandemic. | 10.1093/pm/pnaa294 |
| Kennedy-Hendricks | 2023 | Impact of High Deductible Health Plans on U.S. Adults With Chronic Pain. | 10.1016/j.amepre.2023.05.008 |
| Kernaghan | 2024 | Review of lidocaine plaster prescribing and monitoring in primary care | 10.1093/ijpp/riae013.061 |
| Kleebayoon | 2024 | Gabapentinoid prescribing patterns and predictors utilizing neural networks:Comment | 10.1016/j.ajem.2024.09.066 |
| Lazkani | 2013 | Do male and female general practitioners (GPs) prescribe differently the analgesics in the elderly patients suffering from chronic pain? | 10.1111/fcp.12025 |
| Lazkani | 2015 | Do Male and Female General Practitioners Differently Prescribe Chronic Pain Drugs to Older Patients? | 10.1111/pme.12659 |
| Levy | 2024 | Patterns of gabapentin prescription and of hospitalization in a national cohort of US Veterans. | 10.1093/pm/pnae027 |
| Lyu | 2024 | Clinical Predictors of Medication Compliance in Patients With Acute Herpetic Neuralgia. | 10.1016/j.pmn.2024.07.002 |
| Markotic | 2013 | Adherence to pharmacological treatment of chronic nonmalignant pain in individuals aged 65 and older | 10.1111/pme.12035 |
| Mathieson | 2018 | Worsening trends in analgesics recommended for spinal pain in primary care. | 10.1007/s00586-017-5178-4 |
| May | 2024 | Chronic pain management in primary care: Using population-based data to examine family physician practice patterns. | 10.46747/cfp.7009570 |
| McLintock | 2023 | The quality of prison primary care: cross-sectional cluster-level analyses of prison healthcare data in the North of England | 10.1016/j.eclinm.2023.102171 |
| Milani | 2025 | Dementia Medications and Their Association with Pain Medication Use in Medicare Beneficiaries with Alzheimer's Disease/Alzheimer's Disease-Related Dementias and Chronic Pain | 10.1007/s40266-025-01181-w |
| Mnatzaganian | 2024 | Sex disparities in the prevalence, incidence, and management of diabetes mellitus: an Australian retrospective primary healthcare study involving 668,891 individuals | 10.1186/s12916-024-03698-0 |
| Nagy | 2024 | Socio-demographic, clinical variables and pain among Egyptian patients of opioid use disorder with and without comorbid gabapentin use | 10.1186/s43045-024-00469-8 |
| Nesttvold | 2024 | Socioeconomic risk factors for long-term opioid use: A national registry-linkage study | 10.1002/ejp.2163 |
| New | 2025 | Persistent pain management in prison: an exploration of current practice and patient needs, facilitators and barriers to intervention engagement | 10.1016/j.physio.2025.101500 |
| Pathak | 2023 | Association of Gabapentin Use with Functional Limitations Among Stroke Survivors: A Multi-Institutional Electronic Health Records Database Analysis | 10.1016/j.jval.2023.03.159 |
| Peterson | 2025 | Medication and Therapy Profiles for Pain and Symptom Management Among Adults With Cerebral Palsy | 10.1016/j.mayocpiqo.2025.100597 |
| Ramdin | 2024 | Gabapentinoid prescribing patterns and predictors utilizing neural networks: An analysis across emergency departments Nationwide between 2012 and 2021 | 10.1016/j.ajem.2024.08.033 |
| Rankin | 2024 | Pharmacological treatment of pain in Swedish nursing homes: Prevalence and associations with cognitive impairment and depressive mood. | 10.1515/sjpain-2024-0007 |
| Rassi | 2024 | Gabapentinoid prescriptions for neuropathic and musculoskeletal pain in Lebanon. | 10.2144/fsoa-2023-0219 |
| Rees | 2024 | Patient and clinician beliefs about potential barriers to treatment of neuropathic pain for adolescents with sickle cell disease. | 10.1002/jha2.829 |
| Richeimer | 1997 | Utilization patterns of tricyclic antidepressants in a multidisciplinary pain clinic: a survey. | 10.1097/00002508-199712000-00010 |
| Samu | 2017 | Assessment of patient medication adherence among the type 2 diabetes mellitus population with peripheral diabetic neuropathy in South India. | 10.1016/j.jtumed.2016.12.006 |
| Schaffer | 2022 | Trajectories of pregabalin use and their association with longitudinal changes in opioid and benzodiazepine use | 10.1097/j.pain.0000000000002433 |
| Shrestha | 2024 | Comprehensive assessment of pain characteristics, quality of life, and pain management in cancer patients: a multi-center cross-sectional study | 10.1007/s11136-024-03725-w |
| Stillman | 2019 | Survey on current treatments for pain after spinal cord damage | 10.1038/s41394-019-0160-5 |
| Su | 2023 | Opioids prescribing among patients with zoster-related pain in real-life: A retrospective, cohort study based on clinical database | 10.2147/JPR.S430439 |
| Sweiss | 2024 | National Healthtree Survey on Pain and Opioid Use Patterns and Perceptions Among Patients with Multiple Myeloma | 10.1182/blood-2024-210314 |
| Tran | 2025 | Prescribing Patterns of Gabapentinoids in Chronic Pain Management: A Single Institution Retrospective Chart Review. | NA |
| Vacchani | 2024 | Evaluation of Pharmacotherapy of Cancer Pain in Patients with Head and Neck Cancer at a Tertiary Care Teaching Hospital | 10.1177/0976500X241246414 |
| Westra | 2024 | Patterns of gabapentinoid use among long-term opioid users. | 10.1016/j.ypmed.2024.108046 |
| Widerström-Noga | 2003 | Types and effectiveness of treatments used by people with chronic pain associated with spinal cord injuries: influence of pain and psychosocial characteristics. | 10.1038/sj.sc.3101511 |
| Yajima | 2016 | Pregabalin prescription for terminally ill cancer patients receiving specialist palliative care in an acute hospital. | 10.1186/s40780-016-0063-6 |
| Zaganjor | 2025 | Pain management and social functioning limitations among adults with chronic pain by diabetes status: National Health Interview Survey, United States, 2019-2020 | 10.1016/j.pcd.2024.12.008 |
| Exclusion reason: No predictors (n=35) | | | |
| Able | 2014 | Duloxetine treatment adherence across mental health and chronic pain conditions. | 10.2147/CEOR.S52950 |
| Alhowiti | 2024 | Pregabalin and amitriptyline as first-line drugs among patients with painful peripheral diabetic neuropathy: a systematic review and meta-analysis. | 10.26355/eurrev_202405_36296 |
| Angarita-Fonseca | 2024 | Trajectories of opioid consumption as predictors of patient-reported outcomes among individuals attending multidisciplinary pain treatment clinics | 10.1002/pds.5706 |
| Antony | 2024 | The Prevalence and Impact of Painful Diabetic Neuropathy on Quality of Life and Pattern of Drug Usage for the Alleviation of Neuropathic Pain in Kerala, India - Cross-Sectional Survey Analysis | NA |
| Augendre | 2014 | Prescribing patterns of duloxetine in France: a prescription assessment study in real-world conditions. | 10.5414/CP201894 |
| Chan | 2023 | Prevalence and healthcare utilization in managing herpes zoster in primary care: a retrospective study in an Asian urban population | 10.3389/fpubh.2023.1213736 |
| Chaudakshetrin | 2006 | A survey of patients with neuropathic pain at Siriraj Pain Clinic. | N/A |
| Chenaf | 2021 | Pharmacotherapy of chronic neuropathic pain: Recommendations versus clinical practice in a real-life setting-A pharmacoepidemiological study in France | 10.1111/fcp.12667 |
| Euasobhon | 2010 | Characterization of treatment strategies for neuropathic pain: Evidence from a pain specialist setting in Thailand | N/A |
| Gilron | 2002 | Patients' attitudes and prior treatments in neuropathic pain: a pilot study. | 10.1155/2002/274631 |
| Gilron | 2003 | Trends in opioid use for chronic neuropathic pain: a survey of patients pursuing enrollment in clinical trials. | 10.1007/BF03020185 |
| Gudin | 2016 | The treatment of postherpetic neuralgia reveals widespread use of opioids, CNS depressants, and polypharmacy | 10.1016/j.jval.2016.03.1018 |
| Huang | 2023 | Gabapentinoid Prescribing Practices at a Large Academic Medical Center | 10.1016/j.mayocpiqo.2022.12.002 |
| Jaganathan | 2025 | Treatment satisfaction and quality of life among neuropathic pain patients: A cross-sectional study | 10.54029/2025cma |
| Janzen | 2012 | The diagnosis and treatment of pain on a spinal cord rehabilitation unit | 10.1016/j.apmr.2012.08.136 |
| Jena | 2014 | Patterns of prescription and adr monitoring of drugs in the management of neuropathic pain in a tertiary care teaching hospital | N/A |
| Machado-Duque | 2018 | EVALUATION OF DIRECT COSTS AND TREATMENT PATTERNS ASSOCIATED WITH THE MANAGEMENT OF NEUROPATHIC PAIN IN COLOMBIA | 10.1016/j.jval.2018.09.2736 |
| Mathieson | 2018 | Increasing prescription of opioid analgesics and neuropathic pain medicines for spinal pain in australia | 10.1136/bmjebm-2018-111070.27 |
| Noyes | 2019 | Gabapentinoids and high-dose opioids for chronic non-malignant pain: A potentially ineffective and dangerous cocktail? | 10.1177/2049463719836538 |
| Possidente | 2009 | A survey of treatment practices in diabetic peripheral neuropathy. | 10.1016/j.pcd.2009.08.008 |
| Prawiroharjo | 2024 | Factors correlating to decisions for prescribing pharmacological treatment and referrals in suspected peripheral neuropathy cases in chat consultation-based application. | 10.1016/j.heliyon.2024.e30713 |
| Rigatto | 2024 | Gabapentin Use and Adverse Effects among Hemodialysis Patients Diagnosed with Pruritus or Neuropathic Pain in US Claims Data | 10.1681/ASN.2024zp3a09kx |
| Sabitha | 2008 | Prescribing practices for painful diabetic neuropathy. | N/A |
| Salinas | 2011 | Gaps in the management of postherpetic neuralgia in the elderly population: The basik PHN survey | 10.1007/s11606-011-1730-9 |
| Schaufler | 2024 | Pregabalin Utilization and Side Effects Among Hemodialysis Patients Diagnosed With Pruritus or Neuropathic Pain in US Claims Data | 10.1016/j.jval.2024.10.157 |
| Singh | 2020 | Prescription Pattern of Drugs Used for Neuropathic Pain and Adherence to NeuPSIG Guidelines in Cancer. | 10.4103/IJPC.IJPC_172_19 |
| Song | 2017 | Incidence of taxane-induced peripheral neuropathy receiving treatment and prescription patterns in patients with breast cancer. | 10.1007/s00520-017-3631-x |
| Tetens | 2024 | Obtainment of prescribed analgesics among patients with Lyme neuroborreliosis; a nationwide, population-based matched cohort study | 10.1016/j.ttbdis.2024.102371 |
| Thanasatirakul | 2014 | Patient adherence to generic gabapentin: A pragmatic study | 10.1177/1741134314552929 |
| Überall | 2025 | CASPAR: a retrospective cohort study of the high-concentration capsaicin topical system in patients with painful diabetic peripheral neuropathy of the feet. | 10.1136/bmjdrc-2024-004864 |
| Überall | 2025 | Progressive improvements in patient-reported outcomes with the high-concentration capsaicin patch: A retrospective cohort study in patients with painful diabetic peripheral neuropathy (CASPAR study). | 10.1016/j.jdiacomp.2025.109085 |
| Valladales-Restrepo | 2023 | Chronic pain and continuity of analgesic treatment during the COVID-19 pandemic. | 10.1111/papr.13197 |
| vanKollenburgh | 2012 | Prevalence, causes, and treatment of neuropathic pain in Dutch nursing home residents: a retrospective chart review. | 10.1111/j.1532-5415.2012.04078.x |
| Wagner | 2013 | The capsaicin 8% patch for neuropathic pain in clinical practice: a retrospective analysis. | 10.1111/pme.12143 |
| Wong | 2018 | A comparison of chronic pain with and without neuropathic characteristics in a Hong Kong Chinese population: An analysis of pain related outcomes and patient help seeking behaviour. | 10.1371/journal.pone.0204054 |
| Exclusion reason: Medications/treatments irrelevant to our analysis (n=7) | | | |
| Algeffari | 2018 | Painful Diabetic Peripheral Neuropathy among Saudi Diabetic Patients is Common but Under-recognized: Multicenter Cross-sectional study at primary health care setting. | 10.4103/jfcm.JFCM_145_16 |
| Bromberg | 2024 | Healthcare costs and medical utilization patterns associated with painful and severe painful diabetic peripheral neuropathy | 10.1007/s12020-024-03954-6 |
| Knoerl | 2021 | Exploring the impact of a decision support algorithm to improve clinicians’ chemotherapy-induced peripheral neuropathy assessment and management practices: a two-phase, longitudinal study | 10.1186/s12885-021-07965-8 |
| Lucey | 2011 | Relationship of depression and catastrophizing to pain, disability, and medication adherence in patients with HIV-associated sensory neuropathy. | 10.1080/09540121.2010.543883 |
| Shrestha | 2016 | A Prospective Study of Commonly Prescribed Drugs in the Management of Neuropathic Pain and its Medication Adherence Pattern. | N/A |
| Wang | 2025 | The Economic Burden of Patients with Diabetic Peripheral Neuropathic Pain Based on a Real-World Study in China | 10.2147/CEOR.S501243 |
| Zhang | 2021 | Diabetes distress and peripheral neuropathy are associated with medication non-adherence in individuals with type 2 diabetes in primary care. | 10.1007/s00592-020-01609-2 |
| Exclusion reason: Wrong study design (n=1) | | | |
| Altier | 2005 | Management of chronic neuropathic pain with methadone: a review of 13 cases. | 10.1097/01.ajp.0000125247.95213.53 |
| Exclusion reason: Hypothetical prescribing study (n=3) | | | |
| Belsky | 2025 | Practice patterns in the diagnosis and management of chemotherapy-induced peripheral neuropathy in adolescents and young adults with cancer: a survey of oncologists. | 10.1007/s00520-025-09387-9 |
| Elhomsy | 2025 | Neuropathic Pain Management in France: A Comparison of French Recommendations Using Case-Vignette Surveys. | NA |
| Martinez | 2014 | Adherence of French GPs to Chronic Neuropathic Pain Clinical Guidelines: Results of a Cross-Sectional, Randomized, ‘‘e’’ Case-Vignette Survey | 10.1371/journal.pone.0093855 |
| Exclusion reason: Before vs after surgery (n=2) | | | |
| Nagai | 2023 | Efficacy of surgical treatment on polypharmacy of elderly patients with lumbar spinal canal stenosis: retrospective exploratory research | 10.1186/s12877-023-03853-x |
| Fuzier | 2018 | Analgesic Drug Prescription After Carpal Tunnel Surgery: A Pharmacoepidemiological Study Investigating Postoperative Pain | 10.1097/AAP.0000000000000685 |
| Exclusion reason: Carpal tunnel syndrome (n=3) | | | |
| Billig | 2020 | Inappropriate Preoperative Gabapentinoid Use Among Patients With Carpal Tunnel Syndrome. | 10.1016/j.jhsa.2020.04.011 |
| Billig | 2023 | Gabapentinoid Prescribing for Carpal Tunnel Syndrome. | 10.1177/15589447211063544 |
| Exclusion reason: RCT where discontinuation was predicted by the trial medication (n=6) | | | |
| Baron | 2009 | Efficacy and safety of 5% lidocaine (lignocaine) medicated plaster in comparison with pregabalin in patients with postherpetic neuralgia and diabetic polyneuropathy: Interim analysis from an open-label, two-stage adaptive, randomized, controlled trial | 10.2165/00044011-200929040-00002 |
| Irving | 2014 | Comparative safety and tolerability of duloxetine vs. pregabalin vs. duloxetine plus gabapentin in patients with diabetic peripheral neuropathic pain. | 10.1111/ijcp.12452 |
| Joharchi | 2019 | Efficacy and safety of duloxetine and Pregabalin in Iranian patients with diabetic peripheral neuropathic pain: a double-blind, randomized clinical trial. | 10.1007/s40200-019-00427-w |
| Lee | 2020 | Efficacy and Safety of the Controlled-release Pregabalin Tablet (GLA5PR GLARS-NF1) and Immediate-release Pregabalin Capsule for Peripheral Neuropathic Pain: A Multicenter, Randomized, Double-blind, Parallel-group, Active-controlled, Phase III Clinical Tri | 10.1016/j.clinthera.2020.10.009 |
| Majdinasab | 2019 | A comparative double-blind randomized study on the effectiveness of Duloxetine and Gabapentin on painful diabetic peripheral polyneuropathy. | 10.2147/DDDT.S185995 |
| Stacey | 2008 | Pregabalin for postherpetic neuralgia: placebo-controlled trial of fixed and flexible dosing regimens on allodynia and time to onset of pain relief. | 10.1016/j.jpain.2008.05.014 |
| Exclusion reason: Not a full-text article (n=30) | | | |
| Allen | 2023 | Disparities in the use of pain medication in patients with pancreatic cancer: Focus on racial and ethnic minorities | 10.1158/1538-7755.DISP22-A026 |
| Anderson | 2013 | A higher index of multiple socioeconomic deprivation predisposes to development of painful peripheral neuropathy in Type 1 diabetes | 10.1111/dme.12091_1 |
| Anderson | 2015 | Socioeconomic deprivation independently predicts symptomatic painful diabetic neuropathy in type 2 diabetes | 10.1007/s00125-014-3355-0 |
| Appleyard | 2024 | USE OF NEUROPATHIC PAIN MEDICATIONS PRIOR TO TOTAL KNEE REPLACEMENT: A NATIONAL POPULATION-BASED CASE-CONTROL STUDY IN ENGLAND | 10.1016/j.joca.2024.02.489 |
| Bax | 2019 | Painful diabetic neuropathy in the real world setting: Epidemiology, treatment and follow-up | 10.1111/jns.12312 |
| Boehmer | 2025 | Evaluation of Neuropathic Pain Management for the Treatment of Malignant Pain | 10.1093/ajhp/zxae345 |
| Brandow | 2013 | The use of neuropathic pain drugs in patients with sickle cell disease is associated with older age and female gender | 10.1182/blood.V122.21.1717.1717 |
| Brookfield | 2016 | Medication adherence, treatment patterns, and health care costs among members taking gastroretentive gabapentin versus pregabalin | 10.1016/j.jval.2016.03.994 |
| Furter | 2011 | Pregabalin is not used in accordance with national recommendations in Sweden - Analysis of dosage and treatment order 2007-2010 | 10.1002/pds.2206 |
| Garrell | 2023 | Characteristics, treatment, and healthcare resource utilisation in patients diagnosed with chronic pain in a United Kingdom Primary Care database | 10.1177/20494637231177771 |
| Gilbert | 2016 | Healthcare utilization in patients with postherpetic neuralgia | 10.1016/j.jpain.2016.01.381 |
| Heald | 2014 | Socioeconomic deprivation independently predicts symptomatic treated painful diabetic neuropathy in Type 2 diabetes | 10.1111/dme.12378_2 |
| Hung | 2015 | Prescription patterns for postherpetic neuralgia patients with or without diabetes | 10.1097/AAP.0000000000000308 |
| Johnson | 2012 | Use of opioid analgesics in patients with postherpetic neuralgia (PHN) first treated with gabapentin or pregabalin | 10.1016/j.jpain.2012.01.293 |
| Lin | 2016 | Provider response to EHR-linked patient-reported effectiveness of diabetic peripheral neuropathy treatment | 10.1007/s11606-016-3657-7 |
| Liu | 2016 | Pharmacologic treatment utilization among patients with trigeminal neuralgia | 10.1080/00325481.2016.1224633 |
| Lobo | 2013 | Comparative dose response modeling of efficacy, adverse events and discontinuation rates in neuropathic pain | 10.1007/s10928-013-9308-2 |
| Lu | 2024 | Associations between acupuncture and uses of opioids and neuropsychiatric medication following chemotherapy-induced peripheral neuropathy among patients with breast cancer | 10.1200/OP.2024.20.10_suppl.216 |
| Mshelia | 2021 | Exploring factors which influence the prescription of medicines to manage neuropathic pain from the perspective of prescribers: a qualitative study | 10.1093/ijpp/riab015.031 |
| Raouf | 2017 | A two-year retrospective study of neuropathic pain management with gabapentin or pregabalin- A re we optimizing use in clinical practice? | 10.1080/00325481.2017.1367065 |
| Risson | 2017 | A real word treatment patterns and trends in painful diabetic neuropathy and postherpatic neuralgia | 10.1016/j.jval.2017.08.950 |
| Risson | 2018 | A REVIEW ON EPIDEMIOLOGY, RISK FACTORS AND TREATMENT PATTERNS IN MODERATE AND SEVERE PAINFUL DIABETIC NEUROPATHY AND POST-HERPETIC NEURALGIA | 10.1016/j.jval.2018.09.2599 |
| Sankyo | ongoing | Real-World Pharmacological Treatment Pattern of Neuropathic Pain in China | ClinicalTrials.gov ID NCT06546202 |
| Schnarr | 2011 | Assessment of medication utilization before and after initiation of pregabalin utilizing a pharmacy claims database | N/A |
| TomeBermejo | 2017 | Multicentric multidisciplinary study on neuropathic pain knowledge and treatment patterns by various spanish specialists | 10.1007/s00586-017-5270-9 |
| Velasco | 2016 | Duloxetine in chemotherapy-induced peripheral neuropathy: Experience beyond the clinical trial | N/A |
| Vinikoor-Imler | 2018 | Identifying pain medications for trigeminal neuralgia patients in a US claims database | 10.1002/pds.4629 |
| Yang | 2014 | Real-world evidence of sub-optimal treatment of diabetic peripheral neuropathic pain in U.S | 10.2337/db14-1317-1629 |
| Yeh | 2017 | A real-world study on treatment patterns and dose titration of pregabalin for neuropathic pain | N/A |
| Zhao | 2010 | Time to opioid use among commercially-insured patients with diabetic peripheral neuropathic pain who initiated duloxetine versus other treatments-a propensity score approach | N/A |
| Exclusion reason: Not in English (n=1) | | | |
| Navarro-Artieda | 2018 | Clinical and economic consequences of treating patients with peripheral neuropathic pain with brand name or generic drugs in routine clinical practice: The effects of age and sex. | 10.1016/j.nrl.2016.03.012 |
| Exclusion reason: No data to support the relevant statements (n=1) | | | |
| Arai | 2010 | Low-dose gabapentin as useful adjuvant to opioids for neuropathic cancer pain when combined with low-dose imipramine. | 10.1007/s00540-010-0913-6 |

Table S7. Subcategories of prescribing studies. Medications considered recommended are provided in Table S5. “Pain medication” subcategory includes studies that investigated predictors for receiving pain medication compared to not receiving pain medication. “Individual medication” subcategory includes studies that investigated predictors for receiving a specific medication (e.g. gabapentin). In this review, it was no possible to analyse the predictors for all individual medications.

|  | Recommended medication (excluding opioids) | Recommended medication (including opioids) | Recommended medication as first choice | Opioids | Opioids as first choice | Non-opioid analgesics | Pain medication | Individual medication | First choice medication (not pre-defined) |
| --- | --- | --- | --- | --- | --- | --- | --- | --- | --- |
| Adhikari et al. (2024) | 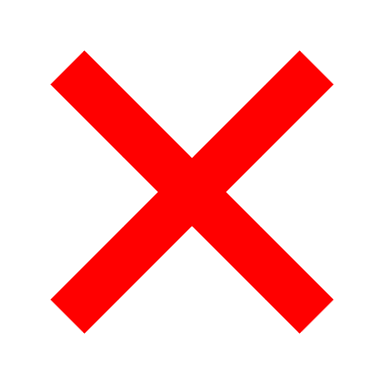 | 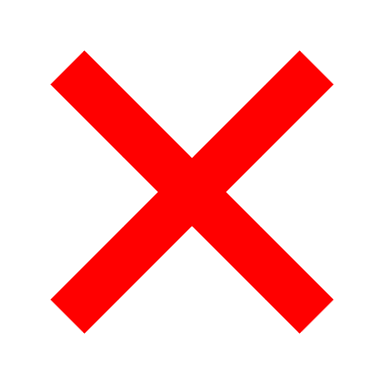 | 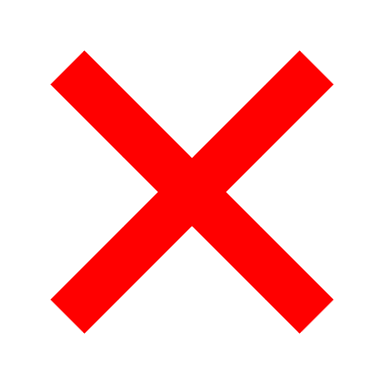 | 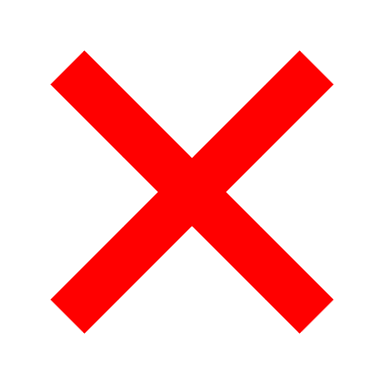 | 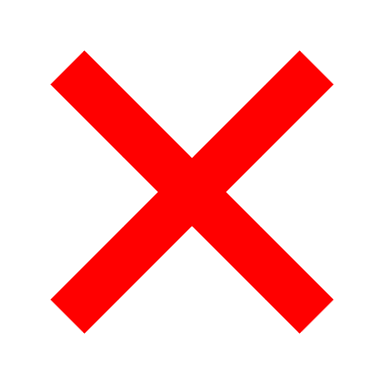 | 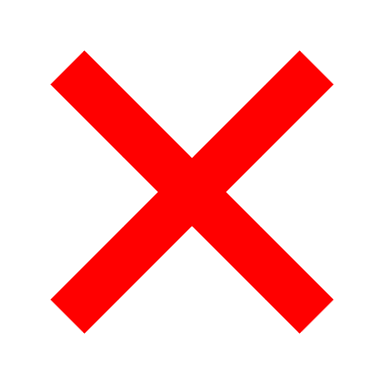 | 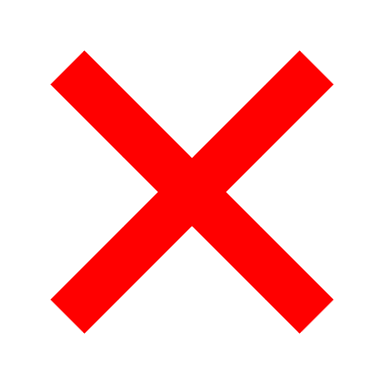 | 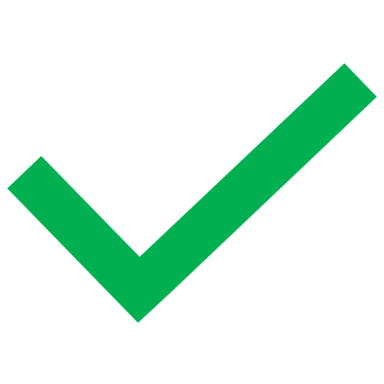 | 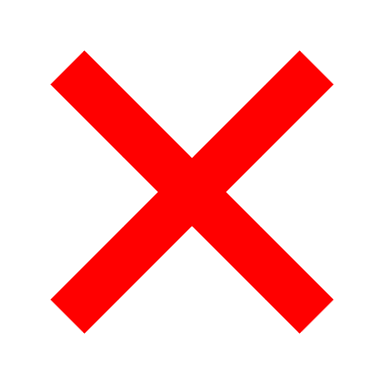 |
| Aggarwal et al. (2025) | 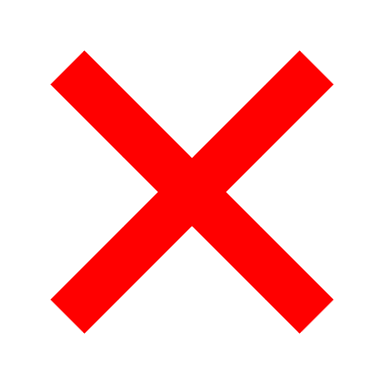 | 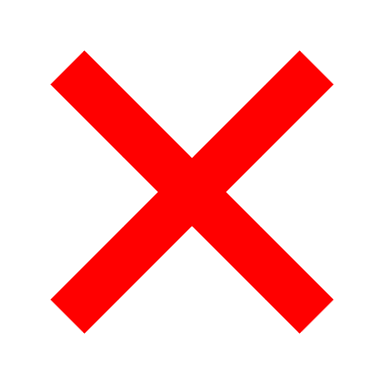 | 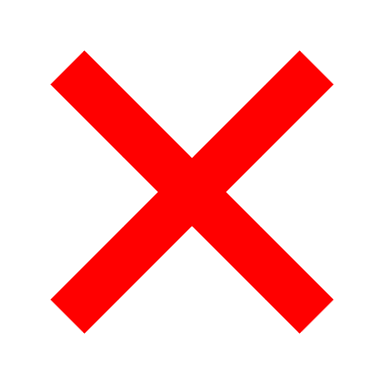 | 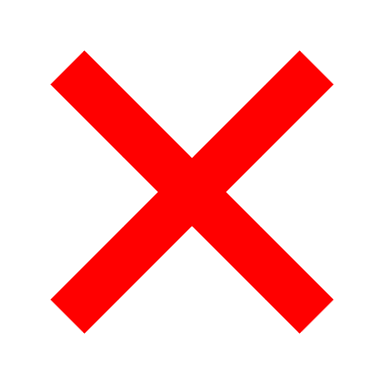 | 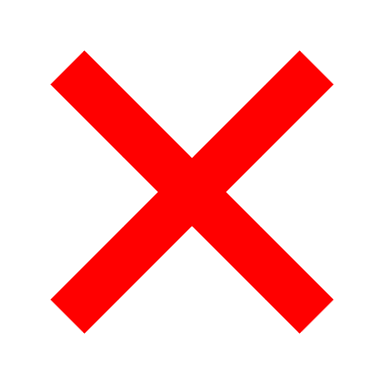 | 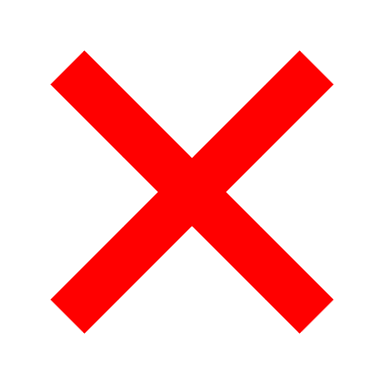 | 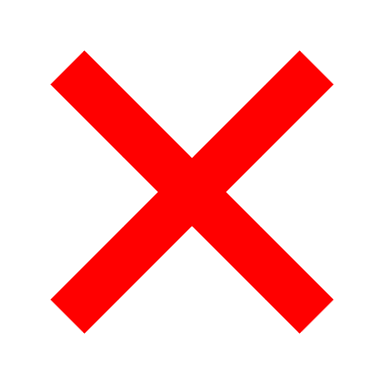 | 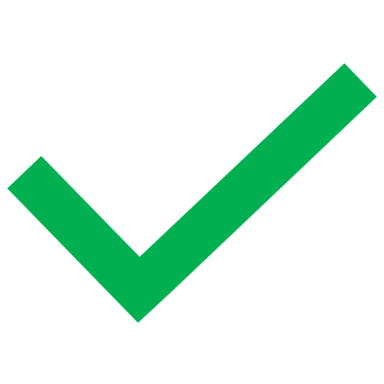 | 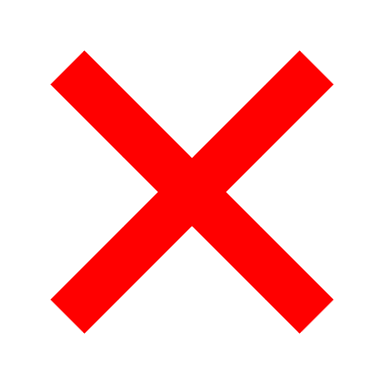 |
| Anderson et al. (2014) | 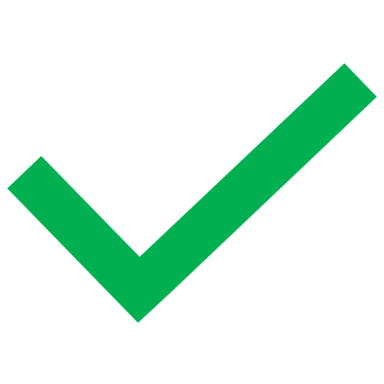 | 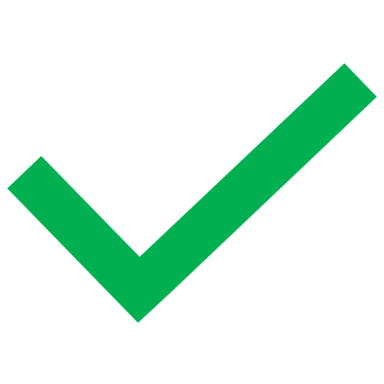 | 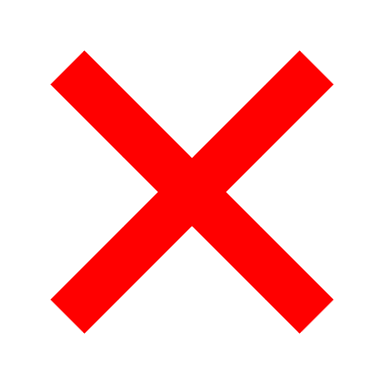 | 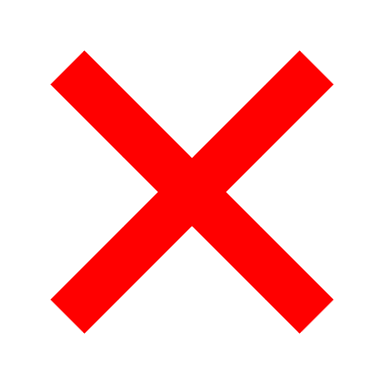 | 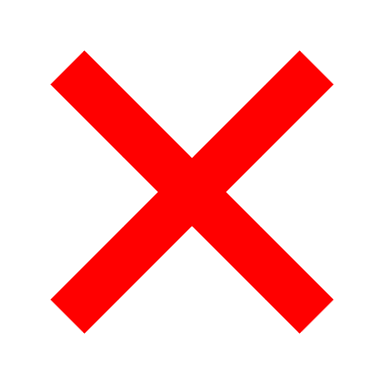 | 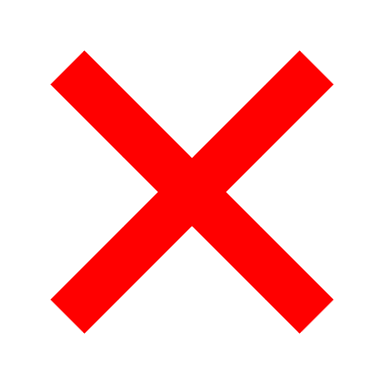 | 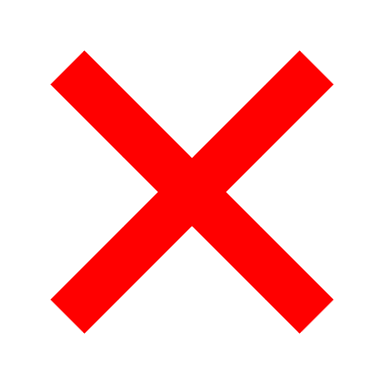 | 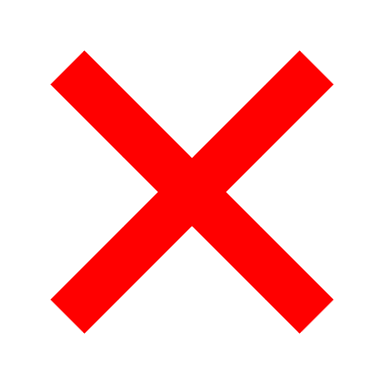 | 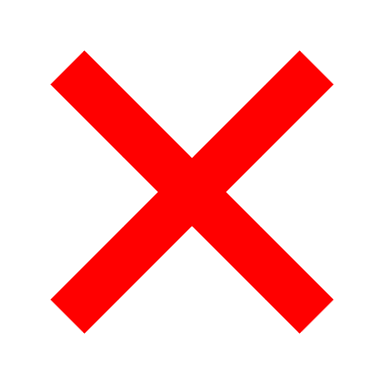 |
| Anderson et al. (2015) | 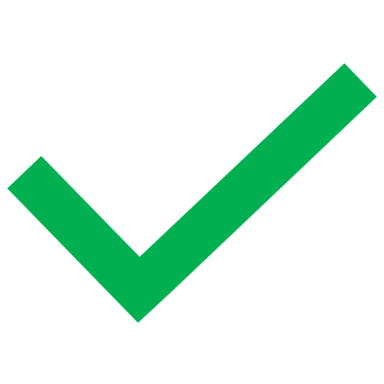 | 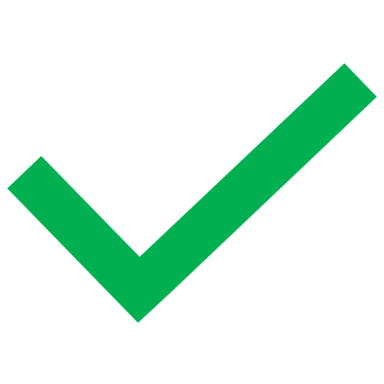 | 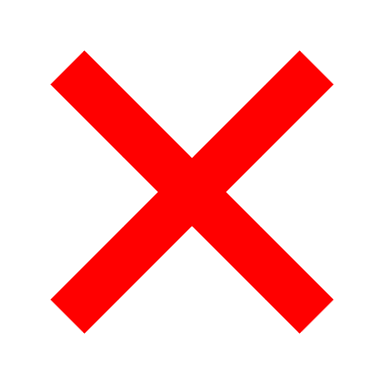 | 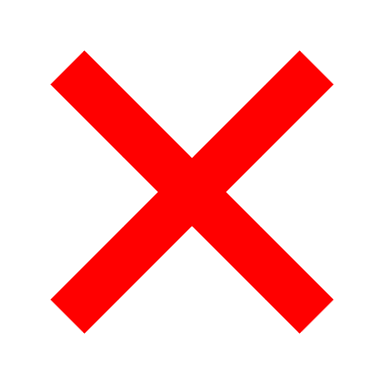 | 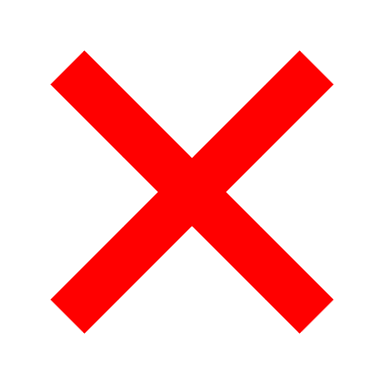 | 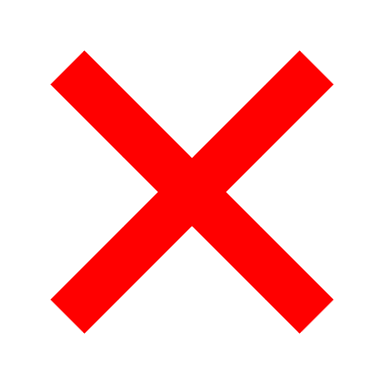 | 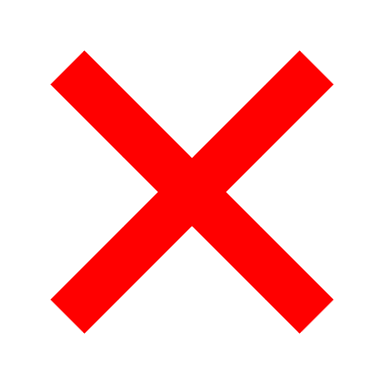 | 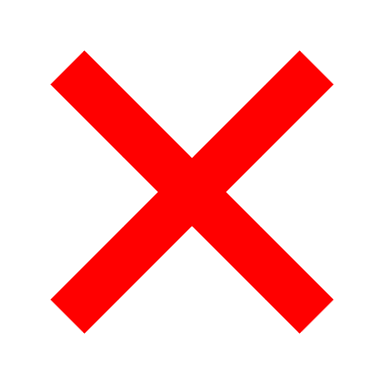 | 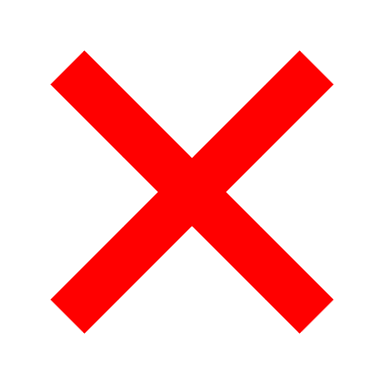 |
| Berger et al. (2003) | 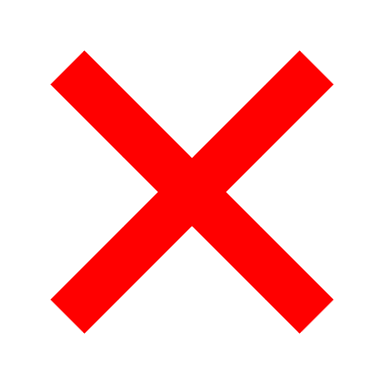 | 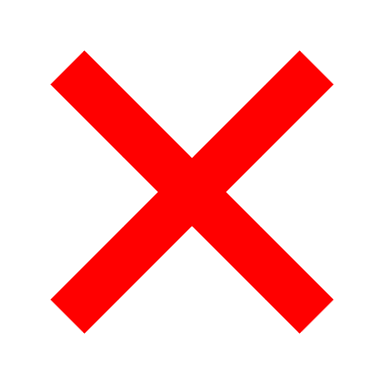 | 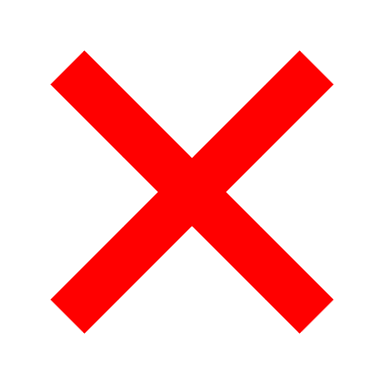 | 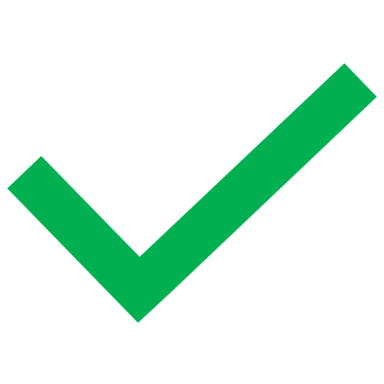 | 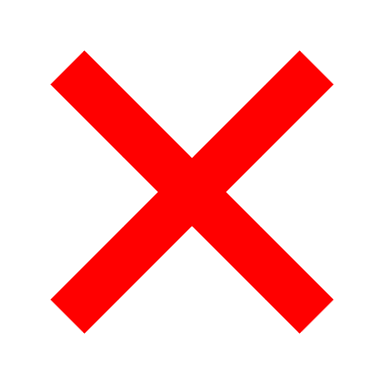 | 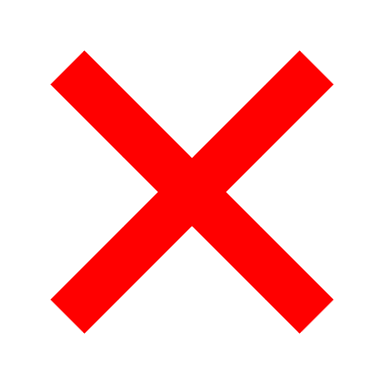 | 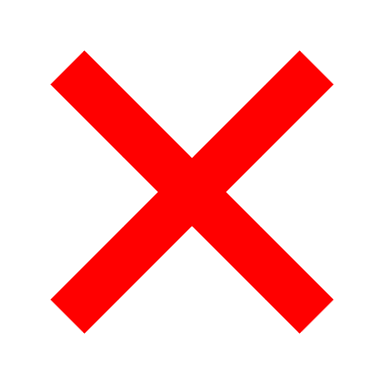 | 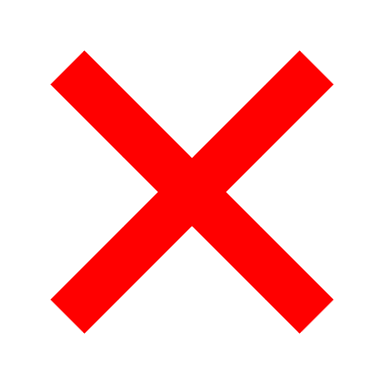 | 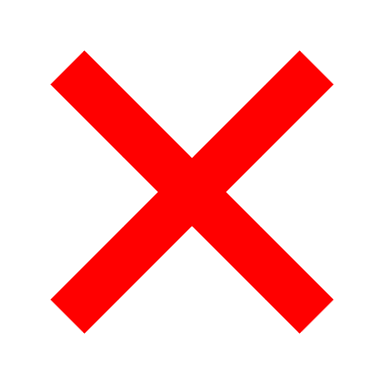 |
| Boulanger et al. (2009) | 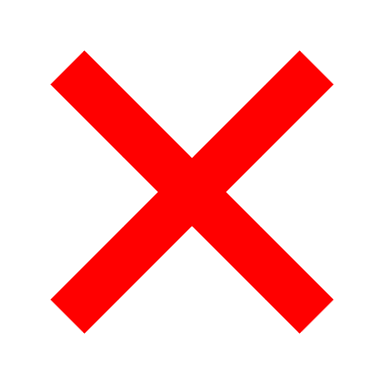 | 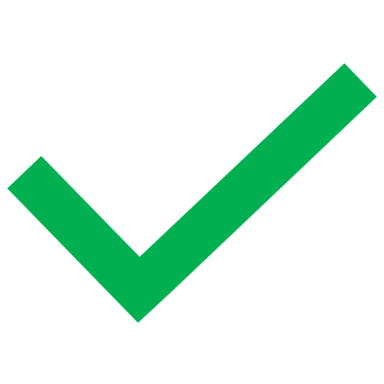 | 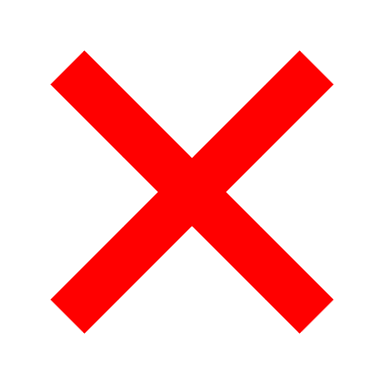 | 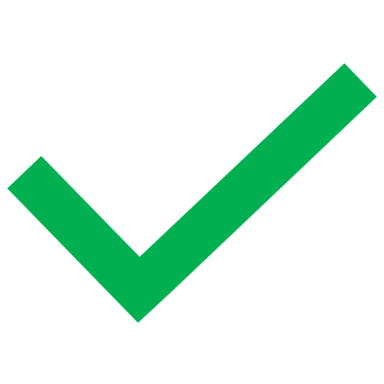 | 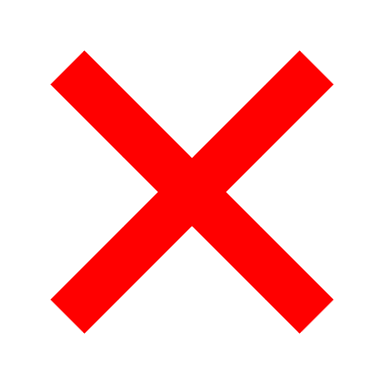 | 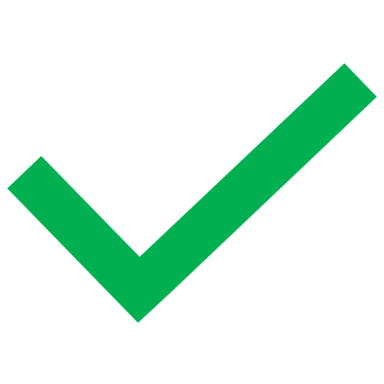 | 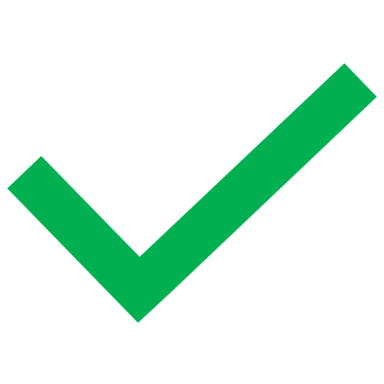 | 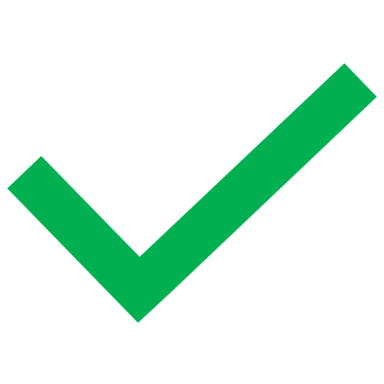 | 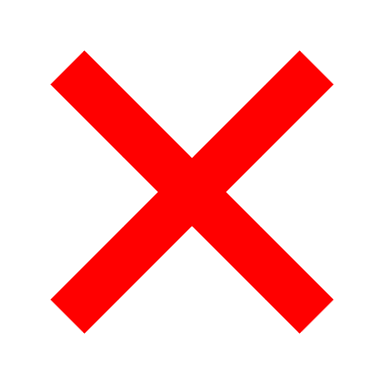 |
| Butler et al. (2018) | 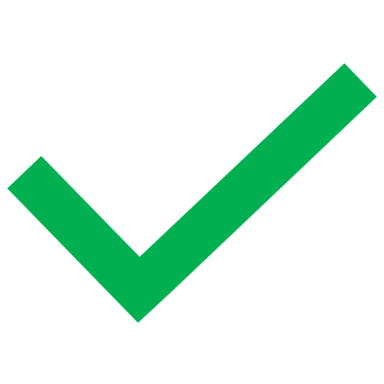 | 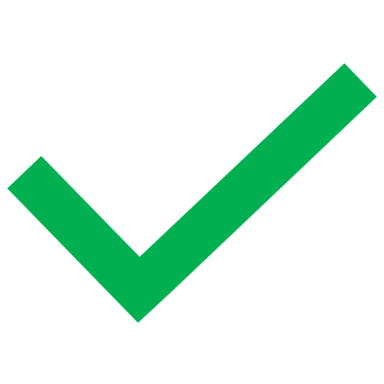 | 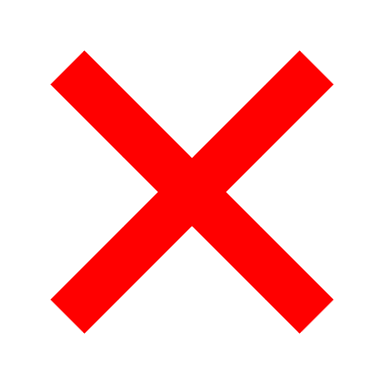 | 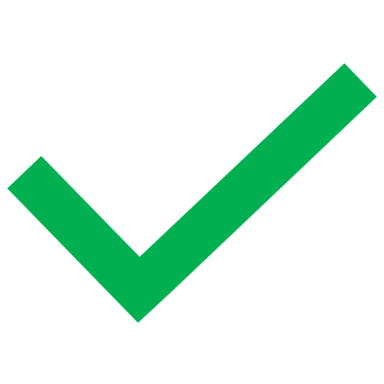 | 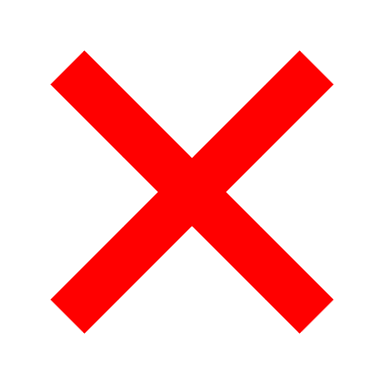 | 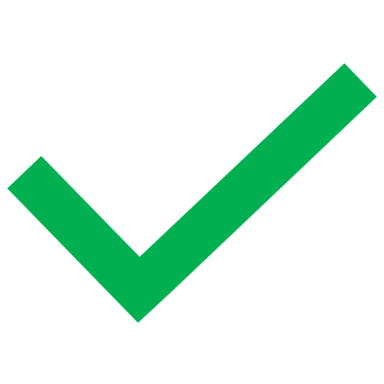 | 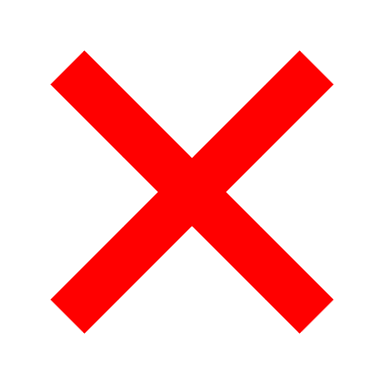 | 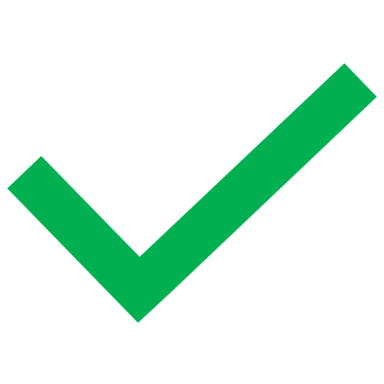 | 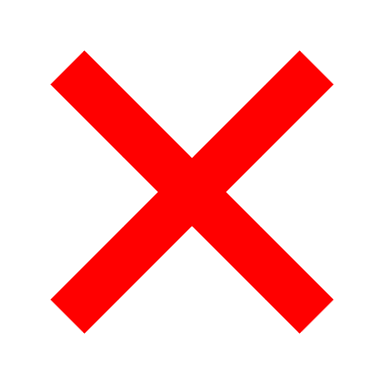 |
| Callaghan et al. (2019) | 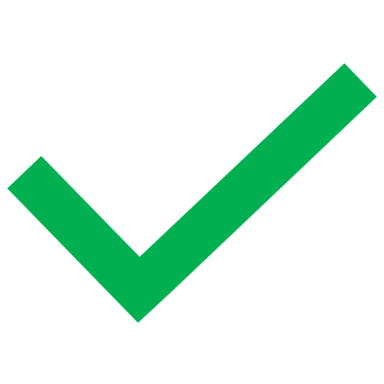 | 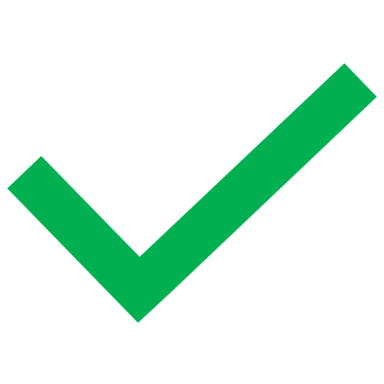 | 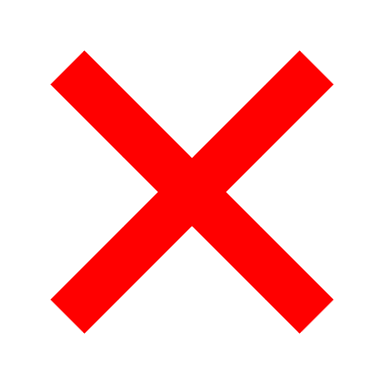 | 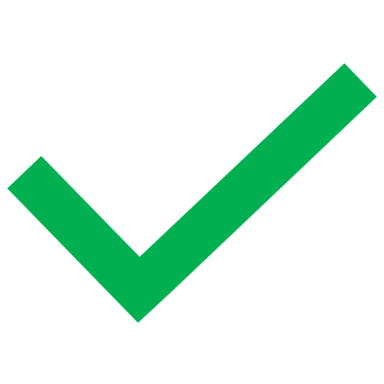 | 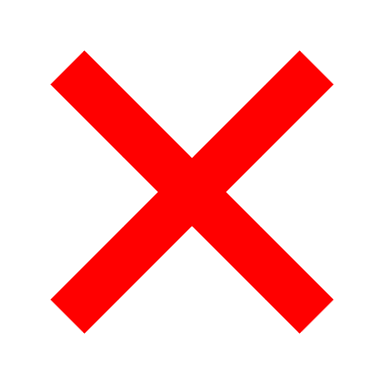 | 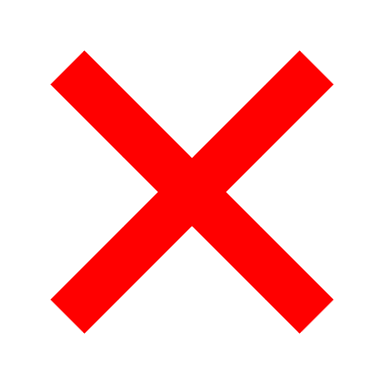 | 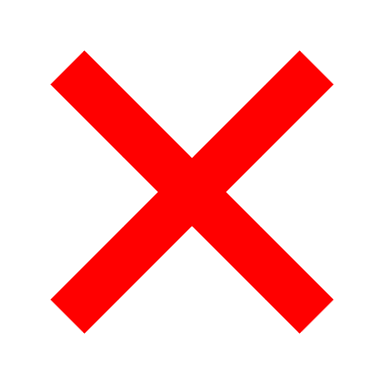 | 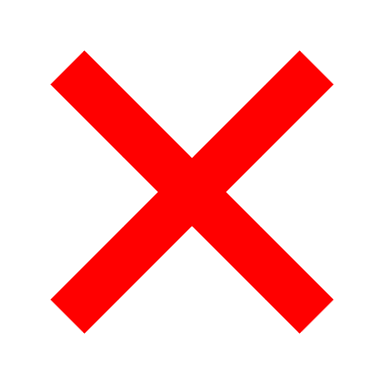 | 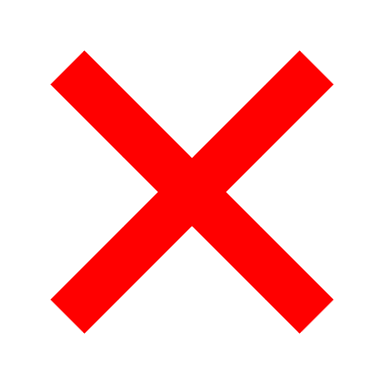 |
| Chahine et al. (2021) | 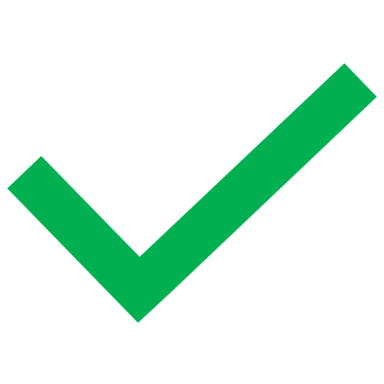 | 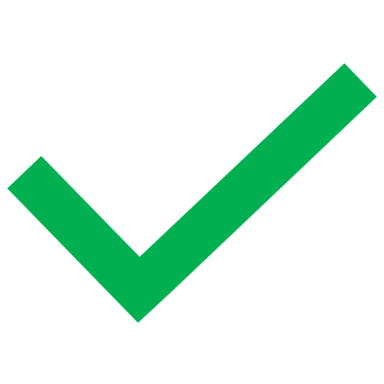 | 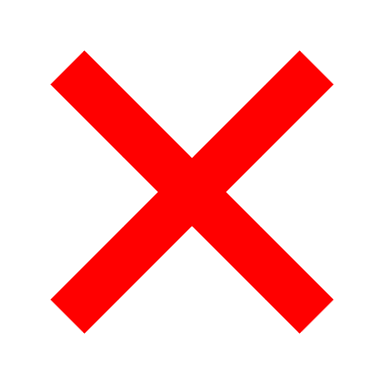 | 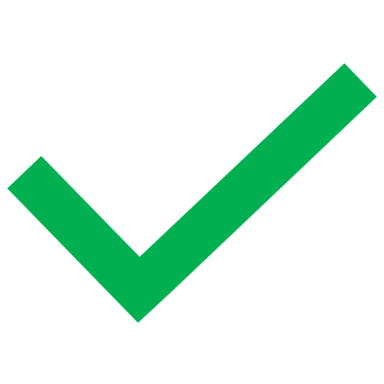 | 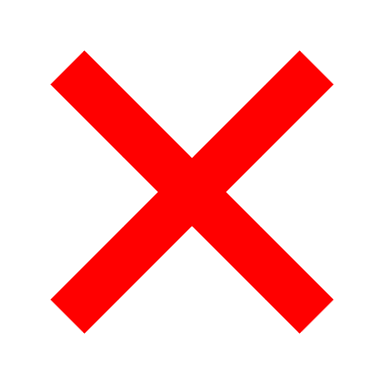 | 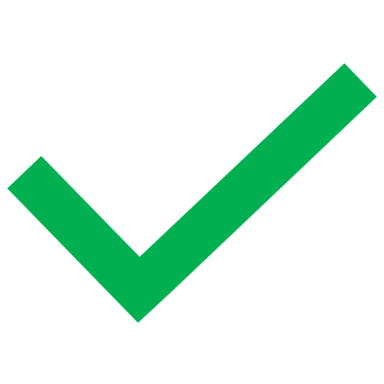 | 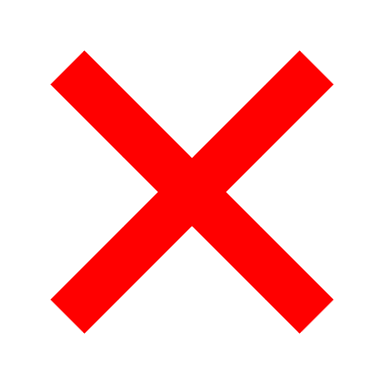 | 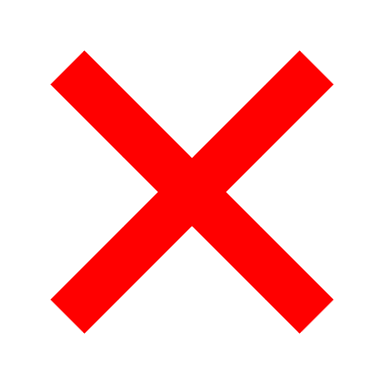 | 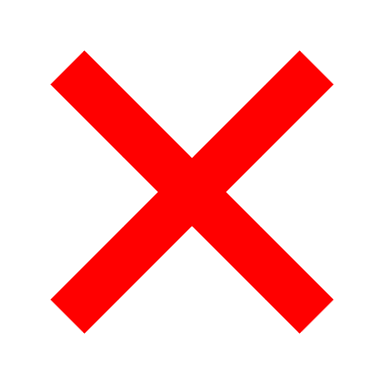 |
| Chen et al. (2010a) | 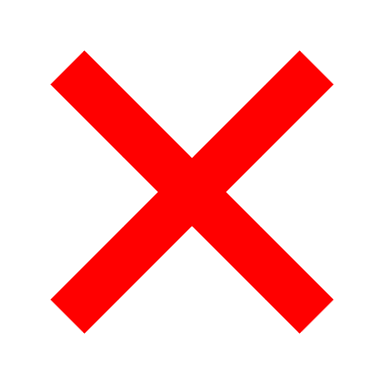 | 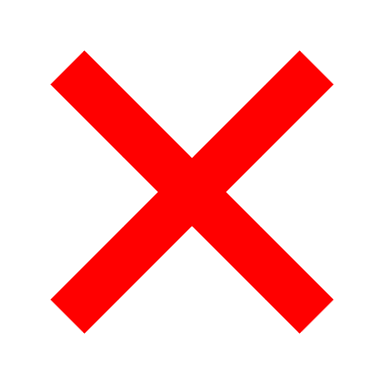 | 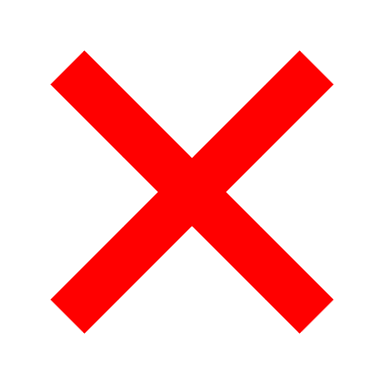 | 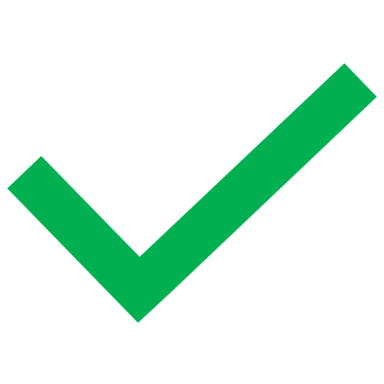 | 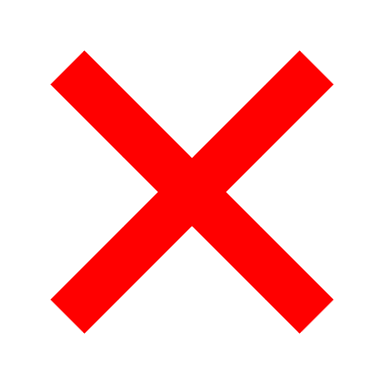 | 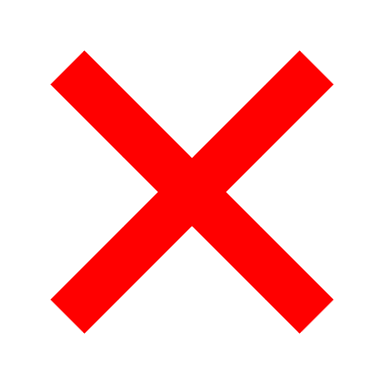 | 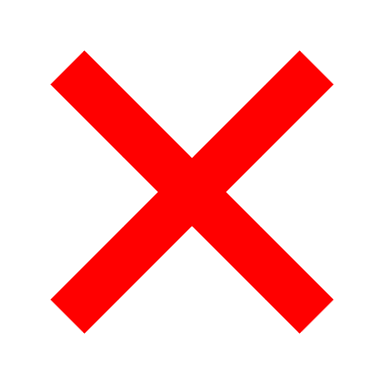 | 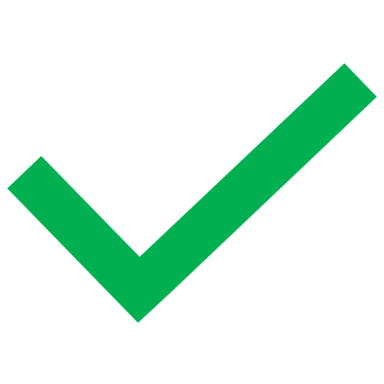 | 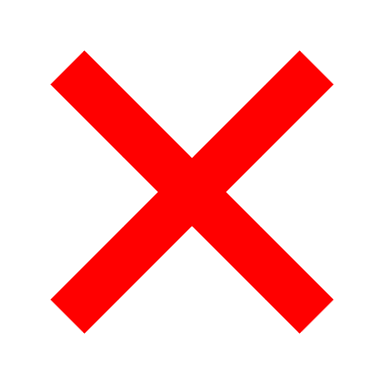 |
| Chen et al. (2011) | 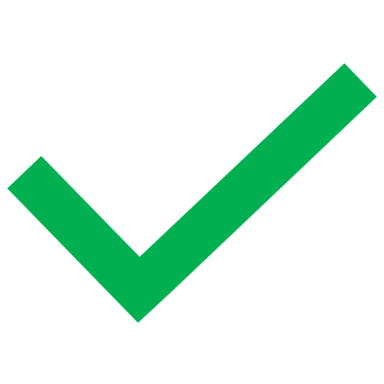 | 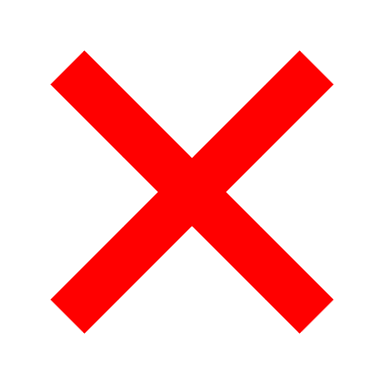 | 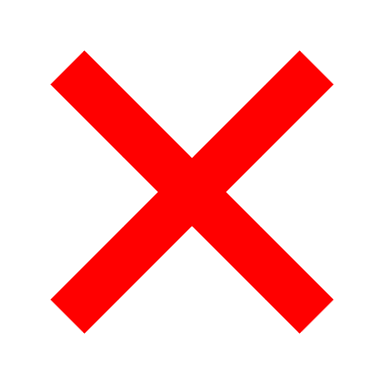 | 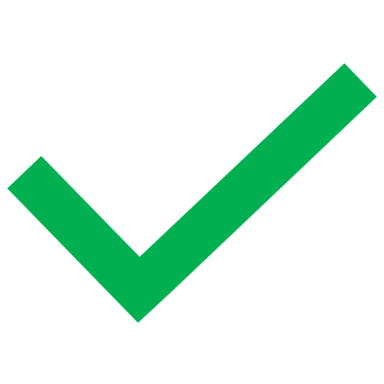 | 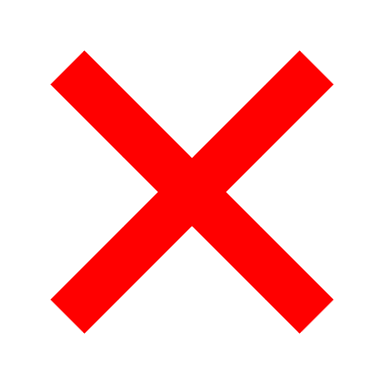 | 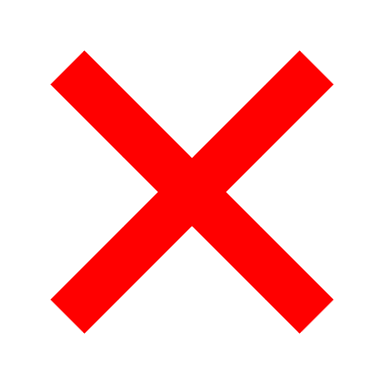 | 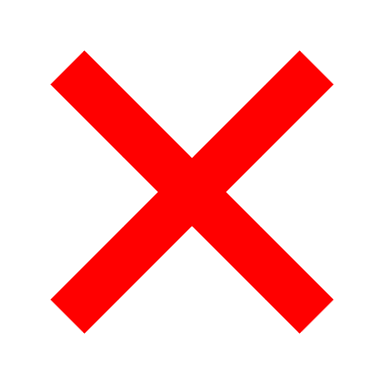 | 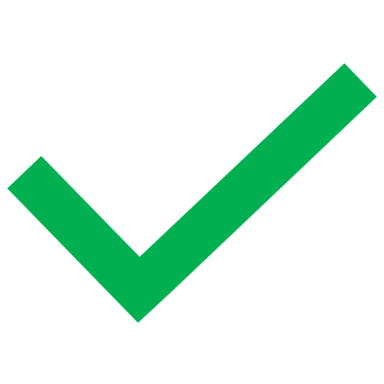 | 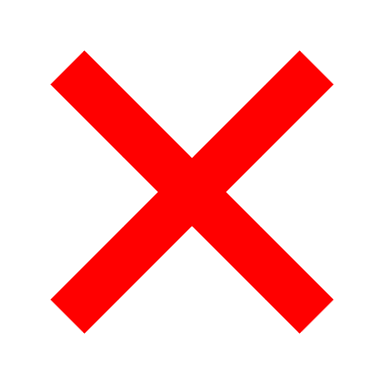 |
| Dieleman et al. (2008) | 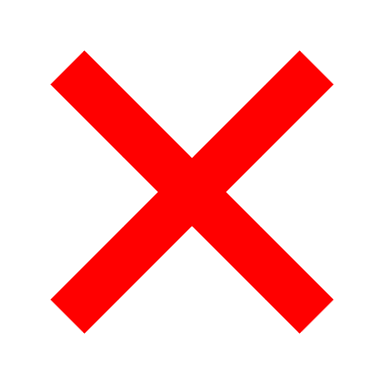 |  |  |  |  |  |  |  |  |
| Dinesh Babu et al. (2024) |  |  |  |  |  |  |  |  |  |
| Gewandter et al. (2020) |  |  |  |  |  |  |  |  |  |
| Gore et al. (2007a) |  |  |  |  |  |  |  |  |  |
| Gore et al. (2007b) |  |  |  |  |  |  |  |  |  |
| Gore et al. (2011a) |  |  |  |  |  |  |  |  |  |
| Gore et al. (2011b) |  |  |  |  |  |  |  |  |  |
| Goswami et al. (2023) |  |  |  |  |  |  |  |  |  |
| Gustavsson et al. (2013) |  |  |  |  |  |  |  |  |  |
| Hall et al. (2006) |  |  |  |  |  |  |  |  |  |
| Hall et al. (2008) |  |  |  |  |  |  |  |  |  |
| Hall et al. (2013) |  |  |  |  |  |  |  |  |  |
| Han et al. (2023) |  |  |  |  |  |  |  |  |  |
| Jacob et al. (2021) |  |  |  |  |  |  |  |  |  |
| Jha and Zaman (2024) |  |  |  |  |  |  |  |  |  |
| Johnson et al. (2013) |  |  |  |  |  |  |  |  |  |
| Johnston et al. (2014) |  |  |  |  |  |  |  |  |  |
| Knoerl et al. (2024) |  |  |  |  |  |  |  |  |  |
| Koopman et al. (2010) |  |  |  |  |  |  |  |  |  |
| Lakkad et al. (2023) |  |  |  |  |  |  |  |  |  |
| Lin et al. (2023) |  |  |  |  |  |  |  |  |  |
| Marcianò et al. (2024) |  |  |  |  |  |  |  |  |  |
| Margolis et al. (2017) |  |  |  |  |  |  |  |  |  |
| Mbrah et al. (2022) |  |  |  |  |  |  |  |  |  |
| Mittal et al. (2011) |  |  |  |  |  |  |  |  |  |
| Muñoz-Vendrell et al. (2025) |  |  |  |  |  |  |  |  |  |
| Nygvist et al. (2024) |  |  |  |  |  |  |  |  |  |
| Patil et al. (2015) |  |  |  |  |  |  |  |  |  |
| Pérez et al. (2013) |  |  |  |  |  |  |  |  |  |
| Reynolds et al. (2020a) |  |  |  |  |  |  |  |  |  |
| Reynolds et al. (2020b) |  |  |  |  |  |  |  |  |  |
| Sadosky et al. (2013) |  |  |  |  |  |  |  |  |  |
| Sicras-Mainar et al. (2015) |  |  |  |  |  |  |  |  |  |
| Sicras-Mainar et al. (2019) |  |  |  |  |  |  |  |  |  |
| Udall et al. (2019) |  |  |  |  |  |  |  |  |  |
| Wu et al. (2009) |  |  |  |  |  |  |  |  |  |
| Wu et al. (2011) |  |  |  |  |  |  |  |  |  |
| Yeh et al. (2021) |  |  |  |  |  |  |  |  |  |
| Zhao et al. (2011) |  |  |  |  |  |  |  |  |  |
| Zhao et al. (2012) |  |  |  |  |  |  |  |  |  |
| Total number | 13 | 15 | 3 | 30 | 4 | 14 | 4 | 37 | 4 |

Table S8. Adherence assessment the included studies. MPR: medication possession ratio, PDC: proportion of days covered.

| Author & Year | Proportion of participants with MPR ≥80% | Mean MPR | Proportion of participants with PCD ≥80% | Other |
| --- | --- | --- | --- | --- |
| Chen et al. (2010b) |  |  |  |  |
| Gharibian et al. (2013) |  |  |  |  |
| Giannopoulos et al. (2007) |  |  |  | Proportion of participants taking ≥75% of their scheduled dosages assessed with pill count and interviews monthly |
| Kato et al. (2023) |  |  |  |  |
| Oladapo et al. (2012) |  |  |  |  |
| Reynolds et al. (2020b) |  |  |  |  |
| Sanchez et al. (2012) |  |  |  |  |
| Sicras-Mainar et al. (2015) |  |  |  |  |
| Sicras-Mainar et al. (2019) |  |  |  |  |
| Sutema et al. (2018) |  |  |  | Proportion of participants with ≥80% pill count |
| Wu et al. (2009) |  |  |  |  |
| Yang et al. (2015) |  |  |  |  |
| Yeh et al. (2021) |  |  |  |  |
| Zhao et al. (2011) |  |  |  |  |

Table S9. Discontinuation assessment the included studies. The included studies often defined a time period (gap duration) without medication which was considered as discontinuation.

| Author & Year | Number of treatment days | Number of prescriptions | Proportion of participants | Gap duration | Comment |
| --- | --- | --- | --- | --- | --- |
| Banks et al. (2023) |  |  |  | NR |  |
| Chen et al. (2010a) |  |  |  | NR |  |
| Chen et al. (2010b) |  |  |  | NR |  |
| Dragic et al. (2020) |  |  |  | NR | Due to adverse events |
| Dworkin et al. (2012) |  |  |  | ≥30 days |  |
| Gharibian et al. (2013) |  |  |  | ≥14 days |  |
| Gore et al. (2007a) |  |  |  | ≥75 days |  |
| Gustavsson et al. (2013) |  |  |  | ≥6 months |  |
| Hall et al. (2006) |  |  |  | ≥56 days |  |
| Hall et al. (2008) |  |  |  | ≥56 days |  |
| Hall et al. (2013) |  |  |  | ≥56 days |  |
| Johnson et al. (2013) |  |  |  | ≥30 days |  |
| Kato et al. (2023) |  |  |  | ≥30 days |  |
| Kuo et al. (2016) |  |  |  | ≥60 days |  |
| Mittal et al. (2011) |  |  |  | NR |  |
| Muñoz-Vendrell et al. (2025) |  |  |  | NR | Due to any reason, adverse events, ineffectiveness, or clinical improvement |
| Sanchez et al. (2012) |  |  |  | ≥90 days | Participants with treatment at 1 year |
| Shaparin et al. (2015) |  |  |  | NR | Due to adverse events during the randomised controlled trials |
| Sicras-Mainar et al. (2015) |  |  |  | NR | Months rather than days. Proportion of participants with treatment given at 3 months, 6 months, 9 month, and 12 months |
| Sicras-Mainar et al. (2019) |  |  |  | NR | Proportion of participants with treatment given at 3 months, 6 months, 9 month, and 12 months |
| Toth et al. (2014) |  |  |  | NR | Due to adverse events or ineffectiveness |
| Wang et al. (2020) |  |  |  | NR |  |
| Winterbottom et al. (2006) |  |  |  | NR | During the prospective study |
| Wu et al. (2011) |  |  |  | ≥90 days |  |
| Yang et al. (2015) |  |  |  | ≥90 days |  |
| Yeh et al. (2021) |  |  |  | ≥45 days |  |
| Zhao et al. (2011) |  |  |  | NR | “Total supply days” |

Table S10. Risk of bias in studies assessed with ROBINS-E. D1: confounding variables D2: exposure measurement D3: participant selection, D4: post-exposure interventions, D5: missing data, D6: outcome measurement, D7: reporting of results. = Low risk of bias, = Some concerns, = High risk of bias.

|  | D1 | D2 | D3 | D4 | D5 | D6 | D7 | Overall |
| --- | --- | --- | --- | --- | --- | --- | --- | --- |
| Adhikari et al. (2024) |  |  |  |  |  |  |  |  |
| Aggarwal et al. (2025) |  |  |  |  |  |  |  |  |
| Anderson et al. (2014) |  |  |  |  |  |  |  |  |
| Anderson et al. (2015) |  |  |  |  |  |  |  |  |
| Banks et al. (2023) |  |  |  |  |  |  |  |  |
| Berger et al. (2003) |  |  |  |  |  |  |  |  |
| Billig et al. (2023) |  |  |  |  |  |  |  |  |
| Boulanger et al. (2009) |  |  |  |  |  |  |  |  |
| Butler et al. (2018) |  |  |  |  |  |  |  |  |
| Callaghan et al. (2019) |  |  |  |  |  |  |  |  |
| Chahine and Al Souheil (2021) |  |  |  |  |  |  |  |  |
| Chen et al. (2010a) |  |  |  |  |  |  |  |  |
| Chen et al. (2010b) |  |  |  |  |  |  |  |  |
| Chen et al. (2011) |  |  |  |  |  |  |  |  |
| Dieleman et al. (2008) |  |  |  |  |  |  |  |  |
| Dinesh Babu et al. (2024) |  |  |  |  |  |  |  |  |
| Dragic et al. (2020) |  |  |  |  |  |  |  |  |
| Dworkin et al. (2012) |  |  |  |  |  |  |  |  |
| Gewandter et al. (2020) |  |  |  |  |  |  |  |  |
| Gharibian et al. (2013) |  |  |  |  |  |  |  |  |
| Gore et al. (2007a) |  |  |  |  |  |  |  |  |
| Gore et al. (2007b) |  |  |  |  |  |  |  |  |
| Gore et al. (2011a) |  |  |  |  |  |  |  |  |
| Gore et al. (2011b) |  |  |  |  |  |  |  |  |
| Goswami et al. (2023) |  |  |  |  |  |  |  |  |
| Gustavsson et al. (2013) |  |  |  |  |  |  |  |  |
| Hall et al. (2006) |  |  |  |  |  |  |  |  |
| Hall et al. (2008) |  |  |  |  |  |  |  |  |
| Hall et al. (2013) |  |  |  |  |  |  |  |  |
| Han et al. (2023) |  |  |  |  |  |  |  |  |
| Jacob et al. (2021) |  |  |  |  |  |  |  |  |
| Jha and Zaman (2024) |  |  |  |  |  |  |  |  |
| Johnson et al. (2013) |  |  |  |  |  |  |  |  |
| Johnston et al. (2014) |  |  |  |  |  |  |  |  |
| Kato et al. (2023) |  |  |  |  |  |  |  |  |
| Knoerl et al. (2024) |  |  |  |  |  |  |  |  |
| Koopman et al. (2010) |  |  |  |  |  |  |  |  |
| Kuo et al. (2016) |  |  |  |  |  |  |  |  |
| Lakkad et al. (2023) |  |  |  |  |  |  |  |  |
| Lin et al. (2023) |  |  |  |  |  |  |  |  |
| Marcianò et al. (2024) |  |  |  |  |  |  |  |  |
| Margolis et al. (2017) |  |  |  |  |  |  |  |  |
| Mbrah et al. (2022) |  |  |  |  |  |  |  |  |
| Mittal et al. (2011) |  |  |  |  |  |  |  |  |
| Muñoz-Vendrell et al. (2025) |  |  |  |  |  |  |  |  |
| Nygvist et al. (2024) |  |  |  |  |  |  |  |  |
| Oladapo et al. (2012) |  |  |  |  |  |  |  |  |
| Patil et al. (2015) |  |  |  |  |  |  |  |  |
| Pérez et al. (2013) |  |  |  |  |  |  |  |  |
| Pillay et al. (2015) |  |  |  |  |  |  |  |  |
| Reed et al. (2013) |  |  |  |  |  |  |  |  |
| Reynolds et al. (2020b) |  |  |  |  |  |  |  |  |
| Sadosky et al. (2013) |  |  |  |  |  |  |  |  |
| Sanchez et al. (2012) |  |  |  |  |  |  |  |  |
| Sicras-Mainar et al. (2015) |  |  |  |  |  |  |  |  |
| Sicras-Mainar et al. (2019) |  |  |  |  |  |  |  |  |
| Toth et al. (2014) |  |  |  |  |  |  |  |  |
| Udall et al. (2019) |  |  |  |  |  |  |  |  |
| Wang et al. (2020) |  |  |  |  |  |  |  |  |
| Winterbottom et al. (2006) |  |  |  |  |  |  |  |  |
| Wu et al. (2009) |  |  |  |  |  |  |  |  |
| Wu et al. (2011) |  |  |  |  |  |  |  |  |
| Yang et al. (2015) |  |  |  |  |  |  |  |  |
| Yeh et al. (2021) |  |  |  |  |  |  |  |  |
| Zhao et al. (2011) |  |  |  |  |  |  |  |  |
| Zhao et al. (2012) |  |  |  |  |  |  |  |  |

Table S11. Risk of bias in studies assessed with RoB 2. D1: intervention allocation and randomisation, D2: deviations of intended interventions (including blinding), D3: missing data, D4: outcome measurement, D5: reporting of results. = Low risk of bias, = Some concerns, = High risk of bias.

|  | D1 | D2 | D3 | D4 | D5 | Overall |
| --- | --- | --- | --- | --- | --- | --- |
| Giannopoulos et al. (2007) |  |  |  |  |  |  |
| Reynolds et al. (2020a) |  |  |  |  |  |  |
| Sutema et al. (2018) |  |  |  |  |  |  |

Table S12. Risk of bias in studies assessed with AMSTAR.

| **Article evaluated:** Shaparin N, Slattum PW, Bucior I, Nalamachu S. Relationships Among Adverse Events, Disease Characteristics, and Demographics in Treatment of Postherpetic Neuralgia With Gastroretentive Gabapentin. Clin J Pain 2015;31:983–991. | |
| --- | --- |
| **Overall answer NO** | **Overall answer YES** |
| 2. Did the report of the review contain an explicit statement that the review methods were established prior to the conduct of the review and did the report justify any significant deviations from the protocol? | 1. Did the research questions and inclusion criteria for the review include the components of PICO? |
| 3. Did the review authors explain their selection of the study designs for inclusion in the review? | 8. Did the review authors describe the included studies in adequate detail? |
| 4. Did the review authors use a comprehensive literature search strategy? | 11. If meta-analysis was performed did the review authors use appropriate methods for statistical combination of results? |
| 5. Did the review authors perform study selection in duplicate? | 16. Did the review authors report any potential sources of conflict of interest, including any funding they received for conducting the review? |
| 6. Did the review authors perform data extraction in duplicate? |  |
| 7. Did the review authors provide a list of excluded studies and justify the exclusions? |  |
| 9. for RCTs. Did the review authors use a satisfactory technique for assessing the risk of bias (RoB) in individual studies that were included in the review? |  |
| 10. Did the review authors report on the sources of funding for the studies included in the review? |  |
| 12. If meta-analysis was performed, did the review authors assess the potential impact of RoB in individual studies on the results of the meta-analysis or other evidence synthesis? |  |
| 13. Did the review authors account for RoB in individual studies when interpreting/ discussing the results of the review |  |
| 14. Did the review authors provide a satisfactory explanation for, and discussion of, any heterogeneity observed in the results of the review? |  |
| 15. If they performed quantitative synthesis did the review authors carry out an adequate investigation of publication bias (small study bias) and discuss its likely impact on the results of the review? |  |
| **Comments:** No database search was conducted. | |

Table S13. Analysis of predictors of being prescribed recommended medication as the first choice. Adjusted odds ratios are provided for variables that were significant in multivariable analyses where applicable. Statistically non-significant results are provided when they were needed for meta-analysis. MA: meta-analysis, OR: odds ratio, aOR: adjusted odds ratio, NS: not significant, NA: not applicable.

| Variable | Study | Reference | Comparison | Result | MA | Discussion |
| --- | --- | --- | --- | --- | --- | --- |
| Sex | Nygvist et al. (2024) | Male | Female | OR: 1.20 (95% CI: 1.16 to 1.23) | NA | In Sweden, in people with NeuP back pain, women were more likely to receive recommended medication as first choice compared to men. |
| Ethnicity | Lakkad et al. (2023) | Black | White | aOR: 1.76 (95% CI: 1.43 to 2.18) | OR: 1.57 (95% CI: 1.34 to 1.84) | In the USA, white ethnicity was associated with receiving recommended medication as first choice. |
|  |  | “Other” |  | aOR: 1.37 (95% CI: 1.07 to 1.75) |  |  |
| Geographic region | Lakkad et al. (2023) | West | Northeast | NS | NA | In the USA, there were no differences in the likelihood of receiving recommended medication as first choice between geographic regions or between urban and rural regions. |
|  |  |  | Midwest | NS | NA |  |
|  |  |  | South | NS | NA |  |
|  |  | Rural | Urban | NS | NA |  |
| Marital status | Lakkad et al. (2023) | Single, divorced, widowed | Married | NS | NA | Marital status was not associated with any differences in the likelihood of receiving recommended medication as first choice. |
| Education | Lakkad et al. (2023) | No college education | College education | NS | NA | The level of education was not associated with any differences the likelihood of receiving recommended medication as first choice. |
| Household income | Lakkad et al. (2023) | <$33K | SEVERAL | NS | NA | The level of household income was not associated with any differences the likelihood of receiving recommended medication as first choice. |
| Previous pain medication | Lakkad et al. (2023) | No continuous opioid use | Continuous opioid use | aOR: 0.61 (95% CI: 0.52 to 0.71) | NA | People who had been using opioids continuously (>90 days), were less likely to receive recommended medication as first choice. The previous use of NSAIDs or long-acting opioids was not associated with any differences the likelihood of receiving recommended medication as first choice. |
|  |  | No NSAIDs | NSAIDs | NS | NA |  |
|  |  | No long-acting opioids | Long-acting opioids | NS | NA |  |
| Other previous medication | Lakkad et al. (2023) | No benzodiazepines | Benzodiazepines | aOR: 1.22 (95% CI: 1.03 to 1.45) | NA | People who had been using antipsychotics, benzodiazepines, or other central nervous system depressants were more likely to receive recommended medication as first choice compared to people with no history of using these medications. The previous use of muscle relaxants or steroids was not associated with any differences the likelihood of receiving recommended medication as first choice. |
|  |  | No antipsychotics | Antipsychotics | aOR: 1.89 (95% CI: 1.36 to 2.63) | NA |  |
|  |  | No non-benzodiazepine CNS depressants | Non-benzodiazepine CNS depressants | aOR: 1.27 (95% CI: 1.08 to 1.50) | NA |  |
|  |  | No steroids | Steroids | NS | NA |  |
|  |  | No muscle relaxants | Muscle relaxants | NS | NA |  |
| NeuP diagnosis | Margolis et al. (2017) | PHN | DN | OR: 1.20 (95% CI: 1.12 to 1.29) | NA | People with DN were more likely to receive recommended medication as first choice than people with PHN.  People with Nerve root and plexus compressions in intervertebral disc disorders or Lumbar root canal stenosis were more likely to receive recommended medication as first choice compared to people with lumbago with sciatica. (but no difference to Lumbar spinal stenosis) |
|  | Nygvist et al. (2024) | Lumbago with sciatica | Lumbar spinal stenosis | OR: 1.02 (95% CI: 0.97 to 1.08) | NA |  |
|  |  |  | Nerve root and plexus compressions in intervertebral disc  disorders | OR: 1.20 (95% CI: 1.16 to 1.25) | NA |  |
|  |  |  | Lumbar root canal stenosis | OR: 1.14 (95% CI: 1.04 to 1.23) | NA |  |
| NeuP diagnosis year | Nygvist et al. (2024) | 2017 | 2021 | OR: 1.78 (95% CI: 1.76 to 1.80) | NA | Swedish people diagnosed around 2021 with lumbago with sciatica were more likely to receive recommended medication compared to people diagnosed around 2017. (Data for the years in the middle provided too, and for Nerve root and plexus compressions in intervertebral disc disorders) |
| Comorbidities | Lakkad et al. (2023) | No mental health disorder | Mental health disorder | aOR: 3.57 (95% CI: 3.12 to 4.09) | NA | In people with neuropathic pain related to breast cancer treatment, comorbidities that were associated with a higher likelihood of receiving recommended medication as first choice included: mental health conditions, hemiplegia, dementia, diabetes, and rheumatologic disease.  In people with neuropathic pain related to breast cancer, comorbidities that were not associated with any differences the likelihood of receiving recommended medication as first choice included: substance use disorder, non-malignant pain conditions, myocardial infraction, stroke, congestive heart failure, peptic ulcer, peripheral vascular disease, chronic obstructive pulmonary disease, and liver disease. |
|  |  | No diabetes | Diabetes | aOR: 1.32 (95% CI: 1.18 to 1.47) | NA |  |
|  |  | No hemiplegia | Hemiplegia | aOR: 1.93 (95% CI: 1.03 to 3.60) | NA |  |
|  |  | No dementia | Dementia | aOR: 1.68 (95% CI: 1.23 to 2.31) | NA |  |
|  |  | No rheumatologic disease | Rheumatologic disease | OR: 0.79 (95% CI: 0.63 to 0.98) | NA |  |
|  |  | No substance use disorder | Substance use disorder | NS | NA |  |
|  |  | No myocardial infraction | Myocardial infraction | NS | NA |  |
|  |  | No stroke | Stroke | NS | NA |  |
|  |  | No congestive heart failure | Congestive heart failure | NS | NA |  |
|  |  | No peptic ulcer | Peptic ulcer | NS | NA |  |
|  |  | No peripheral vascular disease | Peripheral vascular disease | NS | NA |  |
|  |  | No chronic obstructive pulmonary disease | Chronic obstructive pulmonary disease | NS | NA |  |
|  |  | No liver disease | Liver disease | NS | NA |  |
| Breast-cancer related variables | Lakkad et al. (2023) | No radiation | Radiation | NS | NA | There was no difference the likelihood of receiving recommended medication as first choice based on breast cancer treatment; age of diagnosis; Surveillance, Epidemiology, and End Results (SEER) grade; SEER summary stage; or hormone receptor status.  People diagnosed with breast cancer during 2013 were more likely to receive recommended first-line treatment for NeuP compared to people diagnosed with breast cancer during 2007. However, there were no significant differences in the likelihood of receiving recommended first-line treatment for NeuP between people diagnosed during the years 2008 to 2012 compared to people diagnosed with breast cancer during 2007. |
|  |  | No targeted therapy | Targeted therapy | NS | NA |  |
|  |  | Higher age at cancer diagnosis | Lower age at cancer diagnosis | NS | NA |  |
|  |  | Hormone receptor status | SEVERAL | NS | NA |  |
|  |  | Breast cancer year of diagnosis (2007) | 2013 | OR: 1.29 (95% CI: 1.04 to 1.60) | NA |  |
|  |  |  | SEVERAL | NS | NA |  |
|  |  | SEER grade | SEVERAL | NS | NA |  |
|  |  | SEER summary stage | SEVERAL | NS | NA |  |

Table S14. Analysis of predictors of being prescribed recommended medication at any point. Adjusted odds ratios are provided for variables that were significant in multivariable analyses where applicable. Statistically non-significant results are provided when they were needed for meta-analysis. MA: meta-analysis, OR: odds ratio, aOR: adjusted odds ratio, NS: not significant, NA: not applicable, NR: not reported, #: study included opioids in their definition of recommended medication.

| Variable | Study | Reference | Comparison | Result | MA | Discussion |
| --- | --- | --- | --- | --- | --- | --- |
| Age | Anderson et al. (2014) | Younger | Older | OR: 1.04 (95% CI: 1.03 to 1.06) | OR: 0.98 (95% CI: 0.93 to 1.04) | There was no difference in the likelihood of receiving recommended medication between older and younger people. Anderson et al. (2015) and Reynolds et al. (2020a) could not be included in this meta-analysis due to lack of data. Anderson et al. (2015) reported an adjusted OR=1.05 for increasing age. Reynolds et al. (2020a) reported no significant difference based on age. If these results were added in the meta-analysis, it is likely that the conclusion would remain as there not being significant difference based on age. |
|  | Callaghan et al. (2019) |  |  | aOR: 0.95 (95% CI: 0.94 to 0.97) |  |  |
|  | Chahine and Al Souheil (2021) |  |  | aOR: 0.96 (95% CI: 0.94 to 0.99) |  |  |
|  | Anderson et al. (2015) |  |  | OR: 1.05 (95% CI: NR to NR) | NA |  |
|  | Reynolds et al. (2020a) |  |  | NR & NS | NA |  |
|  | Marcianò et al. (2024) | 18-64 years | ≥ 65 years | OR: 0.98 (95% CI: 0.53 to 1.83) | NA |  |
| Sex | Anderson et al. (2014) | Female | Male | OR: 0.74 (95% CI: 0.56 to 0.99) | OR: 0.94 (95% CI: 0.79 to 1.10) | There was no difference in the likelihood of receiving recommended medication between males and females. |
|  | Callaghan et al. (2019) |  |  | aOR: 0.68 (95% CI: 0.62 to 0.74) |  |  |
|  | Reynolds et al. (2020a) |  |  | OR: 2.10 (95% CI: 1.14 to 3.88) |  |  |
|  | Chahine and Al Souheil (2021) |  |  | aOR: 2.17 (95% CI: 0.99 to 4.37) |  |  |
|  | Marcianò et al. (2024) |  |  | OR: 1.78 (95% CI: 0.92 to 3.46) |  |  |
|  | Nygvist et al. (2024) |  |  | OR: 0.83 (95% CI: 0.81 to 0.86) |  |  |
|  | Chen et al. (2011) |  |  | OR: 1.16 (95% CI: 1.05 to 1.28) |  |  |
| Ethnicity | Lakkad et al. (2023) | Black | White | aOR: 1.76 (95% CI: 1.43 to 2.18) | OR: 1.40 (95% CI: 1.18 to 1.66) | People with white ethnicity were more likely to receive recommended medication compared to people with other ethnicities.  Reynolds et al. (2020a) could not be included in this meta-analysis due to lack of data. Reynolds et al. (2020a) reported no significant difference based on ethnicity. It is unclear how their results might affect the meta-analysis, although their sample was small (n= 202) compared to the other studies (n=7,116 and n=14,426), so a meta-analysis with their results might not be much different to the current meta-analysis. |
|  |  | “Other” |  | aOR: 1.37 (95% CI: 1.07 to 1.75) |  |  |
|  | Callaghan et al. (2019) | Asian |  | aOR: 1.64 (95% CI: 1.22 to 2.17) |  |  |
|  |  | Hispanic |  | aOR: 1.32 (95% CI: 1.02 to 1.52) |  |  |
|  |  | Black |  | aOR: 1.11 (95% CI: 0.97 to 1.27) |  |  |
|  | Reynolds et al. (2020a) | Asian |  | NR & NS | NA |  |
|  |  | Black |  | NR & NS | NA |  |
|  |  | Other |  | NR & NS | NA |  |
| Geographic region | Lakkad et al. (2023) | West | Northeast | NS | NA | In the USA, there were no differences in the likelihood of receiving recommended medication between geographic regions or between urban and rural regions. |
|  |  |  | Midwest | NS | NA |  |
|  |  |  | South | NS | NA |  |
|  |  | Rural | Urban | NS | NA |  |
| Marital status | Lakkad et al. (2023) | Single, divorced, widowed | Married | OR: 1.05 (95% CI: 0.96 to 1.16) | NA | Marital status was not associated with any differences in the likelihood of receiving recommended medication. |
| Education | Lakkad et al. (2023) | No college education | College education | NS | NA | Generally, it seems that there were no differences in the likelihood of receiving recommended medication based on education. There is some evidence that people with a bachelor’s degree or higher education were less likely to receive recommended medication compared to people with below 12th grade education. |
|  | Callaghan et al. (2019) | Education (Less than 12^th^ grade) | High school diploma | NS | NA |  |
|  |  |  | Less than bachelor’s degree | NS | NA |  |
|  |  |  | Bachelor’s degree plus | aOR: 0.50 (95% CI: 0.29 to 0.87) | NA |  |
| Household income | Lakkad et al. (2023) | <$33K | SEVERAL | NS | NA | Generally, it seems that there were no differences in the likelihood of receiving recommended medication based on household income. There is some evidence that people with >$100K household income were less likely to receive recommended medication compared to people with <$40K household income. |
|  | Callaghan et al. (2019) | <$40K | >$100K | aOR: 0.86 (95% CI: 0.76 to 0.99) | NA |  |
|  |  |  | SEVERAL | NS | NA |  |
| Socioeconomic status | Anderson et al. (2014) | Lower Townsend index | Higher Townsend index | aOR: 1.12 (95% CI: 1.06 to 1.17) | NA | Living in a more deprived region was associated with a higher likelihood of receiving recommended medication. |
|  | Anderson et al. (2015) | Lower Townsend index | Higher Townsend index | aOR: 1.06 (95% CI: NR to NR) | NA |  |
| Insurance | Reynolds et al. (2020a) | Insurance (NR) | SEVERAL | NS | NA | There was no difference in the likelihood of receiving recommended medication based on insurance type. |
| Prescriber | Reynolds et al. (2020a) | Not resident | Resident | OR: 2.18 (95% CI: 1.12 to 5.66) | NA | People were more likely to receive recommended medication from a physician compared to a pharmacist.  People were more likely to receive recommended medication from a neurologist, physiatrist, or anaesthesiologist compared to other specialists.  Being treated by a resident was associated with a higher likelihood of receiving recommended medication. |
|  | Callaghan et al. (2019) | Not neurologist | Neurologist | aOR: 1.92 (95% CI: 1.75 to 2.10) | NA |  |
|  |  | Not physiatrist | Physiatrist | aOR: 1.55 (95% CI: 1.35 to 1.79) | NA |  |
|  |  | Not anaesthesiologist | Anaesthesiologist | aOR: 3.07 (95% CI: 2.60 to 3.61) | NA |  |
|  | Chahine and Al Souheil (2021) | Pharmacist | Physician | aOR: 23.33 (95% CI: 9.36 to 58.11) | NA |  |
| Prescribing process | Reynolds et al. (2020a) | No Smartset notification with best practise alert | Smartset notification with best practise alert | NS | NA | Smartset notification with best practise alert was not associated with any differences in the likelihood of receiving recommended medication compared to best practise alert alone. |
| NeuP diagnosis | # Gore et al. (2007a) | Mixed pain | Pure NeuP | OR: 1.37 (95% CI: 1.31 to 1.44) | NA | People with purely NeuP were more likely to receive recommended medication compared to people with mixed pain.  Hall et al. (2008) found that people with PHN were less likely to receive recommended medication compared to people with TN. Koopman et al. (2010) found no difference in the likelihood of receiving recommended medication between people with facial PHN and people with TN (their analysis included opioids).  DN > no DN  DN = PHN  *Udall et al. (2019) – people with PHN n=7, and they all had recommended medication. DN=39, 38.4% had recommended medication. Excluded from meta-analysis.  TN > DN  PLP = DN  DN > NeuP back pain  Overall, the results are mixed about DN. Generally, it’s either that people with DN are more likely to receive recommended medication or that there is no statistical difference. Only people with TN are more likely to receive recommended medication that people with DN. |
|  | Nygvist et al. (2024) | Lumbago with sciatica | Lumbar spinal stenosis | OR: 1.02 (95% CI: 0.97 to 1.08) | NA |  |
|  |  |  | Nerve root and plexus compressions in intervertebral disc  disorders | OR: 1.20 (95% CI: 1.16 to 1.25) | NA |  |
|  |  |  | Lumbar root canal stenosis | OR: 1.14 (95% CI: 1.04 to 1.23) | NA |  |
|  | Hall et al. (2008) | PHN | TN | OR: 1.26 (95% CI: 1.10 to 1.43) | NA |  |
|  | # Koopman et al. (2010) | PHN (first episode of facial pain) | TN (first episode of facial pain) | NS | NA |  |
|  |  | PHN (second episode of facial pain) | TN (second episode of facial pain) | NS | NA |  |
|  | Chahine and Al Souheil (2021) | No PHN | PHN | NS | NA |  |
|  |  | No cervical/ lumbar radiculopathy | Cervical/lumbar radiculopathy | NS | NA |  |
|  |  | No SCI | SCI | NS | NA |  |
|  |  | No neuropathic postoperative pain | Neuropathic postoperative pain | NS | NA |  |
|  |  | No TN | TN | NS | NA |  |
|  |  | No post traumatic neuralgia | Post traumatic neuralgia | NS | NA |  |
|  | Chahine and Al Souheil (2021) | No DN | DN | aOR: 6.27 (95% CI: 1.72 to 22.82) | NA |  |
|  | Hall et al. (2008) | PHN |  | OR: 0.96 (95% CI: 0.83 to 1.10) | OR: 1.20 (95% CI: 0.94 to 1.53) |  |
|  | Hall et al. (2013) |  |  | OR: 1.48 (95% CI: 1.37 to 1.59) |  |  |
|  | Margolis et al. (2017) |  |  | OR: 1.20 (95% CI: 1.12 to 1.29) |  |  |
|  | Udall et al. (2019) |  |  | OR: 0.042 (0.0022 to 0.793) * |  |  |
|  | Hall et al. (2008) | TN |  | OR: 0.76 (95% CI: 0.66 to 0.88) | NA |  |
|  |  | PLP |  | OR: 0.86 (95% CI: 0.50 to 1.47) | OR: 1.38 (95% CI: 0.59 to 3.20) |  |
|  | Hall et al. (2013) |  |  | OR: 2.04 (95% CI: 1.48 to 2.18) |  |  |
|  |  | NeuP back pain |  | OR: 2.01 (95% CI: 1.90 to 2.13) | NA |  |
|  | Udall et al. (2019) | NeuP back pain (n=77) |  | OR: 0.68 (95% CI: 0.31 to 1.47) | NA |  |
|  | Butler et al. (2018) | PTN |  | OR: 1.69 (95% CI: 0.96 to 2.94) | NA |  |
|  | Udall et al. (2019) | PTN (n=36) |  | OR: 0.59 (95% CI: 0.34 to 1.04) | NA |  |
|  |  | PSN (n=28) |  | OR: 0.25 (95% CI: 0.09 to 0.71) | NA |  |
|  |  | Central neuropathic pain (n=37) |  | OR: 0.66 (95% CI: 0.27 to 1.64) | NA |  |
| Previous pain medication | Lakkad et al. (2023) | No continuous opioid use | Continuous opioid use | aOR: 0.61 (95% CI: 0.52 to 0.71) | NA | People who have been using opioids continuously (>90 days), were less likely to receive recommended medication. The previous use of NSAIDs or long-acting opioids was not associated with any differences the likelihood of receiving recommended medication. |
|  |  | No NSAIDs | NSAIDs | NS | NA |  |
|  |  | No long-acting opioids | Long-acting opioids | NS | NA |  |
| Other previous medication | Lakkad et al. (2023) | No benzodiazepines | Benzodiazepines | aOR: 1.22 (95% CI: 1.03 to 1.45) | NA | People who had been using antipsychotics, benzodiazepines, or other central nervous system depressants were more likely to receive recommended medication compared to people with no history of using these medications. The previous use of muscle relaxants or steroids was not associated with any differences the likelihood of receiving recommended medication. |
|  |  | No antipsychotics | Antipsychotics | aOR: 1.89 (95% CI: 1.36 to 2.63) | NA |  |
|  |  | No non-benzodiazepine CNS depressants | Non-benzodiazepine CNS depressants | aOR: 1.27 (95% CI: 1.08 to 1.50) | NA |  |
|  |  | No steroids | Steroids | NS | NA |  |
|  |  | No muscle relaxants | Muscle relaxants | NS | NA |  |
| Diabetes | Lakkad et al. (2023) | No diabetes | Diabetes | aOR: 1.32 (95% CI: 1.18 to 1.47) | OR: 1.41 (95% CI: 1.23 to 1.62) | People with diabetes were more likely to receive recommended medication compared to people without diabetes. |
|  | Callaghan et al. (2019) |  |  | aOR: 1.52 (95% CI: 1.40 to 1.66) |  |  |
|  | Chahine and Al Souheil (2021) |  |  | aOR: 0.75 (95% CI: 0.24 to 2.33) |  |  |
| Mental health disorder | Lakkad et al. (2023) | No mental health disorder | Mental health disorder | aOR: 3.57 (95% CI: 3.12 to 4.09) | NA | People with a mental health disorder were more likely to receive recommended medication compared to people without a mental health disorder. This was also true for those with anxiety and/or depression or depression.  There was no difference in the likelihood of receiving recommended medication between people with or without anxiety (although the results were mixed). |
|  | # Boulanger et al. (2009) | No anxiety and/or depression (commercial insurance group) | Anxiety and/or depression (commercial insurance group) | OR: 3.81 (95% CI: 3.17 to 4.59) | OR: 2.17 to (95% CI: 1.69 to 2.78) |  |
|  |  | No anxiety and/or depression (Medicare supplemental group) | Anxiety and/or depression (Medicare supplemental group) | OR: 1.48 (95% CI: 1.31 to 1.67) |  |  |
|  | Callaghan et al. (2019) | No anxiety | Anxiety | aOR: 1.54 (95% CI: 1.33 to 1.79) | OR: 1.03 (95% CI: 0.74 to 1.44) |  |
|  | Chahine and Al Souheil (2021) |  |  | aOR: 0.34 (95% CI: 0.09 to 1.28) |  |  |
|  | Chen et al. (2011) |  |  | OR: 0.76 (95% CI: 0.61 to 0.94) |  |  |
|  | Callaghan et al. (2019) | No depression | Depression | aOR: 1.65 (95%CI: 1.35 to 2.02) | OR: 1.62 (95% CI: 1.33 to 1.98) |  |
|  | Chahine and Al Souheil (2021) |  |  | aOR: 1.16 (95% CI: 0.47 to 2.88) |  |  |
| Other comorbidities | Lakkad et al. (2023) | No hemiplegia | Hemiplegia | aOR: 1.93 (95% CI: 1.03 to 3.60) | NA | In people with neuropathic pain related to breast cancer treatment, comorbidities that were associated with a higher likelihood of receiving recommended first-line medication included: mental health conditions, hemiplegia, dementia, diabetes, and rheumatologic disease.  In people with neuropathic pain related to breast cancer, comorbidities that were not associated with any differences the likelihood of receiving recommended first-line medication included: substance use disorder, non-malignant pain conditions, myocardial infraction, stroke, congestive heart failure, peptic ulcer, peripheral vascular disease, chronic obstructive pulmonary disease, and liver disease.  Higher Charlson score was associated with a higher likelihood of receiving recommended medication. |
|  |  | No dementia | Dementia | aOR: 1.68 (95% CI: 1.23 to 2.31) | NA |  |
|  |  | No rheumatologic disease | Rheumatologic disease | OR: 0.79 (95% CI: 0.63 to 0.98) | NA |  |
|  |  | No substance use disorder | Substance use disorder | NS | NA |  |
|  |  | No myocardial infraction | Myocardial infraction | NS | NA |  |
|  |  | No stroke | Stroke | NS | NA |  |
|  |  | No congestive heart failure | Congestive heart failure | NS | NA |  |
|  |  | No peptic ulcer | Peptic ulcer | NS | NA |  |
|  |  | No peripheral vascular disease | Peripheral vascular disease | NS | NA |  |
|  |  | No chronic obstructive pulmonary disease | Chronic obstructive pulmonary disease | NS | NA |  |
|  |  | No liver disease | Liver disease | NS | NA |  |
|  | Callaghan et al. (2019) | Lower Charlson score | Higher Charlson score | aOR: 1.16 (95% CI: 1.11 to 1.21) | NA |  |
| Breast-cancer related variables | Lakkad et al. (2023) | No radiation | Radiation | NS | NA | There was no difference the likelihood of receiving recommended medication as first choice based on breast cancer treatment; age of diagnosis; Surveillance, Epidemiology, and End Results (SEER) grade; SEER summary stage; or hormone receptor status.  People diagnosed with breast cancer during 2013 were more likely to receive recommended first-line treatment for NeuP compared to people diagnosed with breast cancer during 2007. However, there were no significant differences in the likelihood of receiving recommended first-line treatment for NeuP between people diagnosed during the years 2008 to 2012 compared to people diagnosed with breast cancer during 2007. |
|  |  | No targeted therapy | Targeted therapy | NS | NA |  |
|  |  | Higher age at cancer diagnosis | Lower age at cancer diagnosis | NS | NA |  |
|  |  | Hormone receptor status | SEVERAL | NS | NA |  |
|  |  | Breast cancer year of diagnosis (2007) | 2013 | OR: 1.29 (95% CI: 1.04 to 1.60) | NA |  |
|  |  |  | SEVERAL | NS | NA |  |
|  |  | SEER grade | SEVERAL | NS | NA |  |
|  |  | SEER summary stage | SEVERAL | NS | NA |  |
| Laboratory measurements | Anderson et al. (2014) | Higher eGFR | Lower eGFR | OR: 1.02 (95% CI: NR to NR) | NA | Characteristics that are associated with a higher likelihood of receiving recommended medication included: lower eGFR, lower LDL.  Characteristics that were not associated with any difference in the likelihood of receiving recommended medication included: diabetes duration, low/high systolic blood pressure, low/high diastolic blood pressure, high/low HDL cholesterol, high/low total cholesterol, high/low HbA1c. |
|  |  | Higher LDL | Lower LDL | With medication 2.3 (2.2 to 2.4) mmol/l. Without medication 2.5 (2.4 to 2.5) mmol/l. | NA |  |
|  |  | Lower HDL | Higher HDL | NS | NA |  |
|  |  | Lower total cholesterol | Higher total cholesterol | NS | NA |  |
|  |  | Shorter diabetes duration | Longer diabetes duration | NS | NA |  |
|  |  | Lower HbA1C | Higher HbA1C | NS | NA |  |
|  |  | Lower systolic blood pressure | Higher systolic blood pressure | NS | NA |  |
|  |  | Lower diastolic blood pressure | Higher diastolic blood pressure | NS | NA |  |
|  | Anderson et al. (2015) | Higher eGFR | Lower eGFR | OR: 0.97 (95% CI: NR to NR) | NA |  |
|  |  | Lower systolic blood pressure | Higher systolic blood pressure | OR: 0.97 (95% CI: NR to NR) | NA |  |
| BMI | Anderson et al. (2014) | Lower BMI | Higher BMI | OR: 1.05 (95% CI: 1.03 to 1.07) | NA | People with higher BMI were more likely to receive recommended medication. |
|  | Anderson et al. (2015) |  |  | OR: 1.03 (95% CI: NR to NR) | NA |  |
|  | Marcianò et al. (2024) | <25 (normal weight & underweight) | 25-30 (Overweight) | OR: 2.40 (95% CI: 1.14 to 5.06) | NA |  |
|  |  |  | ≥30 (Obese) | OR: 3.36 (95% CI: 1.48 to 7.65) | NA |  |
| Alcohol | Chahine and Al Souheil (2021) | Lower alcohol intake | Higher alcohol intake | aOR: 0.41 | NA | Lower alcohol intake was associated with receiving recommended medication. |
| Smoking | Chahine and Al Souheil (2021) | Non-smoker | Smoker | NS | NA | There was no difference in the likelihood of receiving recommended medication between smokers and non-smokers. |

Table S15. Analysis of predictors of being prescribed opioids as the first choice. The results of this table should be complemented with the results from Lakkad et al. (2023) and Nygvist et al. (2024) in Table S13, as their results reflect predictors for NOT being prescribed opioids as the first choice. Adjusted odds ratios are provided for variables that were significant in multivariable analyses where applicable. Statistically non-significant results are provided when they were needed for meta-analysis. MA: meta-analysis, OR: odds ratio, aOR: adjusted odds ratio, NS: not significant, NA: not applicable.

| Variable | Study | Reference | Comparison | Result | MA | Discussion |
| --- | --- | --- | --- | --- | --- | --- |
| Age | Patil et al. (2015) | Age (18-40 years) | SEVERAL | NS | NA | There was no difference in the likelihood of receiving opioids as first-line medication based on age. |
| Sex | Patil et al. (2015) | Female | Male | NS | NA | There was no difference in the likelihood of receiving opioids as first-line medication between males and females. |
| Geographic region | Patil et al. (2015) | West USA | Midwest USA | aOR: 0.49 (95% CI: 0.24 to 0.99) | OR: 0.74 (95% CI: 0.01 to 41.96). Fixed effects OR: 0.92 (95% CI: 0.80 to 1.05). | There was no difference in the likelihood of receiving opioids as first-line medication between people living in the Midwest USA and West USA.  In the USA, there were no differences in the likelihood of receiving recommended first-line medication between geographic regions. |
|  | Lakkad et al. (2023) |  |  | OR: 0.94 (95% CI: 0.82 to 1.08) |  |  |
|  | Patil et al. (2015) |  | South USA | aOR: 0.52 (95% CI: 0.24to 1.12) | OR: 0.81(95% CI: 0.02 to37.37). Fixed effects OR: 0.98 (95% CI: 0.86 to 1.11). |  |
|  | Lakkad et al. (2023) |  |  | OR: 0.99 (95% CI: 0.87 to 1.13) |  |  |
|  | Patil et al. (2015) | South USA | Midwest USA | aOR: 0.93 (95% CI: 0.50 to 1.74) | OR: 0.95 (95% CI: 0.85 to 1.06) |  |
|  | Lakkad et al. (2023) |  |  | OR: 0.95 (95% CI: 0.84 to 1.06) |  |  |
| Other medication | Patil et al. (2015) | No insulin | Insulin | NS | NA | There was no difference in the likelihood of receiving opioids as first-line medication between people who used insulin and people who did not use insulin. |
| Prescriber | Patil et al. (2015) | Prescriber (GP/FP/intern) | SEVERAL | NS | NA | Prescriber speciality/training level was no associated with any differences in the likelihood of receiving opioids as first-line medication. |
| NeuP diagnosis year | Patil et al. (2015) | 2001-2002 | 2003-2004 | NS | NA | People diagnosed with NeuP between 2008 to 2011 were less likely to receive opioids than people diagnosed between 2002 to 2005. |
|  |  |  | 2005-2006 | NS | NA |  |
|  |  |  | 2007-2008 | NS | NA |  |
|  | Reed et al. (2023) | 2002-2005 | 2005-2008 | NS | NA |  |
|  |  |  | 2008-2011 | OR: 0.68 (95% CI: 0.55 to 0.85) | NA |  |
| NeuP diagnosis | Hall et al. (2008) | DN | PHN | OR: 1.38 (95% CI: 1.13 to 1.68) | OR: 1.70 (95% CI: 1.15 to 2.51) | People with PHN were more likely to receive opioids as first-line medication compared to people with DN.  There was no difference in the likelihood of receiving opioids as first-line medication between people who have PLP and people who have PHN.  People with PLP were more likely to receive opioids as first-line medication compared to people with DN.  People with TN were less likely to receive opioids as first choice compared to other NeuP diagnoses.  People with NeuP back pain were more likely to receive opioids as first choice compared to other NeuP diagnoses. |
|  | Hall et al. (2013) |  |  | OR: 2.05 (95% CI: 1.86 to 2.27) |  |  |
|  | Hall et al. (2008) | PLP |  | OR: 1.67 (95% CI: 0.90 to 3.09) | OR: 1.04 (95% CI: 0.44 to 2.45) |  |
|  | Hall et al. (2013) |  |  | OR: 0.70 (95% CI: 0.47 to 1.04) |  |  |
|  | Hall et al. (2008) | DN | PLP | OR: 2.30 (95% CI: 1.23 to 4.29) | OR: 1.70 (95% CI: 1.08 to 2.68) |  |
|  | Hall et al. (2013) |  |  | OR: 1.43 (95% CI: 0.95 to 2.14) |  |  |
|  | Hall et al. (2008) | PHN | TN | OR: 0.53 (95% CI: 0.43 to 0.66) | OR: 0.55 (95% CI: 0.41 to 0.74) |  |
|  |  | PLP |  | OR: 0.32 (95% CI: 0.18 to 0.57) |  |  |
|  |  | DN |  | OR: 0.73 (95% CI: 0.56 to 0.95) |  |  |
|  | Hall et al. (2013) | DN | NeuP back pain | OR: 6.67 (95% CI: 7.14 to 6.25) | OR: 4.55 (95% CI: 4.35 to 4.75) |  |
|  |  | PLP |  | OR: 3.33 (95% CI: 2.44 to 4.55) |  |  |
|  |  | PHN |  | OR: 3.49 (95% CI: 3.23 to 3.70) |  |  |
| Comorbidities | Patil et al. (2015) | No diabetes with organ damage | Diabetes with organ damage | aOR: 9.08 (95% CI: 1.06 to 77.58) | NA | In people with DN, having organ damage or nephropathy increases the likelihood of receiving opioids as first-line treatment. In people with DN, comorbidities that did not affect the likelihood of receiving opioids as first-line treatment included: cerebrovascular and peripheral vascular diseases, metabolic disorders, obesity, and retinopathy. In people with DN, Charlson comorbidity score does not affect the likelihood of receiving opioids as first-line treatment. |
|  |  | No nephropathy | Nephropathy | aOR: 13.25 (95% CI: 1.89 to 92.69) | NA |  |
|  |  | No cerebrovascular and/or peripheral vascular disease | Cerebrovascular and/or peripheral vascular disease | NS | NA |  |
|  |  | No metabolic disorder | Metabolic disorder | NS | NA |  |
|  |  | No retinopathy | Retinopathy | NS | NA |  |
|  |  | No cardiovascular disease | Cardiovascular disease | NS | NA |  |
|  |  | Charlson score (0) | SEVERAL | NS | NA |  |
|  |  | No obesity | Obesity | NS | NA |  |

Table S16. Analysis of predictors of being prescribed opioids at any point. This table includes content of Table S15 and provides additional results that were not relevant to table S15. The results of this table should be complemented with the results from Lakkad et al. (2023) and Nygvist et al. (2024) in Table S13, as their results reflect predictors for NOT being prescribed opioids. Adjusted odds ratios are provided for variables that were significant in multivariable analyses where applicable. Statistically non-significant results are provided when they were needed for meta-analysis. MA: meta-analysis, OR: odds ratio, aOR: adjusted odds ratio, NS: not significant, NA: not applicable.

| Variable | Study | Reference | Comparison | Result | MA | Discussion |
| --- | --- | --- | --- | --- | --- | --- |
| Age | Chahine and Al Souheil (2021) | Younger | Older | aOR: 0.98 (95% CI: 0.95 to 1.01) | OR: 0.92 (95% CI: 0.82 to 1.04) | There was no difference in the likelihood of receiving opioids between “older” and “younger” people based on the results from Chahine and Al Souheil (2021) and Callaghan et al. (2019). Patil et al. (2015) also found no differences between several age groups.  Overall, there was no difference in the likelihood of receiving opioids between people who were 75-84 years old and people who were 85 years old or older. However, Johnston et al. (2014) found significant results based on NeuP diagnosis. In people with DN, those aged 75-84 years were more likely to receive opioids than those aged 85 years old or older. In people with PHN, those aged 85 years old or older were more likely to receive opioids than those aged 75-84 years.  Meta-analysis based on Johnson et al. and Goswami et al. found that people aged 65-74 years were less likely to receive opioids than those aged 85 years old or older. Based on the results from Johnston et al. (2014), this was true for people with PHN whereas in people with DN, those aged 65-74 years were more likely to receive opioids than those aged 85 years or older. The results from Lin et al. are not directly comparable to this due to different reference category, but their results suggest opposite pattern.  It should be noted that Goswami et al. (2023) only compared groups that received opioids, or gabapentin, or both.  Meta-analysis for Johnston et al. (2014) data only:  >85 years vs 75-84 years:  Random effects OR: 0.80 (95% CI: 0.00 to 195.63).  Fixed effects OR: 0.89 (95% CI: 0.83 to 0.96)  >85 years vs 65-74 years  Random effects OR: 0.65 (95% CI: 0.00 to 6392.50).  Fixed effects OR: 0.82 (95% CI: 0.76 to 0.88) |
|  | Callaghan et al. (2019) |  |  | aOR: 0.87 (95% CI: 0.86 to 0.89) |  |  |
|  | Johnston et al. (2014) | ≥85 years (DN) | 75-84 years (DN) | OR: 1.23 (95% CI: 1.13 to 1.35) | OR: 0.88 (95% CI: 0.75 to 1.03) |  |
|  |  | ≥85 years (PHN) | 75-84 years (PHN) | OR: 0.52 (95% CI: 0.46 to 0.58) |  |  |
|  | Goswami et al. (2023) | ≥85 years | 75-84 years | OR: 1.03 (95% CI: 0.83 to 1.28) |  |  |
|  | Johnston et al. (2014) | ≥85 years (DN) | 65-74 years (DN) | OR: 1.34 (95% CI: 1.22 to 1.46) | OR: 0.75 (95% CI: 0.62 to 0.90) |  |
|  |  | ≥85 years (PHN) | 65-74 years (PHN) | OR: 0.31 (95% CI: 0.28 to 0.35) |  |  |
|  | Goswami et al. (2023) | ≥85 years | 65-74 years | OR: 0.97 (95% CI: 0.79 to 1.19) |  |  |
|  | Patil et al. (2015) | 18-40 years | 41-54 years | NS | NA |  |
|  |  |  | 55-64 years | NS | NA |  |
|  |  |  | 65-74 years | NS | NA |  |
|  |  |  | ≥75 years | NS | NA |  |
|  | Lin et al. (2023) | <65 years | 65-74 years | NS | NA |  |
|  |  |  | 75-84 years | aOR: 0.66 (95% CI: 0.50 to 0.86) | NA |  |
|  |  |  | ≥85 years | aOR: 0.50 (95% CI: 0.36 to 0.69) | NA |  |
|  | Marcianò et al. (2024) | 18-64 years (stratified by sex) | ≥65 years (stratified by sex) | NS | NA |  |
| Sex | Chahine and Al Souheil (2021) | Female | Male | OR: 1.36 (95% CI: 0.61 to 3.02) | OR: 1.08 (95% CI: 0.87 to 1.34) | There was no difference in the likelihood of receiving opioids between males and females. |
|  | Callaghan et al. (2019) |  |  | OR: 1.12 (95% CI: 1.03 to 1.22) |  |  |
|  | Goswami et al. (2023) |  |  | OR: 1.52 (95% CI: 0.87 to 2.66) |  |  |
|  | Patil et al. (2015) |  |  | OR: 0.92 (95% CI: 0.81 to 1.06) |  |  |
|  | Lin et al. (2023) |  |  | OR: 0.94 (95% CI: 0.81 to 1.10) |  |  |
|  | Marcianò et al. (2024) |  |  | NS (Reported for individual opioids) | NA |  |
| Ethnicity | Callaghan et al. (2019) | White | Black | aOR: 1.02 (95% CI: 0.89 to 1.16) | OR: 0.81 (95% CI: 0.66 to 1.00) | Lakkad et al. (2023) found that people with white ethnicity were less likely to receive opioids as the first choice compared to people from non-white ethnicities. Callaghan et al. (2019) and Goswami et al. (2023) found that people with white ethnicity were more likely to receive opioids (not limited to as first choice). These contrasting results could be explained by that people with white ethnicity may be more likely to receive any pain medication, which can be true at the same time with people with white ethnicity being more likely to receive recommended medication rather than opioids as first choice.  Overall, the meta-analysis suggests that there was no difference in the likelihood of receiving opioids between people with white ethnicity and people with non-white ethnicity. However, excluding the results from Lakkad et al. (2023) which analysed the likelihood of not receiving opioids as first choice, the result would be:  OR: 0.69 (95% CI: 0.58 to 0.83)  It should be noted that Goswami et al. (2023) only compared groups that received opioids, or gabapentin, or both. Also, the results from Goswami et al. (2023) come from univariate analysis while the other studies reported multivariate ORs. |
|  |  |  | Asian | aOR: 0.57 (95% CI: 0.45 to 0.73) |  |  |
|  |  |  | Hispanic | aOR: 0.87 (95% CI: 0.75 to 1.00) |  |  |
|  | Goswami et al. (2023) |  | Black | OR: 0.60 (95% CI: 0.49 to 0.73) |  |  |
|  |  |  | Asian | OR: 0.54 (95% CI: 0.38 to 0.77) |  |  |
|  |  |  | Hispanic | OR: 0.51 (95% CI: 0.39 to 0.66) |  |  |
|  | Lakkad et al. (2023) |  | Black | aOR: 1.76 (95% CI: 1.43 to 2.18) |  |  |
|  |  |  | “Other” | aOR: 1.37 (95% CI: 1.07 to 1.75) |  |  |
|  | Lin et al. (2023) |  | Black | OR: 0.62 (95% CI: 0.43 to 0.87) |  |  |
|  |  |  | Hispanic | OR: 0.87 (95% CI: 0.62 to 1.23) |  |  |
|  |  |  | Asian | OR:0.68 (95% CI: 0.42 to 1.11) |  |  |
|  |  |  | Other/missing | OR: 0.70 (95% CI: 0.36 to 1.39) |  |  |
| Geographic region | Goswami et al. (2023) | West USA | Midwest USA | OR: 1.26 (95% IC: 1.01 to 1.56) | OR: 0.92 (95% CI: 0.32 to 2.64) | There was no difference in the likelihood of receiving opioids between people living in urban and rural locations, or by geographic location. |
|  | Patil et al. (2015) |  |  | OR: 0.49 (95% CI: 0.24 to 0.99) |  |  |
|  | Lakkad et al. (2023) |  |  | OR: 0.94 (95% CI: 0.82 to 1.08) |  |  |
|  | Goswami et al. (2023) |  | South USA | OR: 1.19 (95% CI: 0.99 to 1.45) | OR: 1.03 (95% CI: 0.56 to 1.89) |  |
|  | Patil et al. (2015) |  |  | OR: 0.52 (95% CI: 0.24 to 1.12) |  |  |
|  | Lakkad et al. (2023) |  |  | OR: 1.19 (95% CI: 0.99 to 1.45) |  |  |
|  | Lakkad et al. (2023) |  | Northeast USA | OR: 0.99 (95% CI: 0.87 to 1.13) | OR: 0.86 (95% CI: 0.65 to 1.13) |  |
|  | Lin et al. (2023) |  |  | aOR: 0.61 (95% CI: 0.44 to 0.70) |  |  |
|  | Goswami et al. (2023) |  |  | OR: 0.95 (95% CI: 0.80 to 1.13) |  |  |
|  | Goswami et al. (2023) | Midwest USA | South USA | OR: 1.05 (95% CI: 0.87 to 1.27) | OR: 0.97 (95% CI: 0.88 to 1.07) |  |
|  | Patil et al. (2015) |  |  | OR: 0.93 (95% CI: 0.50 to 1.74) |  |  |
|  | Lakkad et al. (2023) |  |  | OR: 0.95 (95% CI: 0.84 to 1.06) |  |  |
|  | Lin et al. (2023) | Northeast USA | Midwest USA | aOR: 1.47 (95% CI: 1.08 to 1.99) | NA |  |
|  |  |  | South USA | aOR:1.87 (95% CI: 1.43 to 2.46) | NA |  |
|  | Lin et al. (2023) | Urban | Rural | aOR: 0.95 (95% CI: 0.77 to 1.16) | OR: 0.95 (95% CI: 0.75 to 1.20) |  |
|  | Lakkad et al. (2023) |  |  | OR: 0.97 (95% CI: 0.58 to 1.09) |  |  |
| Education | Callaghan et al. (2019) | Education (less than 12^th^ grade) | SEVERAL | NS | NA | There was no difference in the likelihood of receiving opioids between people with high or low education. |
| Household income | Callaghan et al. (2019) | <$40K | $40K-$49K | OR: 1.05 (95% CI: 0.89 to 1.23) | NA | It is unclear how household income may be associated with the likelihood of receiving opioids. ($75K-$99K was the only significant category compared to >$40K) |
|  |  |  | $55K-$59K | OR: 1.03 (95% CI: 0.87 to 1.21) | NA |  |
|  |  |  | $60K-$74K | OR: 0.95 (95% CI: 0.82 to 1.10) | NA |  |
|  |  |  | $75K-$99K | OR:1.21 (95% CI: 1.05 to 1.39) | NA |  |
|  |  |  | >$100K | OR: 1.02 (95% CI: 0.89 to 1.16) | NA |  |
| Insurance | Patil et al. (2015) | NA | SEVERAL | NS | NA | Generally, it seems that the type of insurance was not associated with any difference in the likelihood of receiving opioids, but there are some results that suggest that there could be differences (the trends are unclear). |
|  | Callaghan et al. (2019) | PPO | “Other” | OR: 0.68 (95% CI: 0.51 to 0.90) | NA |  |
|  |  |  | SEVERAL | NS | NA |  |
| Prescriber | Chahine and Al Souheil (2021) | Pharmacist | Physician | aOR: 5.17 (95% CI: 1.02 to 25.95) | NA | People with NeuP were more likely to receive opioids from pain specialists (anaesthesiologists, neurologist, or physiatrists) compared to non-pain specialists. People with NeuP were more likely to receive opioids from physicians than pharmacists. |
|  | Callaghan et al. (2019) | Not neurologist | Neurologist | aOR: 1.23 (95% CI: 1.12 to 1.36) | NA |  |
|  |  | Not physiatrist | Physiatrist | aOR: 1.97 (95% CI: 1.67 to 2.34) | NA |  |
|  |  | Not anaesthesiologist | Anaesthesiologist | aOR: 3.90 (95% CI: 3.07 to 4.95) | NA |  |
|  | Patil et al. (2015) | Prescriber (GP/FP/intern) | SEVERAL | NS | NA |  |
| Prescribing process | Reynolds et al. (2020a) | No smartset notification with best practise alert | Smartset notification with best practise alert | NS | NA | There was no difference in the likelihood of receiving opioids with the use of Smartset notification. |
| Brand-name medication | Sicras-Mainar et al. (2015) | Generic name gabapentin | Brand name gabapentin | NS | NA | There was no difference in opioid use before and after brand-name gabapentin or generic gabapentin. People using generic pregabalin were more likely to have concomitant opioids compared to people using brand-name pregabalin. |
|  | Sicras-Mainar et al. (2019) | Generic name pregabalin | Brand name pregabalin | OR: 0.80 (95% CI: 0.69 to 0.94) | NA |  |
| Other NeuP medication | Gore et al. (2007b) | Before gabapentin | After gabapentin | OR: 1.33 (95% CI: 0.82 to 2.13) | OR: 1.24 (95% CI: 1.06 to 1.45) | There was no difference in the likelihood of receiving opioids before and after receiving a gabapentin prescription.  There was no difference in the likelihood of receiving opioids before and after receiving a pregabalin prescription.  People with low duloxetine compliance before opioid use were more likely to receive opioids for NeuP compared to people with high duloxetine compliance before opioid use.  People with duloxetine rather than other standard of care medications were less likely to receive opioids. |
|  | Berger et al. (2003) |  |  | OR: 0.31 (95% CI: 0.10 to 0.95) |  |  |
|  | Gore et al. (2011b) |  |  | OR: 1.27 (95% CI: 1.08 to 1.50) |  |  |
|  | Gore et al. (2007b) | Before pregabalin | After pregabalin | OR: 0.49 (95% CI: 0.27 to 0.89) | OR: 0.91 (95% CI: 0.60 to 1.37) |  |
|  | Gore et al. (2011a) |  |  | OR: 1.06 (95% CI: 0.85 to 1.31) |  |  |
|  | Gore et al. (2011b) |  |  | OR: 1.11 (95% CI: 0.94 to 1.31) |  |  |
|  | Wu et al. (2009) | Low duloxetine compliance (commercial insurance) | High duloxetine compliance (commercial insurance) | OR: 1.18 (95% CI: 0.91 to 1.53) | OR: 1.25 (95% CI: 1.03 to 1.51) |  |
|  | Wu et al. (2009) | Low duloxetine compliance (Medicare supplemental) | High duloxetine compliance (Medicare supplemental) | OR: 1.33 (95% CI: 1.00 to 1.76) |  |  |
|  | Wu et al. (2011) | Duloxetine | Any other stand of care NeuP medication | OR: 2.79 (95% CI: 1.83 to 4.25) | OR: 3.36 (95% CI: 2.33 to 4.84) |  |
|  | Chen et al. (2010a) |  |  | OR: 5.05 (95% CI: 2.67 to 9.53) |  |  |
|  | Yeh et al. (2021) | Pregabalin without dose titration | Pregabalin with dose titration | NS | NA |  |
|  | Zhao et al. (2011) | Duloxetine | Pregabalin | NS | NA |  |
|  | Lin et al. (2023) | No NeuP medication | NeuP medication | OR: 0.67 (95% CI: 0.57 to 0.80) | NA |  |
| Other medication | Patil et al. (2015) | No insulin | Insulin | NS | NA | There was no difference in the likelihood of receiving opioids as first-line medication between people who used insulin and people who did not use insulin. |
| Prescriber | Patil et al. (2015) | Prescriber (GP/FP/intern) | SEVERAL | NS | NA | Prescriber speciality/training level was no associated with any differences in the likelihood of receiving opioids as first-line medication. |
| Pain severity | Knoerl et al. (2024) | Mild | Moderate | NS | NA | There was no difference in the likelihood of receiving opioids based on pain severity. |
|  |  |  | Severe | NS | NA |  |
| NeuP diagnosis year | Patil et al. (2015) | 2001-2002 | 2003-2004 | NS | NA | People diagnosed with NeuP between 2008 to 2011 were less likely to receive opioids than people diagnosed between 2002 to 2005.  People diagnosed with NeuP between 2013 to 2017 were less likely to receive opioids than people diagnosed in 2010. |
|  |  |  | 2005-2006 | NS | NA |  |
|  |  |  | 2007-2008 | NS | NA |  |
|  | Reed et al. (2023) | 2002-2005 | 2005-2008 | NS | NA |  |
|  |  |  | 2008-2011 | OR: 0.68 (95% CI: 0.55 to 0.85) | NA |  |
|  | Lin et al. (2023) | 2010 | 2011 | NS | NA |  |
|  |  |  | 2012 | NS | NA |  |
|  |  |  | 2013 | OR: 0.59 (95% CI: 0.44 to 0.79) | NA |  |
|  |  |  | 2014 | OR: 0.52 (95% CI: 0.39 to 0.71) | NA |  |
|  |  |  | 2015 | OR: 0.32 (95% CI: 0.23 to 0.45) | NA |  |
|  |  |  | 2016 | OR: 0.24 (95% CI: 0.16 to 0.35) | NA |  |
|  |  |  | 2017 | OR: 0.21 (95% CI: 0.14 to 0.30) | NA |  |
| NeuP diagnosis | Johnston et al. (2014) | PHN | DN | OR: 1.20 (95% CI: 1.12 to 1.28) | OR: 1.43 (95% CI: 1.02 to 2.02) | People with PHN were more likely to receive opioids compared to people with DN. Dieleman et al. (2008) was excluded from this meta-analysis because their samples sizes were unclear. In their study 7.1% of people with DN received opioids and 15.3% of people with PHN received opioids, which supports the trend observed in the meta-analysis.  There was no difference in the likelihood of receiving opioids between people who have PLP and people who have PHN.  There was no difference in the likelihood of receiving opioids between people who have PLP and people who have PHN.  People with PLP were more likely to receive opioids compared to people with DN.  According to the meta-analysis, there was no difference in the likelihood of receiving opioids between people who have NeuP back pain and people have DN. However, each study alone supported that people with neuropathic back pain are more likely to receive opioids compared to people with DN.  People with NeuP back pain were more likely to receive opioids for NeuP compared to people with PHN.  Having cervical/lumbar radiculopathy was associated with a lower likelihood of receiving opioids.  The results about mixed pain and pure NeuP were mixed. |
|  | Udall et al. (2019) |  |  | OR: 0.92 (95% CI: 0.09 to 9.04) |  |  |
|  | Hall et al. (2008) |  |  | OR: 1.38 (95% CI: 1.13 to 1.68) |  |  |
|  | Hall et al. (2013) |  |  | OR: 1.83 (95% CI: 1.65 to 2.02) |  |  |
|  | Dieleman et al. (2008) |  |  | 15.3% vs 7.1% (sample size unclear) | NA |  |
|  | Hall et al. (2008) | PLP | DN | OR: 2.30 (95% CI: 1.23 to 4.29) | OR: 1.70 (95% CI: 1.08 to 2.68) |  |
|  | Hall et al. (2013) |  |  | OR: 1.43 (95% CI: 0.95 to 2.14) |  |  |
|  | Hall et al. (2008) | PHN | PLP | OR: 1.67 (95% CI: 0.90 to 3.09) | OR: 1.04 (95% CI: 0.44 to 2.45) |  |
|  | Hall et al. (2013) |  |  | OR: 0.70 (95% CI: 0.47 to 1.04) |  |  |
|  | Hall et al. (2008) | PHN | TN | OR: 0.53 (95% CI: 0.43 to 0.66) | NA |  |
|  |  | PLP |  | OR: 0.32 (95% CI: 0.18 to 0.57) | NA |  |
|  |  | DN |  | OR: 0.73 (95% CI: 0.56 to 0.95) | NA |  |
|  | Hall et al. (2013) | NeuP back pain | DN | OR: 7.38 (95% CI: 6.81 to 7.99) | OR: 5.18 (95% CI: 0.01 to 1924.33). Fixed effects model OR: 7.34 (95% CI: 6.78 to 7.95) |  |
|  | Udall et al. (2019) |  |  | OR: 2.80 (95% CI: 1.04 to 7.54) |  |  |
|  | Hall et al. (2013) |  | PHN | OR: 4.24 (95% CI: 3.98 to 4.52) | OR: 4.24 (95% CI: 3.59 to 5.01) |  |
|  | Udall et al. (2019) |  |  | OR: 7.72 (95% CI: 0.42 to 140.39) |  |  |
|  |  |  | PSN | NS | NA |  |
|  |  |  | PTN | NS | NA |  |
|  | Hall et al. (2013) |  | PLP | OR: 3.33 (95% CI: 2.44 to 4.55) | NA |  |
|  | Chahine and Al Souheil (2021) | No DN | DN | aOR: 0.05 (95% CI: 0.01 to 0.62) | NA |  |
|  |  | No PHN | PHN | NS | NA |  |
|  |  | No cervical/ lumbar radiculopathy | Cervical/lumbar radiculopathy | aOR: 0.05 (95% CI: 0.01 to 0.34) | NA |  |
|  |  | No SCI | SCI | NS | NA |  |
|  |  | No neuropathic postoperative pain | Neuropathic postoperative pain | NS | NA |  |
|  |  | No TN | TN | NS | NA |  |
|  |  | No post traumatic neuralgia | Post traumatic neuralgia | NS | NA |  |
|  | Dieleman et al. (2008) | DN | Mononeuropathy | 6.8% vs 7.1% (sample size unclear) | NA |  |
|  |  |  | Facial NeuP | 6.8% vs 7.1% (sample size unclear) | NA |  |
|  |  |  | CTS | 3.7% vs 7.1% (sample size unclear) | NA |  |
|  | Gore et al. (2007a) | Mixed pain | Pure NeuP | OR: 0.56 (95% CI: 0.52 to 0.60) | OR: 0.59 (95% CI: 0.55 to 0.63) |  |
|  | Gustavson et al. (2013) |  |  | OR: 0.78 (95% CI: 0.66 to 0.93) |  |  |
|  | Butler et al. (2018) | PTN (Visit 1) | DN (Visit 1) | OR: 3.32 (95% CI: 1.61 to 6.83) | OR: 3.84 (95% CI: 2.48 to 5.94) |  |
|  |  | PTN (Visit 3) | DN (Visit 3) | OR: 3.22 (95% CI: 1.53 to 6.77) |  |  |
|  |  | PTN (Visit 4) | DN (Visit 4) | OR: 5.70 (95% CI: 2.48 to 5.94) |  |  |
| Diabetes | Chahine and Al Souheil (2021) | No diabetes | Diabetes | aOR: 0.05 (95% CI: 0.00 to 0.62) | OR: 0.60 (95% CI: 0.15 to 2.42) | There was no difference in the likelihood of receiving opioids between people who have diabetes and people who do not have diabetes. |
|  | Gallaghan et al. (2019) |  |  | aOR: 1.38 (95% CI: 1.27 to1.50) |  |  |
|  | Lakkad et al. (2023) |  |  | aOR: 0.76 (95% CI: 0.68 to 0.85) |  |  |
| Mental health disorder | Boulanger et al. (2009) | No anxiety and/or depression (commercial insurance) | Anxiety and/or depression (commercial insurance) | OR: 1.54 (95% CI:1.42 to 1.67) | OR: 1.48 (95% CI: 1.36 to 1.2) | People with anxiety and/or depression were more likely to receive opioids for NeuP compared to people without depression.  It should be noted that Goswami et al. (2023) only compared groups that received opioids, or gabapentin, or both.The OR from Goswami et al. (2023) comes from univariate analysis while the other studies reported multivariate ORs. |
|  | Boulanger et al. (2009) | No anxiety and/or depression (Medicare supplemental insurance) | Anxiety and/or depression (Medicare supplemental insurance) | OR: 1.41 (95% CI: 1.29 to 1.53) |  |  |
|  | Chahine and Al Souheil (2021) | No depression | Depression | aOR: 1.00 (95% CI: 0.41 to 2.44) | OR: 1.18 (95% CI: 1.01 to 1.38) |  |
|  | Gallaghan et al. (2019) |  |  | aOR: 1.25 (95% CI: 1.01 to 1.55) |  |  |
|  | Goswami et al. (2023) |  |  | OR: 1.11 (95% CI: 0.87 to 1.41) |  |  |
|  | Chahine and Al Souheil (2021) | No anxiety | Anxiety | aOR: 1.00 (95% CI: 0.29 to 3.41) | OR: 1.18 (95% CI: 1.03 to 1.35) |  |
|  | Gallaghan et al. (2019) |  |  | aOR: 1.14 (95% CI: 0.97 to 1.33) |  |  |
|  | Goswami et al. (2023) |  |  | OR: 1.33 (95% CI: 0.99 to 1.78) |  |  |
| Comorbidity score | Gallaghan et al. (2019) | Lower Charlson score | Higher Charlson score | aOR: 1.15 (95% CI: 1.10 to 1.20) | NA | It is not clear how Charlson comorbidity score affects the likelihood of receiving opioids.  The results from Gallaghan et al. (2019) and Lin et al. (2023) suggest that increasing comorbidity increases the likelihood of receiving opioids. The results from Patil et al. (2015) and Goswami et al. (2023) found no difference in the likelihood of receiving opioids based on comorbidity level. |
|  | Goswami et al. (2023) | Lower Charlson score | Higher Charlson score | NS (groups compared by CCI mean) | NA |  |
|  | Patil et al. (2015) | 0 | 1 | OR: 0.43 (95% CI: 0.10 to 1.99) | OR: 1.08 (95% CI: 0.34 to 3.46) |  |
|  | Lin et al. (2023) |  |  | OR: 1.34 (95% CI: 1.09 to 1.65) |  |  |
|  | Patil et al. (2015) |  | 2 | OR: 0.92 (95% CI: 0.16 to 5.36) | OR: 1.20 (95% CI: 0.39 to 3.72) |  |
|  | Lin et al. (2023) |  |  | OR: 1.31 (95% CI: 1.02 to 1.69) |  |  |
|  | Patil et al. (2015) |  | 3 or above | OR: 2.30 (95% CI: 0.26 to 20.604) | OR: 1.80 (95% CI: 0.56 to 5.83) |  |
|  | Lin et al. (2023) |  |  | OR: 1.76 (95% CI: 1.41 to 2.20) |  |  |
| Other comorbidities | Patil et al. (2015) | No diabetes with organ damage | Diabetes with organ damage | OR: 9.08 (95% CI: 1.06 to 77.58) | NA | Having diabetes with organ damage or nephropathy increases the likelihood of receiving opioids.  In people with DN, comorbidities that did not have any change in the likelihood of receiving opioids included: cerebrovascular and peripheral vascular diseases, metabolic disorders, obesity, retinopathy, and cardiovascular diseases.  In people with DN, having organ damage or nephropathy increases the likelihood of receiving opioids as first-line treatment. In people with DN, comorbidities that did not affect the likelihood of receiving opioids as first-line treatment included: cerebrovascular and peripheral vascular diseases, metabolic disorders, obesity, and retinopathy. In people with DN, Charlson comorbidity score does not affect the likelihood of receiving opioids as first-line treatment. |
|  |  | No nephropathy | Nephropathy | OR: 13.25 (95% CI: 1.89 to 92.69) | NA |  |
|  |  | No cerebrovascular and/or peripheral vascular disease | Cerebrovascular and/or peripheral vascular disease | NS | NA |  |
|  |  | No metabolic disorder | Metabolic disorder | NS | NA |  |
|  |  | No retinopathy | Retinopathy | NS | NA |  |
|  |  | No cardiovascular disease | Cardiovascular disease | NS | NA |  |
|  |  | No obesity | Obesity | NS | NA |  |
| BMI | Marcianò et al. (2024) | <25 | 25-30 | NS | NA | There was no difference in the likelihood of receiving opioids based on BMI. |
|  |  |  | >30 |  | NA |  |
| Alcohol | Chahine and Al Souheil (2021) | Lower alcohol intake | Higher alcohol intake | aOR: 2.32 (95% CI: 1.01 to 5.35) | NA | Higher alcohol intake was associated with a higher likelihood of receiving opioids. |
| Smoking | Chahine and Al Souheil (2021) | Non-smoker | Smoker | aOR: 3.59 (95% CI: 1.46 to 8.81) | NA | Being a smoker was associated with a higher likelihood of receiving opioids. |

Table S17. Analysis of predictors of being prescribed non-opioid analgesics. Adjusted odds ratios are provided for variables that were significant in multivariable analyses where applicable. Statistically non-significant results are provided when they were needed for meta-analysis. MA: meta-analysis, OR: odds ratio, aOR: adjusted odds ratio, NS: not significant, NA: not applicable.

| Variable | Study | Reference | Comparison | Result | MA | Discussion |
| --- | --- | --- | --- | --- | --- | --- |
| Age | Chahine and Al Souheil (2021) | Younger | Older | aOR: 1.03 (95% CI: 1.01 to 1.05) | NA | Overall, it seems that there was no difference in the likelihood of receiving non-opioid analgesics based on age.  Johnston et al. (2014) found that in people with DN younger age was associated with a higher likelihood of receiving non-opioids analgesics, while this was opposite for people with PHN. Chahine and Al Souheil (2021) investigated variety of different NeuP diagnoses, although DN was most represented ~30%.  Johnston et al. (2014) included only people who were 65 years or older and the sample had a mean age of 76 years. Chahine and Al Souheil (2021) included people 23-88 years old, and the sample had a mean age of 50.2 years. Moreover, Johnston et al. (2014) had a much bigger sample size (N=30,428) than Chahine and Al Souheil (2021) (N=360). |
|  | Johnston et al. (2014) | >85 years (DN) | 65-74 years (DN) | OR: 1.48 (95% CI: 1.34 to 1.63) | OR: 0.88 (95% CI: 0.32 to 2.43) |  |
|  |  |  | 65-74 years (PHN) | OR: 0.52 (95% CI: 0.44 to 0.62) |  |  |
|  |  | >85 years (PHN) | 75-84 years (DN) | OR: 1.15 (95% CI: 1.04 to 1.27) | OR: 0.85 (95% CI: 0.58 to 1.26) |  |
|  |  |  | 75-84 years (PHN) | OR: 0.62 (95% CI: 0.52 to 0.73) |  |  |
| Sex | Chahine and Al Souheil (2021) | Male | Female | aOR: 0.45 (95% CI: 0.26 to 0.79) | NA | Being male was associated with a higher likelihood of receiving non-opioid analgesics. |
| Prescriber | Chahine and Al Souheil (2021) | Pharmacist | Physician | aOR: 0.10 (95% CI: 0.03 to 0.26) | NA | In Lebanon, people were more likely to receive non-opioid analgesics from a pharmacist than a physician. |
| Concomitant NeuP medication | Gore et al. (2011a) | Before pregabalin | After pregabalin | OR:1.01 (95% CI: 0.81 to 1.25) | OR: 1.11 (95% CI: 0.37 to 3.33) | Using concomitant gabapentin, pregabalin, or duloxetine with non-opioid analgesics was not associated with any change in the prescribing of non-opioid analgesics.  Duloxetine compliance was not associated with any change in the prescribing of non-opioid analgesics.  People with gabapentin were more likely to be prescribed concomitant tramadol compared to people with pregabalin. |
|  | Gore et al. (2011b) |  |  | OR: 1.20 (95% CI: 1.01 to 1.42) |  |  |
|  | Wu et al. (2009) | Low duloxetine compliance (commercial insurance) | High duloxetine compliance (commercial insurance) | OR: 1.21 (95% CI: 0.58 to 2.47) | OR: 0.11 (95% CI: 058 to 2.10) |  |
|  |  | Low duloxetine compliance (Medicare supplemental) | High duloxetine compliance (Medicare supplemental) | OR: 0.82 (95% CI: 0.20 to 3.28) |  |  |
|  | Zhao et al. (2011) | Duloxetine | Pregabalin | NS | NA |  |
|  | Han et al. (2023) | Pregabalin | Gabapentin | NR (~4% vs ~16%) | NA |  |
|  | Gore et al. (2011a) | Before duloxetine | After duloxetine | NS | NA |  |
|  | Gore et al. (2011b) | Before gabapentin | After gabapentin | NS | NA |  |
| Concomitant brand-name medication | Sicras-Mainar et al. (2015) | Brand-name gabapentin | Generic name gabapentin | OR: 2.48 (95% CI: 1.95 to 3.15) | NA | Compared to people using brand-name medication, people using generic medication were more likely to receive non-opioid analgesics. |
| NeuP diagnosis | Johnston et al. (2014) | DN | PHN | OR: 1.40 (95% CI: 1.31 to 1.50) | OR: 0.92 (95% CI: 0.43 to 1.97) | There was no difference in the likelihood of receiving non-opioid analgesics between people who have DN or PHN. Dieleman et al. (2008) was excluded from this meta-analysis because their samples sizes were unclear. In their study 22.9% of people with DN received non-opioid analgesics and 26.1% of people with PHN received non-opioid analgesics. It is unlikely that adding these results to the meta-analysis would produce a significant difference in the likelihood of receiving non-opioid analgesics between people who have DN or PHN.  There was no difference in the likelihood of receiving non-opioid analgesics between people who have PLP or PHN.  There was no difference in the likelihood of receiving non-opioid analgesics between people who have PLP or DN.  There was no difference in the likelihood of receiving non-opioid analgesics between people who have neuropathic back pain or DN. The results from Udall et al. (2019) most likely did not show a significant different difference due to a small sample (N=116).  People with neuropathic back pain were more likely to receive non-opioid analgesics compared to people with PHN.  There was no difference in the likelihood of receiving non-opioid analgesics between people who have mixed pain or pure NeuP.  There was no difference in the likelihood of receiving non-opioid analgesics between people who had SCI or did not have SCI. There was no difference in the likelihood of receiving non-opioid analgesics between people who had cervical/lumbar radiculopathy or did not have cervical/lumbar radiculopathy.  People with mononeuropathy were more likely to receive non-opioid analgesics compared to people with other NeuP diagnosis. |
|  | Udall et al. (2019) |  |  | OR: 2.07 (95% CI: 0.22 to 19.35) |  |  |
|  | Hall et al. (2008) |  |  | OR: 0.84 (95% CI: 0.70 to 1.02) |  |  |
|  | Hall et al. (2013) |  |  | OR: 0.57 (95% CI: 0.51 to 0.63) |  |  |
|  | Dieleman et al. (2008) |  |  | NA | NA |  |
|  | Hall et al. (2008) | PLP |  | OR: 1.15 (95% CI: 0.59 to 2.24) | OR: 0.61 (95% CI: 0.00 to 1432.26). Fixed effects model OR: 0.47 (95% CI: 0.32 to 0.70) |  |
|  | Hall et al. (2013) |  |  | OR: 0.34 (95% CI: 0.21 to 0.55) |  |  |
|  | Hall et al. (2008) | PLP | DN | OR: 1.36 (95% CI: 0.69 to 2.67) | OR: 0.87 (95% CI: 0.00 to 164.71). Fixed effects model OR: 0.76 (95% CI: 0.51 to 1.12) |  |
|  | Hall et al. (2013) |  |  | OR: 0.59 (95% CI: 0.36 to 0.97) |  |  |
|  | Udall et al. (2019) | DN | NeuP backpain | OR: 1.66 (95% CI: 0.70 to 3.90) | OR: 2.56 (95% CI: 0.92 to 7.11) |  |
|  | Hall et al. (2013) |  |  | OR: 2.60 (95% CI: 2.39 to 2.83) |  |  |
|  | Udall et al. (2019) | PHN |  | OR: 3.43 (95% CI: 0.39 to 29.95) | OR: 1.48 (95% CI: 1.05 to 2.07) |  |
|  | Hall et al. (2013) |  |  | OR: 1.48 (95% CI: 1.38 to 1.58) |  |  |
|  | Gore et al. (2007a) | Pure NeuP | Mixed pain | OR: 2.47 (95% CI: 2.36 to 2.59) | OR: 1.68 (95% CI: 0.01 to 244.98). Fixed effects model OR: 2.37 (95% CI: 2.27 to 2.48) |  |
|  | Gustavson et al. (2013) |  |  | OR: 1.13 (95% CI: 0.93 to 1.38) |  |  |
|  | Hall et al. (2008) | TN | DN | OR: 1.01 (95% CI: 0.84 to 1.22) | OR: 0.80 (95% CI: 0.71 to 0.91) |  |
|  |  |  | PHN | OR: 0.57 (95% CI: 0.43 to 0.76) |  |  |
|  |  |  | PLP | OR: 1.28 (95% CI: 0.65 to 2.51) |  |  |
|  | Chahine and Al Souheil (2021) | No DN | DN | NS | NA |  |
|  |  | No PHN | PHN | NS | NA |  |
|  |  | No cervical/ lumbar radiculopathy | Cervical/lumbar radiculopathy | NS | NA |  |
|  |  | No SCI | SCI | NS | NA |  |
|  |  | No neuropathic postoperative pain | Neuropathic postoperative pain | NS | NA |  |
|  |  | No TN | TN | NS | NA |  |
|  |  | No post traumatic neuralgia | Post traumatic neuralgia | NS | NA |  |
|  | Butler et al. (2018) | PTN | DN | NS | NA |  |
|  | Dieleman et al. (2008) | DN | PHN | 26.1% vs 22.9% (sample size unclear) | NA |  |
|  |  |  | CTS | 30.4% vs 22.9% (sample size unclear) | NA |  |
|  |  |  | Facial NeuP | 32.2% vs 22.9% (sample size unclear) | NA |  |
|  |  |  | Mononeuropathy | 40.7% vs 22.9% (sample size unclear) | NA |  |
| NeuP diagnosis year | Reed et al. (2013) | July 2002-June 2005 | July 2005-June 2008 | OR: 0.77 (95% CI: 0.65 to 0.91) | NA | People diagnosed with NeuP during July 2002-June 2005 were more likely to receive non-opioids compared to people diagnosed during July 2005-June 2008 or July 2008-June 2011. Similarly, people diagnosed with NeuP during July 2005-June 2008 were more likely to receive non-opioids compared to people diagnosed during or July 2008-June 2011. |
|  |  |  | July 2008-June 2011 | OR: 0.52 (95% CI: 0.43 to 0.63) | NA |  |
|  |  | July 2005-June 2008 | July 2008-June 2011 | OR: 0.67 (95% CI: 0.55 to 0.80) | NA |  |
| Diabetes | Chahine and Al Souheil (2021) | No diabetes | Diabetes | NS | NA | There was no difference in the likelihood of receiving non-opioid analgesics between people with or without diabetes. |
| Mental health disorder | Boulanger et al. (2009) | No anxiety and/or depression (commercial insurance) | Anxiety and/or depression (commercial insurance) | OR: 1.03 (95% CI: 0.94 to 1.12) | OR: 0.98 (95% CI: 0.92 to 1.05) | There was no difference in the likelihood of receiving non-opioid analgesics between people with or without anxiety and/or depression. |
|  |  | No anxiety and/or depression (Medicare supplemental insurance) | Anxiety and/or depression (Medicare supplemental insurance) | OR: 0.90 (95% CI: 0.80 to 1.01) |  |  |
|  | Chahine and Al Souheil (2021) | No anxiety | Anxiety | NS | NA |  |
|  |  | No depression | Depression | NS | NA |  |
| Alcohol | Chahine and Al Souheil (2021) | Lower alcohol intake | Higher alcohol intake | NS | NA | Higher alcohol intake was associated with a higher likelihood of receiving opioids. |
| Smoking | Chahine and Al Souheil (2021) | Non-smoker | Smoker | NS | NA | Being a smoker was associated with a higher likelihood of receiving opioids. |

Table S18. Analysis of predictors of being prescribed any pain medication. Adjusted odds ratios are provided for variables that were significant in multivariable analyses where applicable. Statistically non-significant results are provided when they were needed for meta-analysis. MA: meta-analysis, OR: odds ratio, aOR: adjusted odds ratio, PR: prevalence ratio, NS: not significant, NA: not applicable.

| Variable | Study | Reference | Comparison | Result | MA | Discussion |
| --- | --- | --- | --- | --- | --- | --- |
| Age | Mbrah et al. (2022) | 65-74 years | 50-64 years | PR: 1.10 (95% CI: 1.04 to 1.16) | NA | It was not possible to calculate a prevalence ratio from Pillay et al. (2015) data, thus it was not possible to conduct a meta-analysis from these two studies. In a large sample of people with different types of NeuP, increasing age was associated with being less likely to receive pharmacological treatment (Mbrah et al., 2022). In a small sample of people with HIV-associated NeuP, there was no association between age and the likelihood of receiving pain medication (Pillay et al., 2015). The mean age in Pillay et al. (2015) was 45.7 years, while Mbrah et al. (2022) included people aged 50 years or older and had the largest proportion in the category 85 years or older. Thus, it seems that younger age is associated with a higher likelihood of receiving pain medication in people who are 50 years or older, while there does not seem to be any difference in the likelihood of receiving pain medication in people who are below 50 years old. This division may be arbitrary and should be confirmed when further evidence is collected. |
|  |  | 75-84 years |  | PR: 1.24 (95% CI: 1.18 to 1.30) | NA |  |
|  |  | >85 years |  | PR: 1.45 (95% CI: 1.38 to 1.52) | NA |  |
|  | Pillay et al. (2015) | Older | Younger | NS | NA |  |
| Sex | Mbrah et al. (2022) | Female | Male | PR: 0.79 (95% CI: 0.77 to 0.81) | PR: 0.93 (95% CI: 0.65 to 1.34) | There was no difference in the likelihood of receiving any pain medication for NeuP between females and males.  In a large sample of people with NeuP, being male was associated with being less likely to receive pain medication (Mbrah et al., 2022). In a small sample of people with HIV-associated NeuP, there was no association between sex and the likelihood of receiving pain medication (Pillay et al., 2015).  A meta-analysis of these results found overall no difference in the likelihood of receiving pain medication for NeuP between females and males. However, it seems that females were more likely to receive pain medication than males. |
|  | Pillay et al. (2015) |  |  | PR: 1.15 (95% CI: 0.88 to 1.49) |  |  |
| Ethnicity | Mbrah et al. (2022) | White | SEVERAL | NS | NA | There was no difference in the likelihood of receiving pain medication based on ethnicity. |
| Education | Pillay et al. (2015) | Shorter education | Longer education | NS | NA | There was no difference in the likelihood of receiving pain medication based on education. |
| Other treatments | Pillay et al. (2015) | HIV treatment type | SEVERAL | NS | NA | The type of HIV treatment was not associated with any differences in the likelihood of receiving pain medication. |
| Pain | Pillay et al. (2015) | Mild pain intensity | Moderate pain intensity | OR: 3.38 (95% CI: 0.78 to 14.66) | NA | Higher pain intensity, and paraesthesia (tingling / feeling of pins and needles) were associated with a higher likelihood of receiving pain medication. The number of pain sites and numbness was not associated with any difference in the likelihood of receiving pain medication.  Higher pain severity was associated with a higher likelihood of receiving pain medication. |
|  |  |  | Severe pain intensity | OR: 8.46 (95% CI: 2.10 to 34.20) | NA |  |
|  |  | Lower number of pain sites | Higher number of pain sites | NS | NA |  |
|  |  | No paraesthesia | Paraesthesia | OR: 6.77 (95% CI: 2.04 to 22.44) | NA |  |
|  |  | No numbness | Numbness | NS | NA |  |
|  | Sadosky et al. (2013) | Mild pain severity | Moderate pain severity | 84.2% vs 54.5% | NA |  |
|  |  |  | Severe pain severity | 96.9% vs 54.5% | NA |  |
| NeuP diagnosis | Gore et al. (2007a) | Pure NeuP | Mixed pain | OR: 1.31 (95% CI: 1.25 to 1.37) | NA | Having mixed pain was associated with a higher likelihood of receiving pain medication. |
| Diabetes | Mbrah et al. (2022) | No diabetes | Diabetes | NS | NA | There was no difference in the likelihood of receiving pain medication for NeuP between people who had or didn’t have diabetes. |
| Mental health disorder | Boulanger et al. (2009) | No anxiety and/or depression (commercial insurance) | Anxiety and/or depression (commercial insurance) | OR: 3.86 (95% CI: 3.17 to 4.70) | OR: 3.66 (95% CI: 3.14 to 4.26) | People with anxiety and/or depression were more likely to receive pain medication compared to people without anxiety or depression. |
|  |  | No anxiety and/or depression (Medicare supplemental insurance) | Anxiety and/or depression (Medicare supplemental insurance) | OR: 3.38 (95% CI: 2.66 to 4.28) |  |  |
|  | Mbrah et al. (2022) | No anxiety | Anxiety | PR: 1.20 (95% CI: 1.18 to 1.23) | NA |  |
|  |  | No depression | Depression | PR: 1.47 (95% CI: 1.43 to 1.52) | NA |  |
| Other comorbidities | Mbrah et al. (2022) | Mild | No cognitive impairment | PR: 1.21 (95% CI: 1.17 to 1.25) | NA | Comorbidities that were associated with an increased likelihood of receiving pain medication included: anxiety, depression, multiple sclerosis, arthritis, and seizure disorder/epilepsy.  Comorbidities that were associated with a decreased likelihood of receiving pain medication included: cognitive impairment, Alzheimer's disease/dementia, cancer, fracture, and urinary tract infection.  Comorbidities that did not have any difference in the likelihood of receiving pain medication included: diabetes, HIV, cerebrovascular incidents, osteoporosis, heart failure, venous thromboembolism, peripheral vascular/arterial disease, coronary artery disease, and TB infection. |
|  |  | Moderate |  | PR: 1.47 (95% CI: 1.42 to 1.52) | NA |  |
|  |  | Severe |  | PR: 1.65 (95% CI: 1.57 to 1.73) | NA |  |
|  |  | Alzheimer’s disease or dementia | No Alzheimer’s disease or dementia | PR: 1.13 (95% CI: 1.10 to 1.16) | NA |  |
|  |  | Cancer | No cancer | PR: 1.16 (95% CI: 1.11 to 1.21) | NA |  |
|  |  | Coronary artery disease | No coronary artery disease | NS | NA |  |
|  |  | Heart failure | No heart failure | NS | NA |  |
|  |  | Venous thromboembolism | No venous thromboembolism | NS | NA |  |
|  |  | Peripheral vascular or arterial disease | No peripheral vascular or arterial disease | NS | NA |  |
|  |  | Cerebrovascular accident, transient ischemic attack, or stroke | No cerebrovascular accident, transient ischemic attack, or stroke | NS | NA |  |
|  |  | Multiple sclerosis | No multiple sclerosis | PR: 0.86 (95% CI: 0.78 to 0.95) | NA |  |
|  |  | HIV | No HIV | NS | NA |  |
|  |  | Arthritis | No arthritis | PR: 0.90 (95% CI: 0.88 to 0.92) | NA |  |
|  |  | Osteoporosis | No osteoporosis | NS | NA |  |
|  |  | Fracture | No fracture | PR: 1.11 (95% CI: 1.07 to 1.15) | NA |  |
|  |  | Seizure disorder or epilepsy | No seizure disorder or epilepsy | PR: 0.78 (95% CI: 0.75 to 0.82) | NA |  |
|  |  | Urinary tract infection | No urinary tract infection | PR: 1.03 (95% CI: 1.00 to 1.06) | NA |  |
|  | Pillay et al. (2015) | No current TB infection | Current TB infection | NS | NA |  |
| Laboratory measurements | Pillay et al. (2015) | Low CD4 T-cell count | High CD4 T-cell count | Median cell count: 369 (with medication) > 507 (no medication) | NA | In people with HIV-associated NeuP, low CD4 T-cell count was associated with a higher likelihood of receiving pain medication. |
| Rejects care | Mbrah et al. (2022) | Does not reject care | Rejects care | PR: 1.07 (95% CI: 1.02 to 1.12) | NA | People who rejected care were less likely to receive pain medication compared to those who did not reject care. |
| Daily activity dependence | Mbrah et al. (2022) | Independent in daily activities | Modified dependence | PR: 1.09 (95% CI: 1.06 to 1.12) | NA | People who were able to be independent in their daily activities were more likely to receive pain medication compared to those who were dependent on others in their daily activities. |
|  |  |  | Dependent in daily activities | PR: 1.09 (95% CI: 1.05 to 1.14) | NA |  |

Table S19. Analysis of predictors of adherence. The results are reported as probability for higher mean Medication Possession Ratio unless otherwise stated. Adjusted odds ratios are provided for variables that were significant in multivariable analyses where applicable. Statistically non-significant results are provided when they were needed for meta-analysis. MA: meta-analysis, OR: odds ratio, aOR: adjusted odds ratio, IRR: incidence rate ratio, NS: not significant, NA: not applicable.

| Variable | Study | Reference | Comparison | Result | MA | Discussion |
| --- | --- | --- | --- | --- | --- | --- |
| Age | Chen et al. (2010b) | 18-44 years | 45-54years | OR: 1.59 (95% CI: 0.97 to 2.61) | OR: 1.61 (95% CI: 1.13 to 2.29) | According to a meta-analysis, there was no difference in adherence to NeuP medications between older and younger people. In addition to Reynolds et al. (2020b), four other studies have investigated the association between age and adherence. Kato et al. (2023) et al. was the only other study that defined age in the same way (older vs younger). Kato et al. (2023) could not be included in this analysis because their results were reported as ORs rather than IRRs. Kato et al. (2023) showed that increasing age was associated with better mirogabalin adherence (OR=1.22, 95% CI=1.10-1.36). The results from Reynolds et al. (2020b) suggest that in the group with SNRIs (duloxetine or venlafaxine) increased age was associated with a slightly poorer adherence (note: non-significant IRR) unlike in the gabapentinoid (gabapentin or pregabalin) group. This discrepancy is unlikely to be due to the medications as the results from Chen et al. (2010b) and Wu et al. (2009) suggest that increasing age (particularly being 55-64 years old compared to 18-44 years old) is associated with better duloxetine compliance (OR=2.11, 95% CI=1.50-2.97). Furthermore, whilst it seems that older age is associated with better adherence, the results from Wu et al. (2009) suggest that after the age of 65 years the trend begins to decline. Their results showed that people aged 75-84 years old had significantly poorer adherence to duloxetine compared to people aged 65-74 years (OR=0.75, 95% CI=0.56-0.99). The comparison of people aged 75-84 years old and people aged 85 years or older was not significant (OR=0.84, 95% CI=0.48-1.48), which could be due to a small number of people in this group. The mean age in Reynolds et al. (2020b) SNRI group was 60.4 years (SD=15.0 years), so one would expect that increased age would be associated with better adherence in this group based on the evidence from other studies. Given that the IRR is very small and non-significant (IRR=0.98, 95% CI=0.97-1.00), it would be reasonable to conclude that altogether current evidence shows a trend of better adherence with increased age until 75 years. |
|  | Wu et al. (2009) |  |  | OR: 1.63 (95% CI: 0.99 to 2.70) |  |  |
|  | Chen et al. (2010b) |  | 55-64 years | OR: 2.10 (95% CI: 1.30 to 3.38) | OR: 2.11 (95% CI: 1.50 to 2.97) |  |
|  | Wu et al. (2009) |  |  | OR: 2.12 (95% CI: 1.30 to 3.46) |  |  |
|  | Yeh et al. (2021) |  | 45-64 years | OR: 1.02 (95% CI: 1.01 to 1.02) | NA |  |
|  | Wu et al. (2009) | 65-74 years | 75-84 years | OR: 0.75 (95% CI: 0.56 to 0.99) | NA |  |
|  |  |  | >85 years | NS | NA |  |
|  | Reynolds et al. (2020b) | Younger (gabapentinoids) | Older (gabapentinoids) | IRR: 1.01 (95% CI: 1.01 to 1.01) | IRR: 1.00 (95% CI: 0.97 to 1.03) |  |
|  |  | Younger (SNRIs) | Older (SNRIs) | IRR: 0.98 (95% CI: 0.97 to 1.00) |  |  |
|  | Kato et al. (2023) | Younger | Older | OR: 1.22 (95% CI: 1.10 to 1.36) | NA |  |
| Sex | Chen et al. (2010b) | Male | Female | OR: 1.43 (95% CI: 1.13 to 1.82) | OR: 0.99 (95% CI: 0.69 to 1.44) | There was no difference in adherence to NeuP medications between females and males. It should be noted that the OR from Kato et al. (2023) comes from univariate analysis while the other studies reported multivariate ORs. Reynolds et al. (2020b) could not be included in this analysis as they reported results as incidence rate ratios (IRRs) for adherence. Their results showed that in the sample of people using duloxetine or venlafaxine there was no difference in adherence between males and females. On the other hand, in their sample of people using pregabalin or gabapentin, men had significantly higher adherence (IRR=0.94, 95% CI=0.93-0.96). Overall, it seems that there is no difference in adherence to NeuP medications between females and males. However, the contrast between the results from Chen et al. (2010b) and Yeh et al. (2021) suggests that it is possible that there are significant differences in adherence between males and females in different populations. These trends could be analysed when further evidence is collected. |
|  | Yeh et al. (2021) |  |  | OR: 0.76 (95% CI: 0.67 to 0.86) |  |  |
|  | Kato et al. (2023) |  |  | OR: 0.93 (95% CI: 0.72 to 1.21) |  |  |
|  | Reynolds et al. (2020b) | Male (gabapentinoids) | Female (gabapentinoids) | IRR: 0.94 (95% CI: 0.93 to 0.96) | IRR: 0.97 (95% CI: 0.90 to 1.04) |  |
|  |  | Male (SNRIs) | Female (SNRIs) | IRR: 1.01 (95% CI: 0.96 to 1.07) |  |  |
| Ethnicity | Reynolds et al. (2020b) | White (gabapentinoids) | Black (gabapentinoids) | IRR: 0.89 (95% CI: 0.87 to 0.91) | IRR: 0.88 (95% CI: 0.86 to 0.89) | People with white ethnicity were more likely to adhere to their medication compared to people with other ethnicities.  White vs black IRR: 0.89 (0.87 to 0.91)  White vs Hispanic IRR: 0.87 (0.85 to 0.89)  White vs Asian IRR: 0.85 (0.82 to 0.88) |
|  |  |  | Asian (gabapentinoids) | IRR: 0.85 (95% CI: 0.82 to 0.89) |  |  |
|  |  |  | Hispanic (gabapentinoids) | IRR: 0.87 (95% CI: 0.85 to 0.89) |  |  |
|  |  | White (SNRIs) | Black (SNRIs) | IRR: 0.93 (95% CI: 0.85 to 1.02) |  |  |
|  |  |  | Asian (SNRIs) | IRR: 0.80 (95% CI: 0.61 to 1.04) |  |  |
|  |  |  | Hispanic (SNRIs) | IRR: 0.84 (95% CI: 0.76 to 0.92) |  |  |
| Geographic region | Chen et al. (2010b) | South USA | West USA | OR: 0.94 (95% CI: 0.64 to 1.39) | OR: 1.39 (95% CI: 0.65 to 3.00) | People living in the North Central USA were more likely to adhere to their medication compared to people living in the South USA. People living in the West South-Central USA were more likely to adhere to their medication compared to people living in the Middle Atlantic USA. Otherwise, there were no significant differences.  Results from meta-analyses using data from Kato et al. (2023) and Yeh et al. (2021):  People living in the North Central USA were more likely to adhere to their medication compared to people living in the South USA.  There was no difference in adherence to NeuP medications between people living in the West USA or Northeast USA and people living in the South USA.  Results from meta-analyses using data from Reynolds et al. (2020b):  People living in the West North Central USA, East North Central USA, or West South-Central USA were more likely to adhere to their medication compared to people living in the West South-Central USA.  There was no difference in adherence to NeuP medications between people living in the East South-Central USA and people living in the West South-Central USA. |
|  | Yeh et al. (2021) |  |  | OR: 2.06 (95% CI: 1.40 to 3.03) |  |  |
|  | Chen et al. (2010b) |  | North Central USA | OR: 1.23 (95% CI: 0.95 to 1.60) | OR: 1.24 (95% CI: 0.94 to 1.64) |  |
|  | Yeh et al. (2021) |  |  | OR: 1.24 (95% CI: 1.08 to 1.43) |  |  |
|  | Yeh et al. (2021) |  | Northeast USA | OR: 1.61 (95% CI: 0.96 to 2.70) | OR: 1.24 (95% CI: 0.94 to 1.64) |  |
|  | Kato et al. (2023) |  |  | OR: 1.15 (95% CI: 0.96 to 1.38) |  |  |
|  | Reynolds et al. (2020b) | West South-Central USA | East North-Central USA (gabapentinoids) | IRR: 1.03 (95% CI: 1.01 to 1.06) | IRR: 1.03 (95% CI: 1.01 to 1.05) |  |
|  |  |  | East North-Central USA (SNRIs) | IRR: 1.02 (95% CI: 0.93 to 1.12) |  |  |
|  |  |  | East South-Central USA (gabapentinoids) | IRR: 1.00 (95% CI: 0.97 to 1.03) | IRR: 1.00 (95% CI: 0.97 to 1.03) |  |
|  |  |  | East South-Central USA (SNRIs) | IRR: 1.00 (95% CI: 0.87 to 1.14) |  |  |
|  |  |  | Middle Atlantic USA (gabapentinoids) | IRR: 0.93 (95% CI: 0.90 to 0.96) | IRR: 0.93 (95% CI: 0.90 to 0.96) |  |
|  |  |  | Middle Atlantic USA (SNRIs) | IRR: 0.92 (95% CI: 0.81 to 1.05) |  |  |
|  |  |  | Mountain USA (gabapentinoids) | IRR: 1.02 (95% CI: 0.98 to 1.06) | IRR: 1.02 (95% CI: 0.97 to 1.06) |  |
|  |  |  | Mountain USA (SNRIs) | IRR: 0.95 (95% CI: 0.81 to 1.12) |  |  |
|  |  |  | New England USA | IRR: 1.04 (95% CI: 1.00 to 1.09) | IRR: 1.03 (95% CI: 0.99 to 1.08) |  |
|  |  |  | New England USA | IRR: 0.95 (95% CI: 0.81 to 1.12) |  |  |
|  |  |  | Pacific USA | IRR: 0.99 (95% CI: 0.98 to 1.01) | IRR: 0.99 (95% CI: 0.96 to 1.01) |  |
|  |  |  | Pacific USA | IRR: 0.94 (95% CI: 0.85 to 1.03) |  |  |
|  |  |  | South Atlantic | IRR: 0.98 (95% CI: 0.96 to 1.00) | IRR: 0.98 (95% CI: 0.96 to 1.00) |  |
|  |  |  | South Atlantic | IRR: 0.94 (95% CI: 0.86 to 1.02) |  |  |
|  |  |  | West North Central USA | IRR: 1.06 (95% CI: 1.03 to 1.09) | IRR: 1.07 (95% CI: 1.04 to 1.09) |  |
|  |  |  | West North Central USA | IRR: 1.09 (95% CI: 1.03 to 1.15) |  |  |
| Education | Reynolds et al. (2020b) | <12^th^ grade | High school diploma (gabapentinoids) | IRR: 1.07 (95% CI: 1.01 to 1.14) | IRR: 1.00 (95% CI: 0.81 to 1.23) | There was no difference in adherence to NeuP medications based on education. |
|  |  |  | High school diploma (SNRIs) | IRR: 0.85 (95% CI: 0.63 to 1.14) |  |  |
|  |  |  | Less than bachelor’s degree (gabapentinoids) | IRR: 1.08 (95% CI: 1.02 to 1.15) | IRR: 1.01 (95% CI: 0.83 to 1.24) |  |
|  |  |  | Less than bachelor’s degree (SNRIs) | IRR: 0.86 (95% CI: 0.64 to 1.16) |  |  |
|  |  |  | Bachelor’s degree plus (gabapentinoids) | IRR: 1.01 (95% CI: 0.95 to 1.08) | IRR: 0.98 (95% CI: 0.86 to 1.12) |  |
|  |  |  | Bachelor’s degree plus (SNRIs) | IRR: 0.84 (95% CI: 0.62 to 1.14) |  |  |
| Household income | Reynolds et al. (2020b) | <$40K | SEVERAL | NS | NA | There were differences in adherence based on the level of household income. |
| Insurance | Reynolds et al. (2020b) | PPO | EPO (gabapentinoids) | IRR: 0.94 (95% CI: 0.91 to 0.97) | IRR: 0.94 (95% CI: 0.91 to 0.97) | People with EPO or POS insurance were more likely to adhere to their medication compared to people with PPO insurance.  There was no difference in adherence to NeuP medications between people who have HMO, IND, high-deductible health plan, or 'other' insurance and people who have PPO insurance. |
|  |  |  | EPO (SNRIs) | IRR: 0.98 (95% CI: 0.87 to 1.11) |  |  |
|  |  |  | HMO (gabapentinoids) | IRR: 0.99 (95% CI: 0.97 to 1.01) | IRR: 0.99 (95% CI: 0.97 to 1.01) |  |
|  |  |  | HMO (SNRIs) | IRR: 0.96 (95% CI: 0.88 to 1.04) |  |  |
|  |  |  | IND (gabapentinoids) | IRR: 1.05 (95% CI: 1.01 to 1.10) | IRR: 1.05 (95% CI: 1.00 to 1.09) |  |
|  |  |  | IND (SNRIs) | IRR: 0.99 (95% CI: 0.81 to 1.12) |  |  |
|  |  |  | POS (gabapentinoids) | IRR: 0.93 (95% CI: 0.91 to 0.96) | IRR: 0.93 (95% CI: 0.91 to 0.95) |  |
|  |  |  | POS (SNRIs) | IRR: 0.93 (95% CI: 0.85 to 1.01) |  |  |
|  |  |  | High-deductible health plan (gabapentinoids) | IRR: 1.02 (95% CI: 0.98 to 1.06) | IRR: 1.02 (95% CI: 0.99 to 1.06) |  |
|  |  |  | High-deductible health plan (SNRIs) | IRR: 1.07 (95% CI: 0.95 to 1.21) |  |  |
|  |  |  | “Other” (gabapentinoids) | IRR: 1.01 (95% CI: 0.98 to 1.04) | IRR: 1.01 (95% CI: 0.98 to 1.04) |  |
|  |  |  | “Other” (SNRIs) | IRR: 0.99 (95% CI: 0.89 to 1.11) |  |  |
|  | Yeh et al. (2021) | PPO | SEVERAL | NS (reported as OR) | NA |  |
| Medication cost | Reynolds et al. (2020b) | Lower cost (gabapentinoids) | Higher cost (gabapentinoids) | IRR: 0.91 (95% CI: 0.89 to 0.93) | IRR: 0.92 (95% CI: 0.88 to 0.97) | A higher cost of medication to the individual was associated with poorer adherence. |
|  |  | Lower cost (SNRIs) | Higher cost (SNRIs) | IRR: 0.97 (95% CI: 0.88 to 1.07) |  |  |
| Type of medication | Reynolds et al. (2020b) | Venlafaxine | Duloxetine | MD: -0.02 (95% CI: NR/NA) | NA | There was no difference in adherence between people using duloxetine and people using venlafaxine.  People using duloxetine were more likely to adhere to their medication compared to people using gabapentin, pregabalin, or TCAs.  People using gabapentin were more likely to adhere to their medication compared to people using pregabalin.  There was no difference in adherence between people using gabapentin and people using TCAs.  (More comparisons between different medications would have been possible than what has been written here)  (Note that samples in Gharibian et al. (2013) were generally very small) |
|  | Gharibian et al. (2013) |  |  | % with ≥ 80% MPR, OR: 0.51 (95% CI: 0.15 to 1.78) | NA |  |
|  | Oladapo et al. (2012) | Gabapentin |  | MD: 0.11 (95% CI: 0.08 to 0.15) | MD: 0.09 (95% CI: 0.05 to 0.13) |  |
|  | Reynolds et al. (2020b) |  |  | MD: 0.07 (95% CI: 0.05 to 0.09) |  |  |
|  | Yang et al. (2015) |  |  | % with PDC ≥ 80% OR: 1.90 (95% CI: 1.58 to 2.29) | NA |  |
|  | Gharibian et al. (2013) |  |  | % with ≥ 80% MPR, OR: 1.46 (95% CI: 0.48 to 4.41) | NA |  |
|  | Zhao et al. (2011) | Pregabalin |  | MPR MD: 0.21 (95% CI: 0.17 to 0.25) | MPR MD: 0.16 (95% CI: 0.10to 0.22) |  |
|  | Oladapo et al. (2012) |  |  | MPR MD: 0.16 (95% CI: 0.12to 0.20) |  |  |
|  | Reynolds et al. (2020b) |  |  | MPR MD: 0.11 (95% CI: 0.09 to 0.13) |  |  |
|  | Yang et al. (2015) |  |  | % with PDC ≥ 80% OR: 2.30 (95% CI: 1.88 to 2.82) | NA |  |
|  | Zhao et al. (2011) |  |  | % with ≥ 80% MPR, OR: 2.45 (95% CI: 1.88 to 3.19) | % with ≥ 80% MPR, OR: 2.14 (95% CI: 1.01 to 5.80) |  |
|  | Gharibian et al. (2013) |  |  | % with ≥ 80% MPR, OR: 1.69 (95% CI: 0.42 to 6.72) |  |  |
|  | Oladapo et al. (2012) | TCAs |  | MD: 9.6% (95% CI: 7.53% to 11.67%), t=1.06, p<0.05 | NA |  |
|  | Gharibian et al. (2013) |  |  | % with ≥ 80% MPR, OR: 1.63 (95% CI: 0.54 to 4.90) | NA |  |
|  | Oladapo et al. (2012) | Pregabalin | Gabapentin | MD: 0.05 (95% CI: 0.01 to 0.09) | MD: 0.04 (95% CI: 0.03 to 0.05) |  |
|  | Reynolds et al. (2020b) |  |  | MD: 0.04 (95% CI: 0.03 to 0.05) |  |  |
|  | Yang et al. (2015) |  |  | % with PDC ≥ 80% OR: 1.21 (95% CI: 1.06 to 1.37) | NA |  |
|  | Gharibian et al. (2013) |  |  | % with ≥ 80% MPR, OR: 1.15 (95% CI: 0.48 to 2.57) | NA |  |
|  | Gharibian et al. (2013) | Venlafaxine |  | % with ≥ 80% MPR, OR: 0.35 (95% CI: 0.19 to 0.66) | NA |  |
|  | Giannopoulos et al. (2007) | Paroxetine or citalopram |  | % taking >75% of medication OR: 0.29 (95% CI: 0.07 to 1.13) | NA |  |
|  | Oladapo et al. (2012) | TCAs |  | MD: 1.7%, t=1.77, p>0.05 | NA |  |
|  | Gharibian et al. (2013) |  |  | % with ≥ 80% MPR, OR: 1.11 (95% CI: 0.89 to 1.38) | NA |  |
|  | Gharibian et al. (2013) | Pregabalin | Carbamazepine | % with ≥ 80% MPR, OR: 0.67 (95% CI: 0.18 to 2.43). Sample sizes n=22 vs n=19. | NA |  |
|  |  |  | Topiramate | % with ≥ 80% MPR, OR: 0.96 (95% CI: 0.33 to 2.77). Sample sizes n=22 vs n=40. | NA |  |
|  |  |  | Valproic acid | Too small sample for analysis (n=5) | NA |  |
|  |  |  | Lamotrigine | Too small sample for analysis (n=4) | NA |  |
|  | Reynolds et al. (2020b) | SNRI | Gabapentinoid | SNRIs: 55-57%, gabapentinoids: 44-48% | NA |  |
|  | Gharibian et al. (2013) | Antidepressants | Antiepileptics | % with ≥ 80% MPR OR: 0.99 (95% CI: 0.77 to 1.25) | NA |  |
| Dose | Chen et al. (2010b) | Duloxetine 60 mg | Duloxetine 30mg | % with MPR ≥ 80% OR: 0.45 (95% CI: 0.29 to 0.69) | NA | People using duloxetine had poorer adherence with 30 mg daily dose than 60 mg daily dose.  Pregabalin daily dose, or the starting dose of pregabalin or mirogabalin was not associated with any differences in adherence. |
|  |  |  | Duloxetine 31-59mg | % with MPR ≥ 80% OR: 1.54 (95% CI: 1.14 to 2.10) | NA |  |
|  |  |  | Duloxetine >60 mg | % with MPR ≥ 80% OR: 1.14 (95% CI: 1.02to 1.95) | NA |  |
|  | Sanchez et al. (2012) | Pregabalin <300 mg | Pregabalin 300-600 mg (DN) | % with PDC ≥ 80% OR: 1.15 (95% CI: 0.96 to 1.38) | % with PDC ≥ 80% OR: 1.11 (95% CI: 0.94 to 1.31) |  |
|  |  |  | Pregabalin 300-600 mg (PHN) | % with PDC ≥ 80% OR: 0.92 (95% CI: 0.62 to 1.37) |  |  |
|  | Yeh et al. (2021) | Starting dose of pregabalin more than 150 mg/day | Starting dose of pregabalin less than 150 mg/day | NS | NA |  |
|  | Kato et al. (2023) | Mirogabalin initial dose (regular) | Low | NS | NA |  |
|  |  |  | High | NS | NA |  |
| Dose titration | Yeh et al. (2021) | Pregabalin: no titration | Any dose titration within 45 days | % with MPR ≥ 80% OR: 2.59 (95% CI: 2.22 to 3.02) | OR: 2.40 (95% CI: 1.51 to 3.21) | People using pregabalin or mirogabalin had better adherence when the medication dose was titrated compared to those not having dose titration. Mirogabalin dose titration was associated with the biggest change in adherence when it was done during 16-30 days of initiating the treatment. |
|  | Kato et al. (2023) | Mirogabalin: no titration | Any dose titration within 45 days | % with MPR ≥ 80% OR: 1.75 (95% CI: 1.23 to 2.49) |  |  |
|  |  |  | Titration in days 16-30 | % with MPR ≥ 80% OR: 2.59 (95% CI: 1.29 to 5.13) | NA |  |
|  |  |  | Any dose titration | % with MPR ≥ 80% OR: 1.8 (95% CI: 1.26 to 2.57) | NA |  |
|  |  |  | Titration after day 45 | NS | NA |  |
|  |  |  | Titration with 15 days | NS | NA |  |
|  |  |  | Titration during days 31-45 | NS | NA |  |
| Brand-name medication | Sicras-Mainar et al. (2015) | Generic name medication (gabapentin) | Brand-name medication (Neurotin) | MD: 0.05 (95% CI: 0.01 to 0.09) | MD: 0.05 (95% CI: 0.03 to 0.07) | People using brand-name pregabalin or gabapentin had better adherence compared to people using generic pregabalin or gabapentin. |
|  | Sicras-Mainar et al. (2019) | Generic name medication (pregabalin) | Brand-name medication (Lyrica) | MD: 0.05 (95% CI: 0.02 to 0.08) |  |  |
| Medication reminder | Sutema et al. (2018) | No medicine reminder | Medicine reminder | RR for ≥80% adherence 2.38 (95% CI: 1.58 to 3.61) | NA | Having a medicine reminder was associated with better adherence. |
| Concomitant NeuP medication when using pregabalin | Yeh et al. (2021) | No concomitant medication | Lidocaine | MPR ≥ 80% OR: 1.50 (95% CI: 1.03 to 2.19) | NA | In people using pregabalin, concomitant lidocaine use was associated with better adherence. |
|  |  |  | SNRI | NS | NA |  |
|  |  |  | Gabapentin | NS | NA |  |
|  |  |  | Opioids excluding tramadol | NS | NA |  |
|  |  |  | Tramadol | NS | NA |  |
| Previous NeuP medication when using pregabalin | Yeh et al. (2021) | No previous NeuP medication | Lidocaine | NS | * | In people using pregabalin, previous use of gabapentin or TCAs was associated with better adherence.  In people using mirogabalin, no previous NeuP medications were associated with any differences in adherence.  *In meta-analyses combining the results from mirogabalin studies, none of the differences are significant. |
|  |  |  | SNRI | NS | * |  |
|  |  |  | Gabapentin | MPR ≥ 80% OR: 1.20 (95% CI: 1.07 to 1.36) | * |  |
|  |  |  | Opioids excluding tramadol | NS | * |  |
|  |  |  | Tramadol | NS | * |  |
|  |  |  | TCA | MPR ≥ 80% OR: 1.25 (95% CI: 1.04 to 1.51) | * |  |
| Previous NeuP medication when using mirogabalin | Kato et al. (2023) | No previous NeuP medication | pregabalin, neuropin, tramadol, duloxetine, opioids excluding tramadol, TCAs | NS | * | In people using mirogabalin, no previous NeuP medications were associated with any differences in adherence. |
|  |  |  | Gabapentin | OR: 0.21 (95% CI: 0.05 to 0.91) | * |  |
| Pain duration | Kato et al. (2023) | Shorter pain duration | Longer pain duration | NS | NA | Pain duration history was not associated with differences in mirogabalin adherence. |
| NeuP diagnosis | Sanchez et al. (2012) | PHN (therapeutic dose of pregabalin) | DN (therapeutic dose of pregabalin) | PDC MD: 0.15 (95% CI: 0.06 to 0.24) | PDC MD: 0.13 (95% CI: 0.10 to 0.16) | Compared to people with PHN, people with DN had better pregabalin adherence. |
|  |  | PHN (subtherapeutic dose of pregabalin) | DN (subtherapeutic dose of pregabalin) | PDC MD: 0.13 (95% CI: 0.10 to 0.16) |  |  |
| Comorbidity | Chen et al. (2010b) | Lower CCI | Higher CCI | OR: 0.98 (95% CI: 0.90 to 1.07) | OR: 0.97 (95% CI: 0.92 to 1.02) | A meta-analysis using data from Kato et al. (2023), Yeh et al. (2021), and Chen et al. (2010b) for CCI: There was no difference in adherence to NeuP medications between people with higher CCI and people with lower CCI. It should be noted that the OR from Kato et al. (2023) comes from univariate analysis while the other studies reported multivariate ORs.  A meta-analysis using data from Reynolds et al. (2020b) for CCI: People with a higher CCI were more likely to adhere to their medication compared to people with a lower CCI.  A meta-analysis of three other studies that had investigated the association of CCI, and adherence reported their results as ORs and thus were not combined with the results from Reynolds et al. (2020b). Given that the results from the OR meta-analysis suggest that there is no difference in adherence to NeuP medications between people with higher CCI and people with lower CCI, and that IRRs in this meta-analysis are very small, it is likely that there is no association between CCI and adherence. However, it may be best to leave the overall conclusion as ‘unclear’ until further evidence is collected. |
|  | Yeh et al. (2021) |  |  | OR: 0.94 (95% CI: 0.91 to 0.97) |  |  |
|  | Kato et al. (2023) |  |  | OR: 1.02 (95% CI: 0.95 to 1.09) |  |  |
|  | Reynolds et al. (2020b) | Lower CCI | Higher CCI | IRR: 1.02 (95% CI: 1.02 to 1.02) | IRR: 1.02 (95% CI: 1.02 to 1.02) |  |
|  |  | Lower CCI | Higher CCI | IRR: 1.02 (95% CI: 1.01 to 1.03) |  |  |
| Comorbidities | Chen et al. (2010b) | No migraine | Migraine | MPR ≥ 80% OR: 0.45 (95% CI: 0.20 to 0.99) |  | Most of the results suggest that comorbidities do not affect adherence. Migraine can decrease duloxetine adherence in people with DN. |
|  |  | No fibromyalgia | Fibromyalgia | NS | NA |  |
|  |  | No osteoarthritis | Osteoarthritis | NS | NA |  |
|  |  | No rheumatoid arthritis | Rheumatoid arthritis | NS | NA |  |
|  |  | No psoriatic arthropathy | Psoriatic arthropathy | NS | NA |  |
|  |  | No lower backpain | Lower backpain | NS | NA |  |
| Laboratory measurements | Kato et al. (2023) | eGFR chronic kidney disease classification (≥60ml/min/1.73m2) | SEVERAL | NS | NA | Chronic kidney disease classification or the level of renal function were not associated with any change in adherence. |
|  |  | Renal function based on creatine clearance (≥ 60ml/min) | SEVERAL | NS | NA |  |
| Treatment department | Kato et al. (2023) | Cancer/nerve disorder-related field | Neurology | NS | NA | The department where the person with NeuP is being treated was not associated with any differences in adherence. |
|  |  |  | Orthopaedics | NS | NA |  |
|  |  |  | General internal medicine | NS | NA |  |
|  |  |  | Other | NS | NA |  |
| Hospital visits | Yeh et al. (2021) | Lower number of inpatient days | Higher number of inpatient days | OR: 1.01 (95% CI: 1.00 to 1.02) | OR: 1.00 (95% CI: 0.98 to 1.02) | The number of inpatient days, outpatient visits, ER visits, or hospital admissions were not associated with any differences in adherence. |
|  | Kato et al. (2023) |  |  | aOR: 0.99 (95% CI: 0.99 to 0.99) |  |  |
|  | Yeh et al. (2021) | Lower number of outpatient visits | Higher number of outpatient visits | NS | NA |  |
|  |  | Lower number of ER visits | Higher number of ER visits | NS | NA |  |
|  |  | Lower number of hospital admissions | Higher number of hospital admissions | NS | NA |  |

Table S20. Analysis of predictors of discontinuation. Adjusted odds ratios are provided for variables that were significant in multivariable analyses where applicable. Statistically non-significant results are provided when they were needed for meta-analysis. MA: meta-analysis, OR: odds ratio, aOR: adjusted odds ratio, NS: not significant, NA: not applicable.

| Variable | Study | Reference | Comparison | Result | MA | Discussion |
| --- | --- | --- | --- | --- | --- | --- |
| Age | Yeh et al. (2021) | Younger | Older | HR: 0.99 (95% CI: 0.99 to 0.99) | HR: 0.96 (95% CI: 0.91 to 1.02) | Based on the meta-analysis, there was no difference in NeuP medication discontinuation between older and younger people. Individually these studies found that older age was associated with discontinuation, although the difference was very small. Three other studies (Sicras-Mainar et al., 2019; Toth et al., 2014; Shaparin et al., 2015) investigated the association between age and discontinuation and found no significant difference. |
|  | Kato et al. (2023) |  |  | HR: 0.93 (95% CI: 0.89 to 0.97) |  |  |
|  | Sicras-Mainar et al. (2019) |  |  | OR: 0.91 (95% CI: 0.77 to 1.00) | NA |  |
|  | Toth et al. (2014) |  |  | χ2:0.0 p=0.90 | NA |  |
|  | Shaparin et al. (2015) | <75 years | >75 years | NS | NA |  |
| Sex | Yeh et al. (2021) | Male | Female | HR: 1.20 (95% CI: 1.13 to 1.28) | HR: 1.08 (95% CI: 0.87 to 1.34) | There was no difference in NeuP medication discontinuation between females and males. |
|  | Kato et al. (2023) |  |  | HR: 0.96 (95% CI: 0.85 to 1.09) |  |  |
|  | Sicras-Mainar et al. (2019) |  |  | OR: 0.63 (95% CI: 0.53 to 0.77) | NA |  |
|  | Toth et al. (2014) |  |  | χ2 :0.7 p=0.42 | NA |  |
|  | Shaparin et al. (2015) |  |  | NR & NS | NA |  |
| Ethnicity | Shaparin et al. (2015) | White | Non-white | NS | NA | There was no difference in NeuP medication discontinuation based on ethnicity. |
| Geographic region | Yeh et al. (2021) | South USA | West USA | OR: 0.52 (95% CI: 0.42 to 0.66) | NA | People in the West USA were more likely to persist with treatment than people in the South USA. |
|  |  |  | SEVERAL | NS | NA |  |
| Insurance type | Yeh et al. (2021) | PPO | SEVERAL | NS | NA | There were no differences in NeuP medication discontinuation based on insurance types. |
| Prescribing process | Winterbottom et al. (2006) | By a physician | By clinical pharmacy algorithm | NS | NA | There were no differences in gabapentin persistence based on whether it was prescribed through a clinical pharmacy consult algorithm or by a physician. |
| NeuP medication | Gore et al. (2007a) | SEVERAL | SEVERAL | SEE FIGURE S1 | SEE FIGURE S1 | The differences in therapy duration between medications was investigated by ten studies. Gore et al. (2007a) (N= 30,999) and Hall et al. (2008) (N=5101) compared the most medications. Gore et al. (2007a) recorded treatment durations in mean days on medication while Hall et al. (2008) recorded the number of people with stable therapy after one year. The longest therapy duration measured in mean days on medication was 122 days in the group of people using SSRIs. The proportions of people with stable at one year were 15-30%. The results from both of these studies show people are more likely to discontinue TCAs compared to other antidepressants. Heatmaps showing the pairwise meta-analyses between all the investigated medications in these studies are shown in Figure S1 and S2.  Based on the mean treatment days reported by Gore et al. (2007a) and the pairwise meta-analysis heatmap, the medications they investigated can be ranked in the following order: SSRIs (126 days), 2nd generation antidepressants (122 days), antiepileptics (113 days), SNRIs (103 days), gabapentin (100 days), long-acting opioids (100 days), carbamazepine (93 days), COX 1 inhibitors (91 days), TCAs (91 days), NSAIDs (90 days), amitriptyline (87 days), opioids (80 days), COX 2 inhibitors (79 days), short-acting opioids (79 days). There was no significant difference between SSRIs and 2nd generation antidepressants (includes SSRIs and SNRIs), but they were used for a significantly longer time than antiepileptics, which in turn were used for a significantly longer time than SNRIs. There were no significant differences between SNRIs, gabapentin, long-acting opioids, and carbamazepine. COX 1 inhibitors, TCAs, and NSAIDs did not have a statistically significant difference compared to carbamazepine or long-acting opioids, but they were used for a significantly shorter time than gabapentinoids. Amitriptyline was used for an equally long time as TCAs and NSAIDs but were used for a significantly shorter time than COX 1 inhibitors. Opioids, COX 2 inhibitors, and short-acting opioids were used for an equally long times, but for a shorter time than amitriptyline.  Other studies investigating differences in discontinuation between medications showed that people using duloxetine were less likely to continue compared to people using pregabalin or gabapentin, whilst there does not seem to be difference between pregabalin or gabapentin. |
|  | Hall et al. (2008) | SEVERAL | SEVERAL | SEE FIGURE S2 | SEE FIGURE S2 |  |
|  | Mittal et al. (2011) | Duloxetine | Pregabalin | OR: 0.72 (95% CI: 0.37 to 1.42) | NA |  |
|  | Zhao et al. (2011) |  |  | Stopped during days 31-60 OR: 2.89 (95% CI: 2.39 to 3.48) | NA |  |
|  |  |  |  | Still with medication during days 331-365 OR: 33.5 (95% CI: 2.0 to 569.9) | NA |  |
|  | Yang et al. (2015) |  |  | Before day 360 OR: 1.82 (95% CI: 1.49 to 2.17) [Note more time points available] | NA |  |
|  |  |  | Gabapentin | Before day 360 OR: 1.40 (95% CI: 1.17 to 1.67) | NA |  |
|  | Johnson et al. (2013) | Pregabalin | Gabapentin | Mean treatment days, pregabalin 80, gabapentin 73, p>0.05 | NA |  |
|  | Banks et al. (2021) |  |  | Median treatment days, gabapentin 143, pregabalin 90 [statistical analysis not possible, gabapentin sample n=1079 while pregabalin n=142 | NA |  |
|  | Dragic et al. (2020) |  |  | OR: 5.80 (95% CI: 0.76 to 44.3) | OR: |  |
|  | Toth et al. (2014) |  |  | OR: 1.05 (95% CI: 0.21 to 5.29) |  |  |
|  | Muñoz-Vendrell et al. (2025) | Baclofen |  | NS | NA |  |
|  |  | Lacosamide |  | NS | NA |  |
|  | Toth et al. (2014) | Amitriptyline | Nortriptyline | OR: 0.79 (95% CI: 0.18 to 3.42) | NA |  |
|  | Gharibian et al. (2013) | Anticonvulsants | Antidepressants | NR (Kaplan-Meier curve in their manuscript, Figure 1) | NA |  |
|  | Dworkin et al. (2012) | First-line NeuP medication | Second-line NeuP medication | Mean treatment days, first-line 56, second-line 15.9, p<0.0001 | NA |  |
|  |  |  | Third-line NeuP medication | Mean treatment days, first-line 56, third-line 23.3, p=0.164 | NA |  |
| Brand-name NeuP medication | Sicras-Mainar et al. (2015) | Generic name medication (gabapentin) | Brand-name medication (Neurotin) | Mean treatment months, brand-name 7.3, generic 6.3, p<0.001 | NA | In people using pregabalin or gabapentin, brand-name medication is associated with better persistence compared to generic medication. |
|  | Sicras-Mainar et al. (2019) | Generic name medication (pregabalin) | Brand-name medication (Lyrica) | HR: 0.70 (95% CI: 0.58 to 0.85) | NA |  |
| Dose | Sanchez et al. (2012) | Subtherapeutic dose (DN) | Therapeutic dose (DN) | MD: 15.00 (95% CI: -1.63 to 31.63) | MD: 27.34 (95% CI: -0.95 to 55.62). Fixed effect model MD:23.99 (95% CI: 16.69 to 31.29) | There was no difference in NeuP medication discontinuation between people who had therapeutic dose and people who had subtherapeutic dose.  People with 30mg duloxetine were more likely to discontinue quicker compared to people with higher doses. However, people with 31-59 mg duloxetine discontinued slower than people with 60 mg duloxetine. People with >60 mg duloxetine discontinued with slowest rate. |
|  |  | Subtherapeutic dose (PHN) | Therapeutic dose (PHN) | MD: 17.00 (95% CI: 6.08 to 27.92) |  |  |
|  | Johnson et al. (2013) | Subtherapeutic dose (pregabalin) | Therapeutic dose (pregabalin) | MD: 26.35 (95% CI: 10.76 to 41.94) |  |  |
|  |  | Subtherapeutic dose (gabapentin) | Therapeutic dose (gabapentin) | MD: 54.85 (95% CI: 35.38 to 74.32) |  |  |
|  | Chen et al. (2010b) | Duloxetine 60mg | 30 mg | See Figure 1 in their manuscript | NA |  |
|  |  |  | 30-59 mg |  | NA |  |
|  |  |  | >60 mg |  | NA |  |
|  | Yeh et al. (2021) | Pregabalin starting dose above 150 mg | Pregabalin starting dose below 150 mg | NS | NA |  |
|  | Kato et al. (2023) | Mirogabalin starting dose (in patients with renal data): regular | High | NS | NA |  |
|  |  |  | Low | NS | NA |  |
|  | Hall et al. (2008) | SEVERAL | SEVERAL | % with stable therapy in 1 year [See Table 4 in their manuscript. Sample sizes ≤10] | NA |  |
| Dose titration | Yeh et al. (2021) | No dose titration | Dose titration | HR: 0.71 (95% CI: 0.62 to 0.81) | HR: 0.81 (95% CI: 0.63 to 1.03). Fixed effects model HR: 0.81 (95% CI: 0.80 to 0.91). | According to the meta-analysis, there was no difference in NeuP medication discontinuation between people who had dose titration and people who did not have dose titration. It should be noted that individually both of the studies found that having dose titration was associated with a lower likelihood of discontinuation. |
|  | Kato et al. (2023) |  |  | HR: 0.91 (95% CI: 0.84 to 0.98) |  |  |
| Combination pharmacotherapy | Kuo et al. (2016) | Monopharmacotherapy | Combination pharmacotherapy | HR: 1.30 (95% CI: 1.13 to 1.51) | NA | In people using pregabalin, concomitant use of SNRIs or opioids was associated with a higher the likelihood of pregabalin discontinuation. Concomitant use of gabapentin or lidocaine with pregabalin was not associated with any changes in pregabalin persistence. |
| Concomitant NeuP medication when using pregabalin | Yeh et al. (2021) | No concomitant medication | SNRI | HR: 0.69 (95% CI: 0.62 to 0.76) | NA |  |
|  |  |  | Opioids excluding tramadol | HR: 0.81 (95% CI: 0.72 to 0.90) | NA |  |
|  |  |  | Tramadol | HR: 0.65 (95% CI: 0.57 to 0.75) | NA |  |
|  |  |  | Gabapentin | NS | NA |  |
|  |  |  | Lidocaine | NS | NA |  |
| Concomitant NeuP medication when using opioids | Chen et al. (2010a) | Any other recommended NeuP medication | Duloxetine | Mean treatment days, any other medication 59.0, duloxetine 31.8, p>0.05 | NA | People using duloxetine were more likely to use SAOs for a shorter duration compared to people with other standard of care medications.  (Wu et al. (2011) did propensity score matching from Chen et al. (2010a) sample.) |
|  | Wu et al. (2011) |  |  | Mean treatment days, any other medication 51.4, duloxetine 26.9, p>0.05 | NA |  |
| Previous NeuP medication | Kato et al. (2023) | No gabapentin | Gabapentin | HR: 0.58 (95% CI: 0.14 to 2.36) | HR: 0.84 (95% CI: 0.71 to 1.01) | In people using pregabalin, previous medication with opioids excluding tramadol, lidocaine, TCAs, or SNRIs does was not associated with any changes in the likelihood of pregabalin discontinuation. Previous use of gabapentin decreased the likelihood of pregabalin discontinuation whereas previous use of tramadol increased the likelihood of pregabalin discontinuation. In people using mirogabalin, previous medication with pregabalin, neutropin, tramadol, duloxetine, opioids excluding tramadol, or TCA, (but not gabapentin) decreased the likelihood of discontinuation.  However, all of the meta-analyses combining pregabalin and mirogabalin analyses showed 'no significant difference' as the conclusion. |
|  | Yeh et al. (2021) |  |  | HR: 0.85 (95% CI: 0.71 to 1.01) |  |  |
|  | Kato et al. (2023) | No tramadol | Tramadol | HR: 0.23 (95% CI: 0.17 to 0.32) | HR: 0.50 (95% CI: 0.11 to 2.23) |  |
|  | Yeh et al. (2021) |  |  | HR: 1.09 (95% CI: 1.01 to 1.18) |  |  |
|  | Kato et al. (2023) | No opioids excluding tramadol | Opioids excluding tramadol | HR: 0.10 (95% CI: 0.04 to 0.25) | HR: 0.30 (95% CI: 0.04 to 2.33) |  |
|  | Yeh et al. (2021) |  |  | HR: 0.81 (95% CI: 0.72 to 0.91) |  |  |
|  | Kato et al. (2023) | No TCA | TCA | HR: 0.62 (95% CI: 0.48 to 0.80) | HR: 0.77 (95% CI: 0.52 to 1.13) |  |
|  | Yeh et al. (2021) |  |  | HR: 0.92 (95% CI: 0.83 to 1.01) |  |  |
|  | Yeh et al. (2021) | No lidocaine | Lidocaine | HR: 1.11 (95% CI: 1.00 to 1.23) | NA |  |
|  |  | No SNRI | SNRI | NS | NA |  |
|  | Kato et al. (2023) | No pregabalin | Pregabalin | HR: 0.31 (95% CI: 0.25 to 0.38) | NA |  |
|  |  | No duloxetine | Duloxetine | HR: 0.21 (95% CI: 0.14 to 0.32) | NA |  |
| NeuP medication effectiveness | Sicras-Mainar et al. (2019) | Not effective | Effective | OR: 1.2 (95% CI: 1.1 to 1.3) | NA | In people using pregabalin, those who experienced clinically significant pain relief were less likely to discontinue pregabalin compared to those who did not experience clinically significant pain relief. |
| Adverse events | Shaparin et al. (2015) | Mild AE | Moderate | aOR: 8.64 (95% CI: 4.42 to 16.89) | NA | In clinical trials, having moderate or severe adverse events increased the likelihood of discontinuing gabapentin. Nausea was the adverse event with the strongest association. |
|  |  |  | Severe | aOR: 5.37 (95% CI: 2.01 to 14.00) | NA |  |
|  |  | No nausea | Nausea | aOR: 4.93 (95% CI: 1.24 to 19.63) | NA |  |
|  |  | No somnolence | Somnolence | aOR: 4.22 (95% CI: 1.49 to 11.97) | NA |  |
|  |  | No headache | Headache | aOR: 3.52 (95% CI: 1.06 to 11.64) | NA |  |
|  |  | No dizziness | Dizziness | aOR: 3.17 (95% CI: 1.49 to 6.75) | NA |  |
| Pain intensity | Toth et al. (2014) | Low pain intensity | High pain intensity | NS | NA | Pain intensity was not associated with treatment persistence. |
| Pain duration | Toth et al. (2014) | Shorter pain duration | Longer pain duration | χ2: 22.6 p<0.01 | NA | Longer pain duration was associated with a higher likelihood of discontinuation. |
|  | Kato et al. (2023) |  |  | HR: 1.05 (95% CI: 1.04 to 1.08) | NA |  |
| NeuP diagnosis | Hall et al. (2006) | PLP | DN | MD: 12.80 (95% CI: -0.34 to 25.94) | MD: 28.48 (95% CI: -43.95 to 100.91) | People with DN / PLP were less likely to discontinue their NeuP medication compared to people with PHN.  There was no difference in NeuP medication discontinuation between people who have DN and people who have PLP.  There was no difference in NeuP medication discontinuation between people who have TN and people who have DN / PLP / PHN.  There was no difference in NeuP medication discontinuation between people who have mixed pain and people who have pure NeuP. |
|  | Hall et al. (2008) |  |  | MD: 4.00 (95% CI: -46.60 to 54.00) |  |  |
|  | Hall et al. (2013) |  |  | MD: 58.00 (95% CI: 44.17 to 71.83) |  |  |
|  | Hall et al. (2006) | PHN | PLP | MD: 15.70 (95% CI:3.18 to 28.22) | MD: 15.58 (95% CI: 11.56 to 19.50) |  |
|  | Hall et al. (2008) |  |  | MD: 22.60 (95% CI: -27.79 to 72.99) |  |  |
|  | Hall et al. (2013) |  |  | MD: 15.00 (95% CI: 2.27 to 27.73) |  |  |
|  | Hall et al. (2006) | DN | TN | MD: -26.70 (95% CI: -31.37 to -22.03) | MD: -7.20 (95% CI: -258.75 to 244.36). Fixed effects OR: -20.23 (95% CI: -24.50 to -15.96) |  |
|  | Hall et al. (2008) |  |  | MD: 12.90 (95% CI: 2.33 to 23.47) |  |  |
|  | Hall et al. (2006) | PHN |  | MD: 1.80 (95% CI: -0.66 to 4.26) | MD: 20.36 (95% CI: -219.12 to 259.84). Fixed effects MD: 4.16 (95% CI: 1.78 to 6.54) |  |
|  | Hall et al. (2008) |  |  | MD: 39.50 (95% CI: 29.99 to 49.01) |  |  |
|  | Hall et al. (2006) | PLP |  | MD: -13.90 (95% CI: -26.45 to -1.35) | MD: -8.68 (95% CI: -155.50 to 138.14). Fixed effects MD: -12.12 (95% CI: -24.30 to 0.06) |  |
|  | Hall et al. (2008) |  |  | MD: 16.90 (95% CI: -33.76 to 67.56) |  |  |
|  | Hall et al. (2006) | PHN | DN | MD: 28.50 (95% CI: 23.90 to 33.10) | MD: 50.97 (95% CI: 4.15 to 97.79) |  |
|  | Hall et al. (2008) |  |  | MD: 26.60 (95% CI: 17.39 to 35.81) |  |  |
|  | Hall et al. (2013) |  |  | MD: 73.00 (95% CI: 66.20 to 79.80) |  |  |
|  | Wang et al. (2020) |  |  | MD: 132.50 (95% CI: 126.43 to 138.57) |  |  |
|  | Sanchez et al. (2012) | PHN (therapeutic dose) | DN (therapeutic dose) | MD: 23.00 (95% CI: 4.20 to 41.80) |  |  |
|  |  | PHN (subtherapeutic dose) | DN (subtherapeutic dose) | MD: 21.00 (95% CI: 14.48 to 27.52) |  |  |
|  | Gore et al. (2007a) | Pure NeuP | Mixed pain | SEE FIGURE S3 | MD: 3.07 (95% CI: -5.91 to 12.04) |  |
|  | Gustavson et al. (2013) |  |  | NR (Kaplan-Meier curve in their manuscript, Figure 3) | NA |  |
| Neuropathic pain aetiology | Mittal et al. (2011) | Cryptogenic sensory polyneuropathy | Diabetes | OR: 0.28 (95% CI: 0.09 to 0.84) | NA | People with neuropathic pain caused by diabetes were less likely to discontinue pregabalin or duloxetine compared to people with neuropathic pain caused by cryptogenic sensory polyneuropathy. |
|  |  | Nerve entrapment |  | NS | NA |  |
|  |  | Inflammatory |  | NS | NA |  |
|  |  | Hereditary |  | NS | NA |  |
|  |  | Infection/toxic |  | NS | NA |  |
|  |  | Others |  | NS | NA |  |
| Mental health disorder | Toth et al. (2014) | Lower Beck Depression Inventory score | Higher Beck Depression Inventory score | NS | NA | Depression was not associated with any change in treatment persistence. |
| Comorbidity / disability score | Yeh et al. (2021) | Lower Charlson’s score | Higher Charlson’s score | HR: 0.97 (95% CI: 0.95 to 0.99) | HR: 0.97 (95% CI: 0.95 to 0.99) | People with more comorbidities were less likely to discontinue their treatment. Level of disability was not associated with any change in treatment persistence. People with a higher CCI were less likely to discontinue their NeuP medication compared to people with a lower CCI. |
|  | Kato et al. (2023) |  |  | HR: 0.96 (95% CI: 0.85 to 1.09) |  |  |
|  | Toth et al. (2014) | Lower EQ-5D disability index score | Higher EQ-5D disability index score | Pearson's correlation (NS) | NA |  |
| Laboratory measures | Kato et al. (2023) | Chronic kidney disease classification based on eGFR (≥60ml/min/1.73m2) | SEVERAL | NS | NA | Chronic kidney disease classification or the level of renal function were not associated with any change in the treatment persistence. |
|  |  | Renal function based on creatine clearance (≥ 60ml/min) | SEVERAL | NS | NA |  |
| Coping strategies | Toth et al. (2014) | Lower CSQ catastrophising score | Higher CSQ catastrophising score | Pearson’s correlation: -0.36 | NA | Pain-related worrying increased the likelihood of treatment discontinuation. Increased activity level and praying or hoping as coping behaviours were associated with increased persistence. |
|  |  | Lower activity level in CSQ | Higher CSQ catastrophising score | χ2: 6.5 (p=0.01) | NA |  |
|  |  | No praying / hoping in CQS | Praying / hoping in CQS | χ2: 8.2 (p<0.01) | NA |  |
| Hospital visits | Yeh et al. (2021) | Lower number of inpatient visits | Higher number of inpatient visits | HR: 1.00 (95% CI: 0.99 to 1.00) | HR: 0.99 (95% CI: 0.99 to 1.00) | The number of hospital visits was not associated with any change in pregabalin persistence. |
|  | Kato et al. (2023) |  |  | HR: 0.99 (95% CI: 0.99 to 0.99) |  |  |
|  | Yeh et al. (2021) | Lower number of outpatient visits | Higher number of outpatient visits | HR: 1.00 (95% CI: 0.96 to 1.04) | HR: 0.99 (95% CI: 0.99 to 1.00) |  |
|  | Kato et al. (2023) |  |  | HR: 0.99 (95% CI: 0.99 to 0.99) |  |  |
|  | Yeh et al. (2021) | Lower number of ER visits | Higher number of ER visits | NS | NA |  |
|  |  | Lower number of hospital admissions | Higher number of hospital admissions | NS | NA |  |
| Treatment department | Kato et al. (2023) | Cancer/nerve disorder-related field | Neurology | HR: 0.32 (95% CI: 0.23 to 0.46) | NA | Being treated at a neurology department was associated with a lower likelihood of mirogabalin discontinuation. |
|  |  |  | Orthopaedics | NS | NA |  |
|  |  |  | General internal medicine | NS | NA |  |
|  |  |  | Other | HR: 1.50 (95% CI: 1.17 to 1.93) | NA |  |

**Figure S1. Meta-analysis: discontinuation between different medications based on Gore et al. (2007a) data.** Heatmap of mean differences in mean treatment days between medications. Mean differences written in bold text were statistically significant. The figure is easiest read from x-axis to y-axis. For example, SNRIs had 12 days longer treatment duration than TCAs. Note that the data includes super- and subcategories, for example, any second-generation antidepressant would have included SNRIs and SSRIs. For more information about the medication categories see their Table 1.

**Figure S2. Meta-analysis: discontinuation between different medications based on Hall et al. (2008) data.** Heatmap of odds ratios for stable treatment at one year between medications. ** indicates statistically significant result. The figure is easiest read from x-axis to y-axis. For example, the odds ratio for having stable treatment at one year using ‘other antidepressants’ compared to TCAs was 1.56. Local anaesthetics, ‘other antidepressants’, and rubefacients/other topical antirheumatics had small sample sizes (N=22, N=131, N=228 respectively) while the sample sizes in other groups were 1029 to 1463.

**Figure S3. Meta-analysis: discontinuation between mixed pain and pure neuropathic pain based on Gore et al. (2007a) data.** Interpretation: There was no difference in NeuP medication discontinuation between people who have mixed pain and people who have pure NeuP. Reading example: People with pure NeuP have 13.74 days longer treatment duration using any opioids compared to people with mixed pain.

**Table S21. Summary of all results**. This table summarises all potential predictors investigated for pain medication prescribing, adherence, and discontinuation in adults with neuropathic pain. For each outcome, the potential predictors were categorised to either (1) increase the likelihood of that outcome, (2) not have an association with it, or (3) have inconsistent or unclear results. For more information about each variable please see Tables S13-S20. Recommended medication generally included serotonin-norepinephrine reuptake inhibitors, gabapentinoids, and tricyclic antidepressants. For the precise list of medications considered ‘recommended’ in each relevant study please see Table S5. ‘Non-opioid analgesic’ referred to non-steroidal anti-inflammatory drug, paracetamol, aspirin, or nefopam. List of all pain medication considered in the category ‘Any pain medication’ is provided in Table S3. DN: diabetic neuropathy; CKD: chronic kidney disease; CNS: central nervous system; COX: cyclooxygenase; CTS: carpal tunnel syndrome; eGFR: estimated glomerular filtration rate; EPO: exclusive provider organisation (only services in a specified network included); HMO: health maintenance organisation; IND: indemnity; NeuP: neuropathic pain; NSAID: non-steroidal anti-inflammatory drug; SCI: spinal cord injury; SEER: Surveillance, Epidemiology, and End Results; SNRI: serotonin-norepinephrine reuptake inhibitors; SSRI: selective serotonin reuptake inhibitors; PHN: postherpetic neuralgia; PLP: phantom limb pain; PPO: preferred provider organisation (also includes outside of network providers); POS: point-of-service; PSN: post-surgical neuropathic pain; PTN: post-traumatic neuropathic pain; TCA: tricyclic antidepressant; TN: trigeminal neuralgia. >: the variable on the left side of this sign were associated with the outcome compared to the variable on the right side of this sign. =: the variable on the left side of this sign was compared to the variable on the right side of this sign, but the comparison was not statistically significant. Variables following a colon (:) were investigated for the medication mentioned before the colon.

|  | **Recommended medication as the first choice** | **Recommended medication at any point** |
| --- | --- | --- |
| **Increases the likelihood of being prescribed** | Female > male  White ethnicity > non-white ethnicity  No previous opioid use for 3 months or longer  Previous antipsychotic use  Previous benzodiazepine use  Previous CNS depressant use  DN > PHN  Mental health disorder  Hemiplegia  Dementia  Diabetes  Rheumatologic disease  Breast cancer diagnosis year 2013 > 2007 | White ethnicity > non-white ethnicity  Diabetes  Mental health disorder  Depression  Anxiety  Living in a more deprived region (Townsend index)  Prescribed by physician > dispensed by a pharmacist  Treated by a pain specialist > not a pain specialist  Treated by a junior doctor > not a junior doctor  TN > DN  Pure NeuP > mixed pain  No previous opioid use for 3 months or longer  Previous antipsychotic use  Previous benzodiazepine use  Previous CNS depressant use  Higher Charlson comorbidity score  Mental health disorder  Hemiplegia  Dementia  Rheumatologic disease  Breast cancer diagnosis year 2013 > 2007  Lower eGFR  Lower low-density lipoprotein cholesterol  Higher body mass index  Lower alcohol intake |
| **No association** | Geographic region (USA)  Urban location = not urban location  Marital status  Education  Household income  Previous NSAID use  Previous long-acting opioid use  Previous muscle relaxant use  Previous steroid use  Substance use disorder  Non-malignant pain conditions  Myocardial infarction  Stroke  Congestive heart failure  Peptic ulcer  Peripheral vascular disease  Chronic obstructive pulmonary disease  Liver disease  Breast cancer treatment type  Age at breast cancer diagnosis  SEER grade  SEER summary status  Hormone receptor status | Age  Sex  Geographic region (USA)  Urban location = not urban location  Marital status  Insurance type  Smartset with best practice alert  Cervical/lumbar radiculopathy  SCI  PTN  PSN  Previous NSAID use  Previous long-acting opioid use  Previous muscle relaxant use  Previous steroid use  Substance use disorder  Non-malignant pain conditions  Myocardial infarction  Stroke  Congestive heart failure  Peptic ulcer  Peripheral vascular disease  Chronic obstructive pulmonary disease  Liver disease  Breast cancer treatment type  Age at breast cancer diagnosis  SEER grade  SEER summary status  Hormone receptor status  Diabetes duration  Diastolic blood pressure level  High-density lipoprotein cholesterol level  Total cholesterol level  Haemoglobin A1C level  Smoking status |
| **Inconsistent or unclear results** | NA | Education  Household income  Anxiety > no anxiety  DN > other NeuP diagnosis  Systolic blood pressure level |
|  | **Opioids as the first choice** | **Opioids at any point** |
| **Increases the likelihood of being prescribed** | Non-white ethnicity > white ethnicity  Previous opioid use for 3 months or longer  No previous antipsychotic use  No previous benzodiazepine use  No previous CNS depressant use  PLP > DN  PHN > DN  Neuropathic back pain > PHN  PHN > TN  PLP > TN  DN > TN  NeuP diagnosis year 2002-2005 > 2008-2011  No mental health disorder  No hemiplegia  No dementia  No diabetes  No rheumatologic disease  Diabetes with organ damage  Nephropathy  Breast cancer diagnosis year 2007 > 2013 | White ethnicity > Asian ethnicity  Treated by a pain specialist  Prescribed by a physician > dispensed by a pharmacist  Concomitant generic pregabalin > brand-name pregabalin  Previous opioid use for 3 months or longer  No previous antipsychotic use  No previous benzodiazepine use  No previous CNS depressant use  Low duloxetine compliance previously  Pain severity  PHN > DN  PHN > TN  PLP > TN  DN > TN  Neuropathic back pain > PHN  PSN > other NeuP diagnoses  No cervical/lumbar radiculopathy  Older year of NeuP diagnosis > more recent NeuP diagnosis year  Depression  Anxiety  Nephropathy  Diabetes with organ damage  No mental health disorder  No hemiplegia  No dementia  No rheumatologic disease  Breast cancer diagnosis year 2007 > 2013  Higher alcohol intake > lower alcohol intake  Smoking |
| **No association** | Sex  Age  Geographic region (USA)  Urban location = not urban location  Marital status  Education  Household income  Prescriber  PLP = PHN  Neuropathic back pain = DN  Previous NSAID use  Previous long-acting opioid use  Insulin use  Previous muscle relaxant use  Substance use disorder  Non-malignant pain conditions  Myocardial infarction  Stroke  Congestive heart failure  Peptic ulcer  Peripheral vascular disease  Cerebrovascular vascular disease  Cardiovascular disease  Chronic obstructive pulmonary disease  Liver disease  Metabolic disorders  Obesity  Retinopathy  Charlson comorbidity index  DN diagnosis year  Breast cancer treatment type  Age at breast cancer diagnosis  SEER grade  SEER summary status  Hormone receptor status | Age  Sex  White ethnicity > Hispanic ethnicity  White ethnicity = black ethnicity  Geographic region (USA)  Urban = rural  Marital status  Education  Additional pharmacotherapy guideline information with a best practice alert  Concomitant pregabalin prescription  Concomitant gabapentin prescription  Concomitant generic gabapentin = brand-name gabapentin  Dose titration of concomitant pregabalin  Previous NSAID prescription  Previous long-acting opioid prescription  Insulin use  SCI  Diabetes  Peripheral vascular disease  Cerebrovascular disease  Cardiovascular disease  Metabolic disorders  Obesity  Retinopathy  Substance use disorder  Non-malignant pain conditions  Myocardial infarction  Stroke  Congestive heart failure  Peptic ulcer  Chronic obstructive pulmonary disease  Liver disease  Breast cancer treatment type  Age at breast cancer diagnosis  SEER grade  SEER summary status  Hormone receptor status  BMI |
| **Inconsistent or unclear results** | NA | Household income  Insurance type  Concomitant duloxetine prescription  Mixed pain > pure NeuP  Neuropathic back pain > DN  Higher Charlson comorbidity index |
|  | **Non-opioid analgesics** | **Any pain medication** |
| **Increases the likelihood of being prescribed** | PHN: Older age > younger age  DN: Younger age > older age  Male > female  Concomitant generic gabapentin > brand-name gabapentin  Concomitant generic pregabalin > brand-name pregabalin  Dispensed by a pharmacist > prescribed by a physician  Mononeuropathy > other NeuP diagnosis  Neuropathic back pain > PHN  PHN > TN  Older year of NeuP diagnosis | Younger age > older age (in people > 50 years old)  Higher pain intensity  Paraesthesia  Mixed pain > pure NeuP  Anxiety  Depression  No cognitive impairment  No Alzheimer's disease/dementia  No cancer  Multiple sclerosis  Arthritis  No fractures  Seizure disorder/epilepsy  No urinary tract infection  Low CD4 T-cell count  Does not reject care in a nursing home  Independent in daily activities |
| **No association** | Concomitant gabapentin prescription  Concomitant duloxetine prescription  Concomitant pregabalin prescription  Concomitant duloxetine compliance  SCI  Cervical/lumbar radiculopathy  DN = PHN  Mixed pain > pure NeuP  Neuropathic back pain > DN  PLP = PHN  PLP = DN  PLP = TN  DN = TN  Anxiety  Depression  Diabetes  Alcohol intake  Smoking | Older age = younger age (in people <50 years old)  Female > male  Ethnicity  Education  Usual care = prescribing decision support algorithm  Type of HIV treatment  Number of pain sites  Numbness  Diabetes  Heart failure  Venous thromboembolism  Peripheral vascular or arterial disease  Cerebrovascular incidents  HIV  Osteoporosis  Current tuberculosis infection  Coronary artery disease |
| **Inconsistent or unclear results** | NA | NA |
|  | **Adherence** | **Discontinuation** |
| **Increases the likelihood** | Older (until 74 years) > younger  White ethnicity > non-white ethnicity  North Central USA > South USA  Middle Atlantic USA > West South-Central USA  EPO insurance > PPO insurance  POS insurance > PPO insurance  Lower medication cost to the patient  Duloxetine > pregabalin  Duloxetine > gabapentin  Gabapentin > pregabalin  Venlafaxine > TCAs  Duloxetine > TCAs  Duloxetine dose above 30 mg  Dose titration  Brand-name medication > generic medication  Medicine reminder  Pregabalin: concomitant lidocaine  Pregabalin: previous gabapentin  Pregabalin: previous TCAs  DN > PHN  No migraine > migraine | South USA > West USA  TCAs > other antidepressants  SNRIs > SSRIs  Amitriptyline > gabapentin  Pregabalin > duloxetine  Short-acting opioids > long-acting opioids  COX2 inhibitors > COX1 inhibitors  Generic medication > brand-name medication  Duloxetine dose 30 mg > above 30 mg  2^nd^-line medication > 1^st^/3^rd^-line medication  Combination pharmacotherapy > monopharmacotherapy  Pregabalin: no concomitant medication > concomitant SNRIs  Pregabalin: no concomitant medication > concomitant tramadol  Pregabalin: no concomitant medication > concomitant opioids  Short acting opioids: concomitant medication other than duloxetine  Pregabalin: no previous gabapentin use > previous gabapentin use  Pregabalin: previous tramadol use > no previous tramadol use  Mirogabalin: no previous NeuP medication > previous NeuP medication  Not experiencing clinically significant pain relief from NeuP medication  Experiencing moderate adverse events from NeuP medication  Experiencing severe adverse events from NeuP medication  Nausea as an adverse event from NeuP medication  Somnolence as an adverse event from NeuP medication  Headache as an adverse event from NeuP medication  Dizziness as an adverse event from NeuP medication  PHN > DN  PHN > PLP  Neuropathic back pain > DN  PLP > neuropathic back pain  PHN > neuropathic back pain  Pain duration  Lower Charlson comorbidity index  Pain-related worrying  Lower activity level  Not using praying or hoping as coping behaviours  Treatment department other than neurology |
| **No association** | Sex  Education  Household income  HMO insurance = PPO insurance  IND insurance = PPO insurance  High-deductible health plan = PPO insurance  Venlafaxine > duloxetine  Initial dose of NeuP medication  Pregabalin: concomitant/previous SNRIs  Pregabalin: concomitant gabapentin  Pregabalin: previous lidocaine  Pregabalin: concomitant/previous opioids  Pregabalin: concomitant/previous tramadol  Mirogabalin: any previous NeuP medication  Fibromyalgia  Osteoarthritis  Rheumatoid arthritis  Psoriatic arthropathy  Lower back pain  CKD classification based on eGFR  Renal function based on creatine clearance  Treatment department  Hospital visits | Age  Sex  Ethnicity  Insurance type  Clinical pharmacy consult algorithm  Pregabalin: ≥300 mg/day > <300 mg/day  Pregabalin: ≥150 mg/day > <150 mg/day  Gabapentin: ≥1800 mg/day > <1800 mg/day  Initial dose of NeuP medication  No dose titration > dose titration  Experiencing mild adverse events from NeuP medication^24^  Pain intensity  PLP = DN  TN = DN  TN = PHN  TN = PLP  Mixed pain > pure NeuP  Mirogabalin: previous gabapentin use = no previous gabapentin use  Pregabalin: previous opioid use = no previous opioid use  Pregabalin: previous lidocaine use = no previous lidocaine use  Pregabalin: previous TCA use = no previous TCA use  Pregabalin: previous SNRI use = no previous SNRI use  Disability  Depression  CKD classification based on eGFR  Renal function based on creatine clearance  Hospital visits |
| **Inconsistent or unclear results** | Pain duration  Pregabalin dose  Lower Charlson comorbidity index | Gabapentin > pregabalin  TCAs > antiepileptics  Opioids = TCAs  Opioids = antiepileptics |
